# Assessing the Impact of Pretraining Domain Relevance on Large Language Models Across Various Pathology Reporting Tasks

**DOI:** 10.1101/2023.09.10.23295318

**Authors:** Yunrui Lu, Gokul Srinivasan, Sarah Preum, Jason Pettus, Matthew Davis, Jack Greenburg, Louis Vaickus, Joshua Levy

## Abstract

Deep learning (DL) algorithms continue to develop at a rapid pace, providing researchers access to a set of tools capable of solving a wide array of biomedical challenges. While this progress is promising, it also leads to confusion regarding task-specific model choices, where deeper investigation is necessary to determine the optimal model configuration. Natural language processing (NLP) has the unique ability to accurately and efficiently capture a patient’s narrative, which can improve the operational efficiency of modern pathology laboratories through advanced computational solutions that can facilitate rapid access to and reporting of histological and molecular findings. In this study, we use pathology reports from a large academic medical system to assess the generalizability and potential real-world applicability of various deep learning-based NLP models on reports with highly specialized vocabulary and complex reporting structures. The performance of each NLP model examined was compared across four distinct tasks: 1) current procedural terminology (CPT) code classification, 2) pathologist classification, 3) report sign-out time regression, and 4) report text generation, under the hypothesis that models initialized on domain-relevant medical text would perform better than models not attuned to this prior knowledge. Our study highlights that the performance of deep learning-based NLP models can vary meaningfully across pathology-related tasks. Models pretrained on medical data outperform other models where medical domain knowledge is crucial, e.g., current procedural terminology (CPT) code classification. However, where interpretation is more subjective (i.e., teasing apart pathologist-specific lexicon and variable sign-out times), models with medical pretraining do not consistently outperform the other approaches. Instead, fine-tuning models pretrained on general or unrelated text sources achieved comparable or better results. Overall, our findings underscore the importance of considering the nature of the task at hand when selecting a pretraining strategy for NLP models in pathology. The optimal approach may vary depending on the specific requirements and nuances of the task, and related text sources can offer valuable insights and improve performance in certain cases, contradicting established notions about domain adaptation. This research contributes to our understanding of pretraining strategies for large language models and further informs the development and deployment of these models in pathology-related applications.

## Introduction

Pathology is a branch of medicine that focuses on the etiology and progression of disease across various organ systems. Nuanced pathological examinations can inform nearly all aspects of patient care, from the diagnosis of cancer to the management of acute and/or chronic diseases. In general, the role of most anatomic pathologists is to identify characteristics and patterns of cells within tissue (histology) or across a cytological specimen (cytology) for pathological changes^1^.

Currently, digital technologies have enabled the capture of microscopic examination results and descriptions of histomorphological features through whole slide images (WSI), as well as textual reports, respectively. While a significant amount of artificial intelligence research has concentrated on image analysis techniques for WSI, attention has only recently turned toward text analysis^234^. Deep learning (DL) methods, which are computational heuristics inspired by processes of the central nervous system, excel at processing imaging data to predict the risk of lung cancer^5^, recognize melanoma within dermoscopic images^6^, and segment digitized kidney tissue sections^7^, automatically detect early signs of colorectal cancer during colonoscopies^8^, and more recently to flexibly encode and interpret biomedical data including clinical language, imaging, and genomics^9^, amongst other tasks. Through the use of specialized and updatable image filters/shapes, which are used to localize imaging features through their optimal alignment, these algorithms are commonly used for binary and multi-class classification tasks^10,11^, and the localization of heterogenous cell lineages within distinct spatial architectures to inform the pathological assessment^12^.

While DL methods have shown great promise in the case of image data, real-world pathology is complex, and may require a more nuanced description of findings with a differential diagnosis (rather than a single diagnosis). Eventually, real-world application of computer vision algorithms may require leveraging textual description of the case’s particularities to highlight relevant histologic/molecular findings that suffer from loss of information when reported through brief synoptics which are more standardized than diagnostic/discussion text. This challenge is non-trivial, as pathologists record highly descriptive and specific observations through syntactically complex text^13,14^, making it difficult to train algorithms to complete this task. Variations in reporting could indicate a pathologist’s level of comfort, expertise, and complexity of the case. This may also be related to the time it takes to sign off on a case or how a hospital generates revenue using current procedural terminology (CPT) codes, which are interpreted by billing staff. Any potential deviations from standardized and efficient reporting criteria, possibly due to ambiguity in report write-ups, could have an impact on operational efficiency, revenue streams, and pathologists’ compensation. Nonetheless, the pathology report is a vital tool in determining diagnosis, prognosis, and treatment^15^, and more research is necessary to develop algorithms capable of reliably assessing these reports and studying differences in reporting practices by pathologists.

Though limitations exist, natural language processing (NLP) has proven to be broadly useful in healthcare, from clinical decision support to public health and pathology reports^16,17^. In pathology, algorithms have been developed to extract and structure textual reports^18^, address potential underbilling based on the misassignment of CPT codes after interpretation of pathology reports^19^, and standardize the language and style used in reports^19^.

Large language models (LLM) represent recent advances in artificial intelligence and are poised to significantly impact NLP’s application in pathology. LLMs are advanced AI systems trained on vast amounts of text data to generate human-like responses and perform language-related tasks. These algorithms utilize sophisticated deep neural networks which dynamically identify long-range syntactic and semantic dependencies to unpack the complexity of natural text^20^. The process of pretraining is essential for constructing these models, as it entails training on an extensive collection of publicly accessible domain-general text. During pretraining, the model predicts the next word within a given context, allowing it to comprehend language as used in the setting of interest, encompassing grammar, syntax, semantics, and cultural subtleties^21^. In short, pretraining enhances the model’s understanding of language from diverse dataset^21,22^. As data is becoming increasingly siloed, pretraining of LLMs can facilitate the transfer of knowledge from a source dataset to specific tasks, enabling adaptation to various applications without extensive task-specific training or expensive and laborious data collection^23^. By considering the sentence context and capturing long-range dependencies, pretraining improves understanding and coherence in generated text^24,20^. Pretrained models also demonstrate few-shot learning capabilities, enabling them to generalize from limited amounts of task-specific data^25^. They even showcase competence in handling unseen tasks, a phenomenon known as zero-shot learning^25^. This adaptability makes them efficient for new tasks and domains^21^. Through pretraining, large LLMs have been employed to suggest text to pathologists during the generation of pathology reports, which has the potential to enhance the speed at which reports are written^26,27,28^.

Comparing pretraining strategies for LLMs in pathology tasks may enhance our understanding of mechanisms to ameliorate expensive annotation by embedding attribution analysis and how pathology reporting text relates to other textual domains. In this study, we aim to assess the performance of several NLP LLMs across a range of pathology-related tasks from the context of domain adaptation to better determine appropriate model choices for a given research task. It is often thought that models initialized on information from similar sources will perform adeptly and require less data when adapted to a slightly different domain. Accordingly, we aim to evaluate the strengths and weaknesses of initializing models on corpora more germane to pathology, compared to data that is orthogonal or even unrelated to pathology reporting under the hypothesis that medical text will enhance predictions across all tasks. Our findings can enable researchers and practitioners to choose the appropriate approach for their specific pathology-related tasks and will reveal the advantages and disadvantages of medical-domain relevant pretraining strategies. We investigate four distinct tasks across various applications: 1) increase hospital revenue by identifying instances of underbilling through CPT code classification (CPT code prediction), 2) study variations in reporting patterns by classifying pathologist’s reporting patterns (pathologist classification), 3) informing case complexity/pathologist workload through prediction of sign-out time for potential assignment of relative value units (RVUs) (sign-out time prediction), and 4) text generation to improve the efficiency of report writing (pathologist report generation). Our results highlight which model approach and pretraining strategy is most appropriate for each task assessed.

## Methods

### Methods Overview

The primary goal of this work is to compare LLM models’ performances for different tasks given different model architectures and pretraining approaches. Our workflow is as follows (Figure 1):

**Figure 1.**
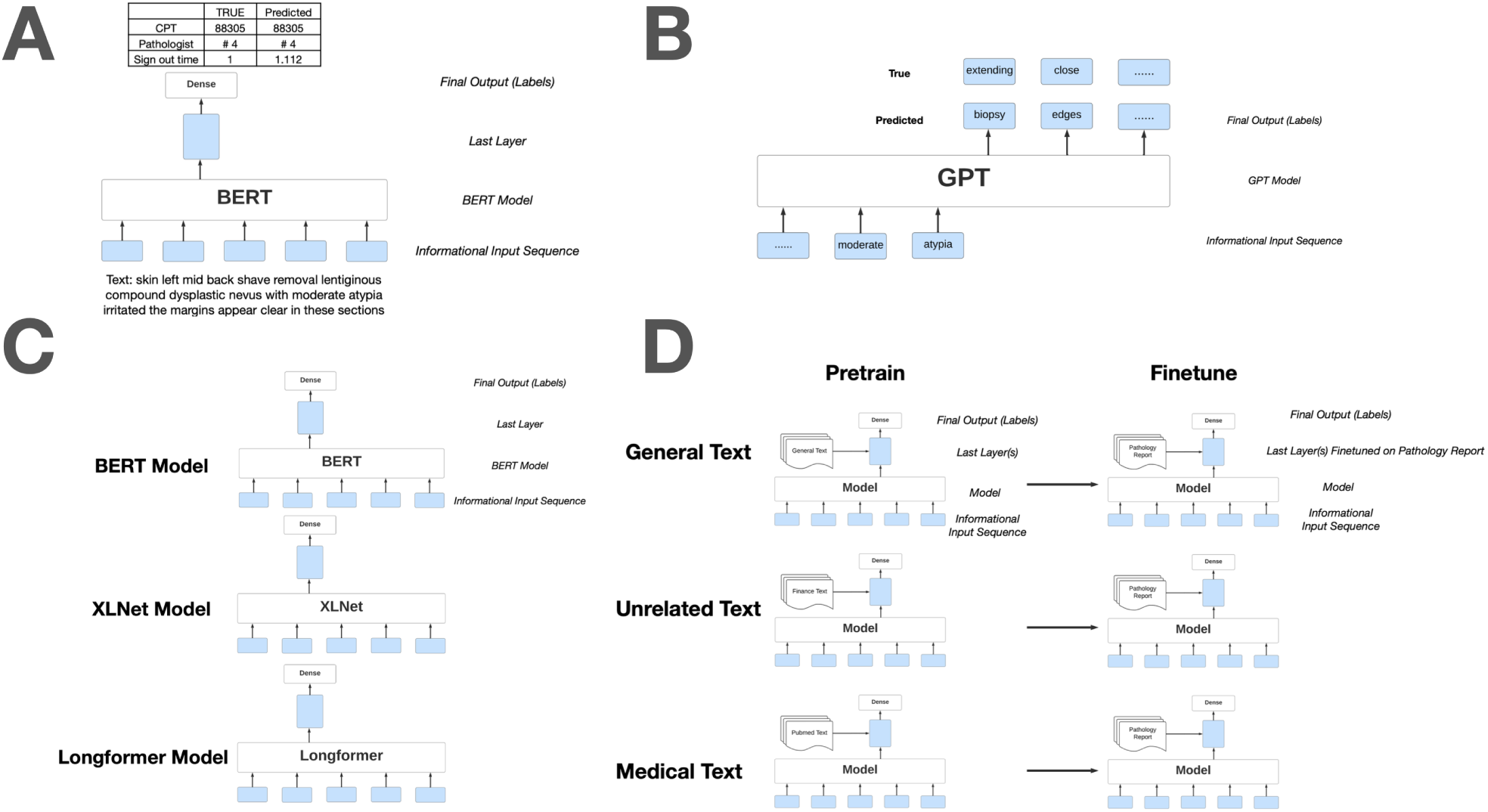
Experiment design workflow: A) Classification task; B) Text generation task; C) Comparison of different models; D) Comparison of different pretraining corpus

**Data preprocessing:** We collected 93,039 pathology reports as detailed in a previous study^19^, with corresponding CPT codes, and recorded the pathologists who signed out the reports and the sign-out time.

**Tasks include:** 1) CPT code prediction, 2) pathologist classification, 3) sign-out time prediction, 4) pathology report generation.

**Models architecture comparison:** Deep Learning and transformer models served as the primary model architectures– i,e, BERT, XLNet, Longformers, GPT, etc.

**Comparison of three pretraining strategies:** Models were initialized based on three separate text corpora: 1) General text; 2) Medical text; 3) Unrelated text, as means to determine which corpus could improve our modeling results. And the models are then fine-tuned on pathology reports.

### Data Preprocessing

Two datasets from a large academic medical center were accessed for this study: Dataset 1 (assigned CPT codes) and Dataset 2 (Dendrite). CPT codes (Dataset 1) were collected from June 2015 to June 2020^19^, with the dataset consisting of a 93,039 reports containing diagnostic text and the corresponding CPT code. The second dataset was collected from December 2011 to December 2021, containing 749,136 reports. The diagnostic text and sign-out time were extracted from these reports. Deidentification and text preprocessing protocols were similar to a previous study^19^. Data was divided into training, validation, and test sets using 80%, 10%, 10% percentage splits. Using these datasets, models were trained for the following tasks and evaluated:

### Tasks

#### Task #1, Primary CPT code classification

Pathology reports were assigned primary CPT codes based on subjective interpretation by hospital billing staff. Primary CPT codes partially reflect case complexity and includes 88300, 88302, 88304, 88305, 88307, and 88309. Primary CPT codes (e.g., CPT 88300, 88302, 88304, 88305, 88307, and 88309) are assigned based on the pathologist’s examination of the specimen. CPT 88300 represents an examination without requiring the use of a microscope (gross examination). CPT codes 88302-88309 include gross and microscopic examination of the specimen and are ordered by the case’s complexity level (as specified by the CPT codebook; an ordinal outcome; e.g., CPT 88305: Pathology examination of tissue using a microscope, intermediate complexity), which determines reimbursement^19^. There were also some instances where no primary code was assigned. Classification models were configured with these seven scenarios in mind (six CPT codes, no CPT code). Forty-one deep-learning classification models were compared for this task from the General, Medical and Unrelated pretraining strategies. Model architectures included but were not limited to: 1) BERT^29^, 2) DistilBERT^30^, 3) RoBERTa^22^, 4) XLNet^31^, and 5) BigBird^32^. Each model outputs a predicted probability or “confidence score” for each class (see Supplementary Table S1). Performance was measured using the accuracy, balanced accuracy score, F1 score, precision, recall, and AUC per CPT code. All models use the same training, validation, and testing sets.

#### Task #2, Pathologist classification

The pathologist classification task is similar to the CPT code classification task. Pathologists’ labels in the dataset are unbalanced. To account for this, only data from six pathologists with the highest number of reports were used, with a total of 146,334 reports included in the final analysis, representing a subset of the CPT code data. We identified the top six pathologists and labeled the pathologists from # 1 to # 6, who wrote the most diagnosis reports. Pretraining strategies and model architectures were the same as those used for the CPT code classification task with similar performance metrics, and the same training, validation, and testing sets were employed across different models.

#### Task #3, Sign-out time regression

Sign-out time is calculated from the date when the specimen is received to the date when the sign-out is completed. Usually, a long sign-out time indicates a complicated case or a case with incomplete information. By training models to predict the sign-out time length of the report, we wanted to test the model’s ability to understand case complexity and detect incomplete information, a task that requires deeper insight compared to understanding just report content. The continuous label—in this case the sign-out time of each report—is calculated based on the difference between the received date and the sign-out date, excluding weekends. We modeled the number of days for sign-out using the negative log-likelihood of the Poisson distribution. Evaluation metrics included explained variance score, mean squared error, mean absolute error, median absolute error, r2 score, and mean Poisson deviance.

#### Task #4, Report text generation

We evaluated the models’ proficiency in suggesting/generating diagnostic text, anticipating that these suggestions might expedite the sign-out process. However, a detailed study of this remains beyond the current scope. For model training only, individual reports were truncated to a length of 128 tokens (approximately 64 words). Text predictions were made by providing a certain proportion of original text, using proportions in increments of 0.1, ranging from 0.1 to 0.9, also including 0.25 and 0.75, see Supplementary Table S4, and predicting the text for the next 3-5 tokens to function as an autocomplete suggestion tool for diagnostic report writing. For instance, a proportion of 0.1 would mean that only the beginning 10% of the original text would be used as input to the model to predict the full text. For obvious reasons, we would expect a larger seed text proportion to facilitate the highest accuracy predictions. For each seed report text, we determine the 5 predicted text sequences with the highest cumulative probabilities using beam search. Beam search is a widely used search algorithm in natural language processing and machine translation. It maintains a fixed number of candidate sequences called the “beam width.” At each step, it considers all possible extensions of the current candidates and selects the top-K sequences with the highest scores. This process continues until a stopping criterion is met. Beam search prunes less-promising candidates to improve efficiency^33^. We used two different evaluation procedures to measure the performance of all 5 predictions and took the average to be the performance of the model on the report generation task. The first method was the accuracy score^34^. Accuracy was defined as the proportion of the next prediction’s tokens correctly predicted. However, this approach does not account for synonyms, motivating our second evaluation method— cosine similarity. We applied the *torch.nn.CosineSimilarity* function from the Torch Neural Networks (torch.nn) module. This computes the cosine similarity between two embedding vectors, as in similarity 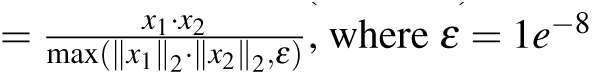. Word embedding cosine similarity score is an assessment of the semantic similarity between the two reports. Here, we used 8 models in total, including GPT2/3^35^, OPT^36^, and BERT^29^. We also compared GPT models by their architecture size (small, medium, large), anticipating that the larger architectures, with their enhanced model capacity, would more effectively discern subtle lexical patterns.

### Model Architectures

Among the four tasks featured in this article, three neural network architectures were used most frequently, including the Bidirectional Encoder Representations from Transformers (BERT), Generalized Autoregressive Pretraining Model (XLNet), and Generative Pretrained Transformer (GPT). Brief descriptions of these architectures have been included below.

#### Model #1, Bidirectional Encoder Representations from Transformers (BERT)

BERT^29^ is a type of neural network which leverages transformers that consist of several self-attention layers^37^. The first step in capturing key semantic, syntactic, and contextual information involves mapping each word to an embedding vector, along with a positional encoding that accounts for the word order in a sentence. These word-level embeddings are then transferred to self-attention layers, which contextualize the information of a single word in a sentence based on both short- and long-term dependencies between all the words in the sentence. The final classification is achieved through a sequence of fully connected layers.

#### Model #2, Generalized Autoregressive Pretraining Model (XLNet)

XLNet is a generalized autoregressive pretrainingmethod that leverages the hidden states of previous segments, differing from BERT’s autoencoding methodology. Unlike BERT, which is initialized by removing and recapitulating significant portions of the text (Masked Language Modeling– MLM), XLNet attempts to consider and predict all possible and plausible permutations (i.e., rearrangements) of the text (Permutation Language Modeling– PLM). Prior research has demonstrated the superior performance of XLNet 20 tasks, often by a large margin, under comparable experiment settings.^31^

#### Model #3, Generative pretrained Transformer (GPT)

Many versions of GPT have gained widespread repute, such as GPT-2, GPT-3, GPT-3.5, and GPT-4. This article will discuss the usage of GPT-2, a large Transformer with 1.5 billion parameters. At the time that this research was conducted, GPT-2 was the only open-source community model supported at the time analysis was conducted. By pretraining on a large, diverse corpus with long stretches of contiguous text, GPT acquires the capability to process long-range dependencies, which can be successfully leveraged to solve discriminative tasks such as question answering, semantic similarity assessment, entailment determination, and text classification, improving the state of the art on 9 of the 12 datasets.^35,21^

### Pretraining Comparison

#### Fine-tuning pretrained model

All models were fine-tuned using the Huggingface python library^38^. During model training, all layers are unfrozen (i.e., parameters were not fixed) based on pretraining via MLM/PLM on select corpora (see supplementary Table S6) as we sought to compare the ability of these layers to extract textual features, all layers were finetuned (i.e., further optimized at a lower learning rate) (Figure 1)^39^. The corpora considered for initial training of these models were broadly characterized by whether they were developed based on a General text corpus, aligned with a Medical text corpus, or comprised of Unrelated text corpora entirely (see supplementary Table S6). Several hyper-parameters were compared using the validation set and selected based on performance on the validation set (see Supplementary Table S5, which includes # of steps, learning rate, etc). Predictive performance on the test set was compared between all architectures and pretraining approaches for the four tasks to identify any salient patterns.

## Results

### CPT Code Classification

Overall, all deep learning models exhibited exemplary performance in assigning primary CPT codes (Average Macro-AUC=0.979; Table 1, Supplementary Figure S1). Of the model types assessed, models initialized on a medical-related corpus performed the best. The other two model types performed similarly, which conformed to our expectations, given the exposure these models had to a diverse array of medical terminology.

**Table 1.** CPT Code Classification results and metrics for all models can be found in Table 1. (25%, 50%, 75% are quantiles of performance among all tested models.)

| Model Type | Mean±SE | 25% | 50% | 75% | # of Models |
| --- | --- | --- | --- | --- | --- |
| **Weighted F1** |  |  |  |  |  |
| General | 0.920±0.001 | 0.919 | 0.920 | 0.920 | 11 |
| Medical | 0.922±0.001 | 0.921 | 0.922 | 0.922 | 26 |
| Unrelated | 0.920±0.001 | 0.920 | 0.920 | 0.921 | 5 |
| **Weighted AUC** |  |  |  |  |  |
| General | 0.954±0.013 | 0.945 | 0.952 | 0.963 | 11 |
| Medical | 0.957±0.017 | 0.943 | 0.957 | 0.974 | 26 |
| Unrelated | 0.971±0.010 | 0.973 | 0.974 | 0.975 | 5 |
| **Macro AUC** |  |  |  |  |  |
| General | 0.976±0.008 | 0.972 | 0.976 | 0.979 | 11 |
| Medical | 0.976±0.009 | 0.972 | 0.975 | 0.984 | 26 |
| Unrelated | 0.984±0.005 | 0.982 | 0.985 | 0.986 | 5 |

### Pathologist Classification

All three pretraining approaches showed great performance in classifying the practicing pathologist. The average AUC scores were 0.993, 0.991, and 0.995 for pretrained models, fine-tuned models, and other models, respectively. However, models pretrained on large, general corpora tended to outperform models initialized on medical corpora (as shown in Supplementary Figure S2, Table 2), possibly because most medical text follows a standardized nature that does not possess many variations in writing patterns or lexicon as would be expected when considering the individuals who populate these reports. The general pretrained models, on the other hand, are better able to uncover the features of the text itself because it has already been trained on a large number of common texts.

**Table 2.** Pathologists Classification Results, and metrics for all models can be found in Table 2. (25%, 50%, 75% are quantiles of performance among all tested models.)

| Model Type | Mean±SE | 25% | 50% | 75% | # of Models |
| --- | --- | --- | --- | --- | --- |
| **Weighted F1** |  |  |  |  |  |
| General | 0.922±0.002 | 0.920 | 0.923 | 0.923 | 9 |
| Medical | 0.921±0.001 | 0.920 | 0.921 | 0.922 | 24 |
| Unrelated | 0.921±0.001 | 0.920 | 0.920 | 0.921 | 5 |
| **Weighted AUC** |  |  |  |  |  |
| General | 0.992±0.001 | 0.991 | 0.992 | 0.992 | 9 |
| Medical | 0.992±0.001 | 0.992 | 0.992 | 0.993 | 24 |
| Unrelated | 0.992±0.001 | 0.991 | 0.992 | 0.993 | 5 |
| **Macro AUC** |  |  |  |  |  |
| General | 0.990±0.001 | 0.990 | 0.990 | 0.991 | 9 |
| Medical | 0.991±0.001 | 0.991 | 0.991 | 0.992 | 24 |
| Unrelated | 0.990±0.001 | 0.989 | 0.991 | 0.991 | 5 |

### Sign-out time regression

As can be seen by Table 3 and Supplementary Figure S3, we used three main performance metrics: explained variance score, mean squared error and R2 score. Figure 2 illustrates that the predicted sign-out time closely aligns with the actual sign-out time across various degrees of domain relevance in the pretraining corpora. Overall, models pretrained on either medical or unrelated corpora exhibited comparable performance for this task. A small number of models pretrained on general corpora exhibited anomalously poor performance, adversely affecting the results.

**Table 3.** Sign-out time regression results and metrics for all models can be found in Table 3. (25%, 50%, 75% are quantiles of performance among all tested models.)

| Model Type | Mean±SE | 25% | 50% | 75% | # of Models |
| --- | --- | --- | --- | --- | --- |
| **Explained Variance** |  |  |  |  |  |
| General | 0.327±0.240 | 0.008 | 0.484 | 0.498 | 12 |
| Medical | 0.469±0.094 | 0.478 | 0.489 | 0.496 | 28 |
| Unrelated | 0.477±0.018 | 0.453 | 0.457 | 0.477 | 5 |
| **Mean Squared Error** |  |  |  |  |  |
| General | 6.676±2.636 | 4.900 | 5.029 | 9.915 | 12 |
| Medical | 5.198±1.000 | 4.916 | 4.981 | 5.087 | 28 |
| Unrelated | 5.211±0.173 | 5.098 | 5.292 | 5.336 | 5 |
| **Mean Absolute Error** |  |  |  |  |  |
| General | 1.336±0.472 | 1.001 | 1.014 | 1.893 | 12 |
| Medical | 1.046±0.184 | 0.997 | 1.004 | 1.010 | 28 |
| Unrelated | 1.014±0.017 | 1.005 | 1.007 | 1.008 | 5 |
| **R2 Score** |  |  |  |  |  |
| General | 0.306±0.270 | -0.017 | 0.484 | 0.497 | 12 |
| Medical | 0.467±0.103 | 0.478 | 0.489 | 0.496 | 28 |
| Unrelated | 0.466±0.018 | 0.453 | 0.457 | 0.477 | 5 |

**Figure 2.**
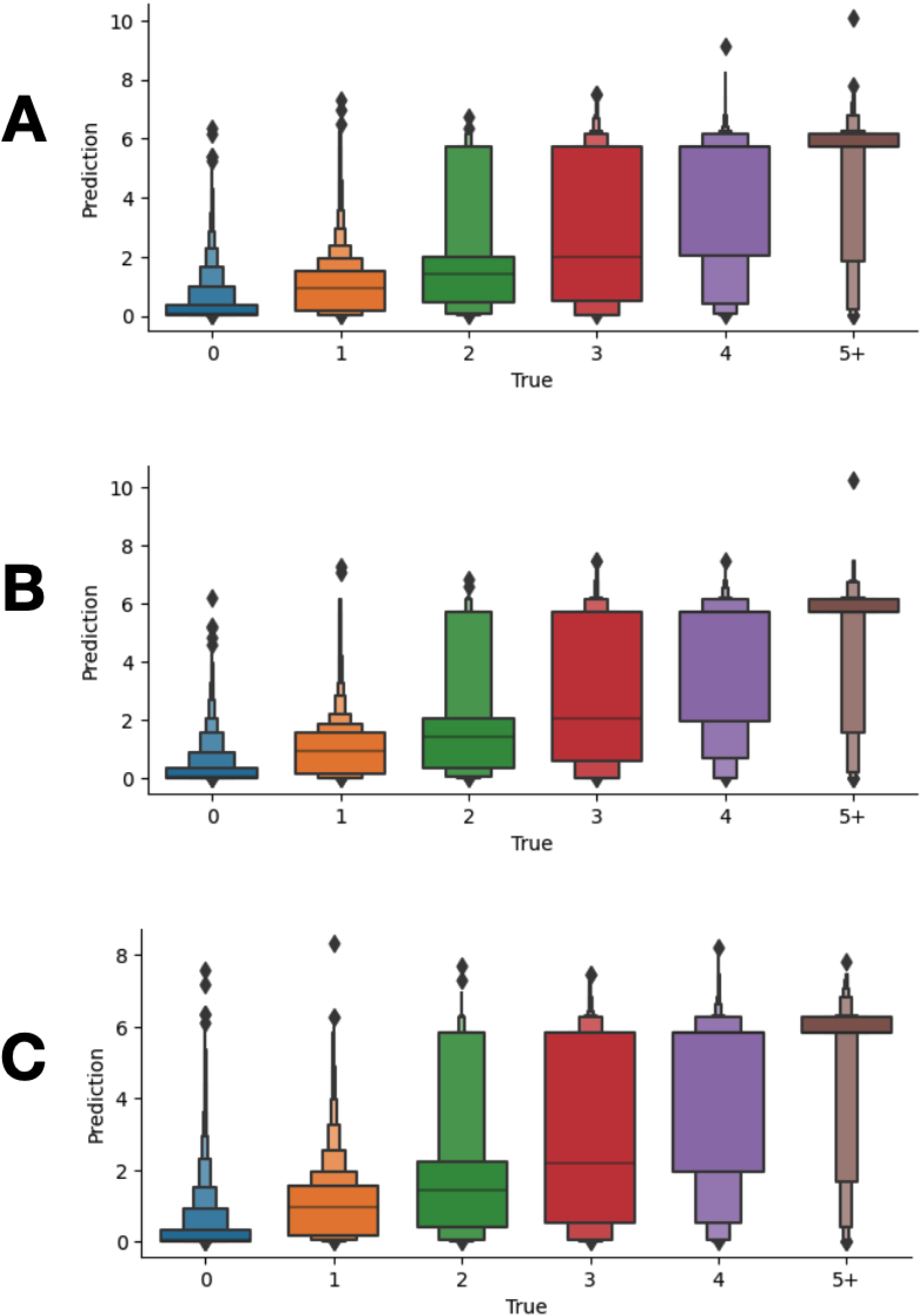
Boxenplots for sign-out time. The X-axis tracks True labels, and the Y-axis measures predictions. Outliers have been excluded. (A) Best performance model in General models, which is bert-base-uncased (B) Best performance model in Medical models, which is bionlp/bluebert_pubmed_uncased_L-12_H-768_A-12 (C) Best performance model in Unrelated models, which is bhadresh-savani/distilbert-base-uncased-emotion

### Text Generation

Figure 3 and Supplementary Figure S4 record the accuracy of next token prediction based on the amount of previous (i.e., seed) text provided, up to a certain percentage of the overall report. As expected, it was easier to predict the next 3 tokens as compared to the next 5 tokens. Notably, all models performed better as the amount of seed text provided increased. Model performance varied based on the number of model parameters (small, medium, large model architectures). The medium-sized model and large-sized GPT-2 models achieved the highest accuracy in both the prediction of the next 3 and 5 tokens. The medium-sized model outperformed the other approaches regarding the semantic similarity of the generated versus original text as assessed using the word embedding cosine similarity score. The GPT-2 model that was trained on the PubMed corpus performed significantly worse than the other models. Models can also be further trained with more training iterations. Results are shown in Supplementary Figure S4 and hyperparameters at Supplementary Table S5.

**Figure 3.**
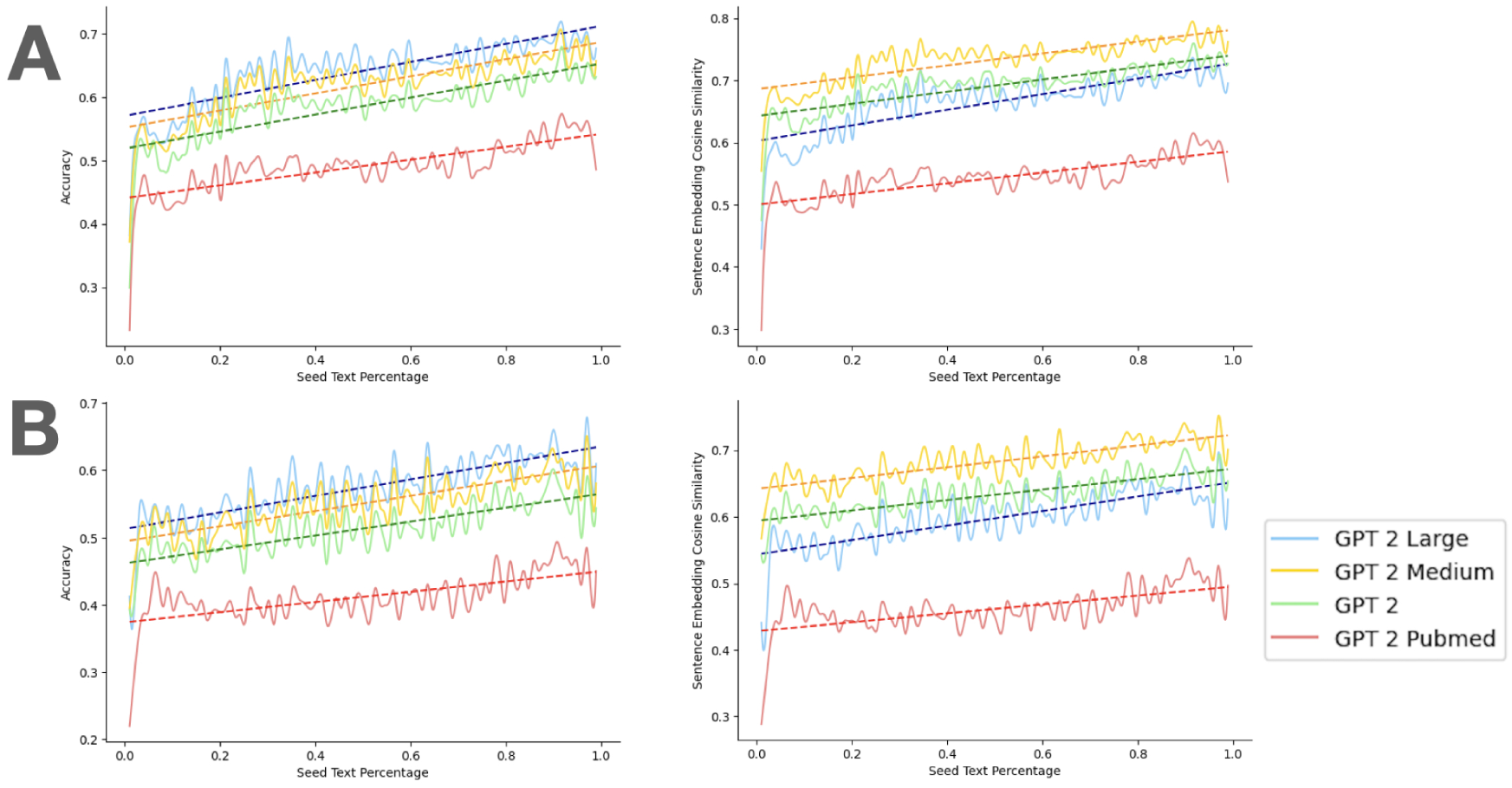
Text generation results with same training iterations: A. Predict next 3 tokens. Accuracy score (left), word embedding cosine similarity (right). B. Predict next 5 tokens. Accuracy score (left), word embedding cosine similarity (right)

## Discussion

The aim of this study was to compare various NLP models in the context of pathological text tasks to address the increasing relevance of NLP in the healthcare domain. Existing literature emphasizes the significance of pretraining strategies. This research compares the performance of large-scale pretrained language models on pathology reports across multiple tasks. Furthermore, it delves into the importance of pretraining language models in the medical and healthcare domain, specifically for pathological text analysis.

### Classification and regression task

Based on our experimental findings, pretraining models using medically related text is a better option for tasks that require extraction and a literal interpretation of pathology or medicine concepts, such as assigning CPT codes. This may, however, reflect the design of the experiment and the limited set of models assessed. For example, we would expect sign-out time and CPT code usage to partially reflect case complexity and uncertainty^40^, where more challenging cases could impact the measurement of relative value units (RVUs). However, sign-out time data is only a one-sided representation of the complexity of the text and may not represent true complexity in several cases. For instance, case sign out time is expected to vary by diagnostic subspecialty but could also reflect a pathologists training/experience with a specific task– communication of medically relevant jargon may reflect these expertise. Surprisingly, fine-tuning models using medical text did not provide a significant advantage in predicting sign-out time. This would suggest that there are textual elements existing beyond pathologist expertise as reflected with familiarity with medical jargon. For the pathologist prediction task, we found that capturing a pathologist-specific lexicon may also reflect subtle differences in subspecialty, a general text corpus is a more suitable choice.

### Text generation task

We also noticed that GPT models did not perform as well when pretrained on the PubMed dataset. To ensure precise word-for-word generation, larger models for the pathology text generation task may incorporate a more extensive collection of information registries. Pathologists who use text generation models are primarily interested in generating text that is semantically similar, rather than focusing on word-for-word accuracy. This is particularly true in cases where exact precision is not necessary. Depending on the situation, the advantage from leveraging large models is not necessarily significant. Based on the experimental results, it was observed that increasing the amount of information provided to the text generation model improves its performance. This finding can be useful in improving the efficiency of writing pathology reports. Further research is required to determine the optimal amount of text that should be written/generated to minimize the time taken to remedy errors during report correction. For example, reducing the number of predicted tokens from five to three will result in a performance improvement at the expense of having to query the model more often to generate additional text. Overall, it was easier to predict the next three tokens as compared to the next five tokens. This can also be interpreted as the prediction of the near future is better than the prediction of the longer term future after the same information provided, i.e., given the seed text. Recent advancements in text generation models have led to the development of large models with increased number of parameters. Based on the accuracies reported for different models in the text generation task, we also find that the size and complexity of the model play a decisive role in model performance, consistent with broader trends in the academic literature^41^.

### Domain adaptation

In the context of domain adaptation, it is generally believed that aligning the source and target domains is crucial for minimizing data requirements and improving models. However, in pathology reporting, there are several complexities that may undermine the validity of this assumption. Although a pretraining corpus may include medical text, there is no guarantee that the resulting model will perform better for a specific reporting style. This is partly because pathologist reporting often involves subjective language that reflects a detailed and nuanced consideration/interpretation of the patient’s case, with multiple perspectives and ambiguous data sources that hedge against potential diagnoses. Moving forward, it is important to note that the performance of medical text fine-tuning models is influenced by the anticipated similarity of pathology reporting text to the source domain in the context of the experimental task, the size of the model, model complexity, and the number of parameters. Based on our findings, we observed that the performance of the model reaches a saturation point with larger architectures. Additionally, the model’s performance is influenced by the similarity between the text from the source domain and the target domain, which includes not only terminology but also lexicon, verbiage, and syntax. All attribution tables can be found in Supplementary Table S12 - S47. This is particularly evident in domains such as pathology reports. Larger base models which have been initialized on generalized corpora tend to outperform smaller, more specialized models with limited training resources, number of training iterations, and amount of data for fine-tuning.

### Limitations and Future Directions

In order to be able to compare the performance of different models under similar experimental conditions, similar hyperparameters were leveraged for each task, which may have resulted in model under/over-fitting. As there were widely different model architectures being compared, we attempted only to optimize the final layers of each neural network, which does not guarantee optimal convergence. To accommodate GPU memory limitations, gradient accumulation was utilized to ensure consistent batch sizes.

We have chosen popularly utilized pretrained models for our study from an ever-expanding collection of corpora. Our selection criteria primarily focused on medically relevant pretrained models, as our goal is to compare their performance across various NLP tasks in Pathology. Yet it is possible that several nascent approaches that may have been more relevant to the target domain may have been omitted^42,43,13^. Although our paper briefly mentions other pretrained models, they are not the primary focus of our experiments but rather serve as a negative control. We found that in certain instances, depending solely on highly specialized medical corpora actually hindered our results. We expect that results may vary by institution, necessitating additional validation.

While modeling approaches have the potential to detect lexical patterns that capture reporting variations and reasons for potential sign-out delays in pathology reports, these reports still lack a structured format. Thus, it is essential to extract valuable clinical information and knowledge from them to establish databases for clinical studies and facilitate rapid querying in clinical practice. For instance, the identification of staining results and relevant diagnostic information can populate specific fields in the database, expediting follow-up examinations. Moreover, pathology reporting often involves uncertainty and hedging against specific diagnoses, reflecting the reflexive ordering of stains and individual pathologists’ comfort level, proficiency, expertise, and practical knowledge along with true case complexity. Future investigations will focus on leveraging deep learning algorithms to extract clinical information and address instances of uncertainty, transforming them into a highly structured format. Extraction of this information could facilitate longitudinal assessments for large-scale epidemiological studies and quality improvement practices.

## Conclusions

Prior studies have indicated that constructing deep learning models for natural language processing (NLP) by incorporating information from a related research and practice domain can improve their performance across different tasks. Our investigation aimed to examine this assumption in the field of Pathology across multiple tasks. Surprisingly, our findings contradicted these expectations, demonstrating that in specific instances, utilizing information from a medically-related domain rather than more general or unrelated corpora may not lead to a significant enhancement in performance. More generally, we conclude that model pertaining strategies are highly contingent upon the specific clinical context being examined. These findings can provide valuable insights when choosing deep learning models for tasks related to Pathology, serving as guiding principles in the selection process.

## Data Availability

Access to manuscript data is limited due to patient privacy concerns. All data produced in the present study are available upon reasonable request to the authors.

## Supplementary Materials

**Table S1.** CPT Code Classification example with diagnostic text, actual and predicted CPT code, and confidence level. For the first case the model is very confident and prediction is correct. For the second case, the model is not as confident, and predictions are incorrect

| Pathology Diagnosis Text | Actual Code | Predict Code | Confidence Score |
| --- | --- | --- | --- |
| skin left mid back shave removal lentiginous compound dysplastic nevus with moderate atypia irritated the margins appear clear in these sections | 88305 | 88305 | 0.974 |
| skin left hip excision no residual of the previously diagnosed compound melanocytic nevus with atypical features reparative changes consistent with previous operative site see comment | 88305 | 88302 | 0.586 |

**Table S2.** Pathologists Classification example with diagnostic text, actual and predicted pathologist, and confidence level. The model is confident about both of these two cases. The first example is correct prediction and the second one is an incorrect prediction, are provided

| Pathology Diagnosis Text | Actual | Predict | Confidence Score |
| --- | --- | --- | --- |
| A - Sigmoid colon, polypectomy: Two tubular adenomas, completely excised. B - Ascending colon, polypectomy: Colonic mucosa, negative for diagnostic abnormality. Submucosal adipose tissue, that may be compatible with submucosal lipoma in an appropriate endoscopic setting. CR-0, CR-PX | # 4 | # 4 | 0.999 |
| A - Esophagus at 20 to 30 cm, biopsy: Esophageal squamous mucosa with focal basal hyperplasia. Dilatation of mucosal capillaries. B - Esophagus at 15 to 20 cm, biopsy: Esophageal squamous mucosa with focal dilatation of mucosal capillaries. CR-0 | # 4 | # 3 | 0.992 |

**Table S3.** Sign-Out Time Regression example with diagnostic text, actual and predicted signout time length. For both of the two cases, the model predicts close to accurate outcomes.

| Pathology Diagnosis Text | Actual Time Length | Predict Time Length |
| --- | --- | --- |
| skin mid central back shave removal intradermal melanocytic nevus with features of congenital onset focally involving biopsy edges | 6 | 5.953 |
| skin right lower leg shave biopsy epidermal hyperplasia with lichenoid inflammation see comment | 1 | 1.112 |

**Table S4.** Text Generation example with diagnostic text, original text proportion.

| Pathology Diagnosis Text | Seed Text Proportion |
| --- | --- |
| atosis ulcerated cr suspicious for malignancy compound dysplastic melanocytic nevus with mild to moderate atypia and features of congenital onset extending to peripheral specimen edge(s a skin left upper outer arm shave biopsy lentiginous compound dysplastic nevus with moderate atypia extending close to the deep specimen edge along adnexal s tructure see comment b skin middle lumbar spine shave biopsy lentiginous compound dysplastic nevus with moderate atypia extending close to the deep specimen edge along adnexal s tructure see comment cr skin right upper | Original Text |
| atosis ulcerated cr suspicious for malignancy compound dysplastic melanocytic nevus with mild to moderate atypia and features of congenital onset extending to peripheral specimen edge(s a skin left upper outer arm shave biopsy lentiginous compound dysplastic nevus with moderate atypia **biopsy edges appear narrowly negative in the plane of sections examined see discussion b skin right upper outer arm shave biopsy lentiginous junctional melanocytic nevus specimen edges not involved on the plane of section examined multiple deeper levels have been examined cr normal negative for intraepithelial lesion or malign** | 50% |

**Table S5.** Hyperparameters Set.

| Hyperparameter | Assignment |
| --- | --- |
| sequence length for CPT | 256 |
| sequence length for pathologists | 512 |
| CPT code classification update steps | 12,000 (5 epochs) |
| pathologists classification update steps | 18,000 (5 epochs) |
| sign-out time regression update steps | 23,000 (10 epochs) |
| first part text generation update steps | 5,600 (15 epochs) |
| second part text generation GPT2-Medium update steps | 16,800 (45 epochs) |
| second part text generation GPT2-Large update steps | 33,600 (90 epochs) |
| batch size for others | 32 |
| batch size for text generation | 16 |
| maximum learning rate (classification) | 1e-5 |
| maximum learning rate (regression) | 5e-5 |
| maximum learning rate (text generation) | 2e-5 |
| learning rate optimizer | Adam Optimizer |
| Adam epsilon | 1e-8 |
| Adam beta weights | 0.9, 0.999 |
| learning rate scheduler | AdamW |
| Weight decay | 0.01 |
| fp16 | True |

**Table S6.** Datasets used for pretraining models and their descriptions.

| Datasets | Description |
| --- | --- |
| **General** |  |
| English Wikipedia Corpus | This dataset comprises over 4.4 million articles, containing a total of 1.9 billion words. It includes various types of text passages, such as lists, tables, and headers. |
| BooksCorpus<sup>44</sup> | The BooksCorpus is a collection of 11,038 books sourced from the web. These books are written by unpublished authors and are available for free. To ensure quality, only books with a minimum word count of 20,000 were included, filtering out shorter and potentially noisier stories. |
| Open WebText<sup>45</sup> | The Open WebText dataset is gathered from the online platform Reddit. Preprocessing techniques are applied to remove similar, very short, and non-English content. It consists of over 8 million documents, making it a substantial resource for language modeling. |
| HotpotQA<sup>46</sup> | HotpotQA is a relatively new dataset containing 113,000 question-answer pairs that are based on information from Wikipedia. It is specifically designed to challenge question-answering models. |
| CommonCrawl News EN<sup>47,2248</sup> | This dataset comprises 44 million English documents collected between September 2016 and March 2018. It serves as a valuable resource for training language models with real-world text data. |
| **Medical** |  |
| n2c2 dataset | The n2c2 dataset is a collection of datasets used for various NLP challenges. It spans from the year 2006 to 2019 and contains diverse NLP data related to the medical domain. |
| An Evaluation Dataset for Intent Classification and Out-of-Scope Prediction<sup>49</sup> | This dataset is a crowdsourced collection of 23,700 queries, including 22,500 in-scope queries covering 150 intents grouped into 10 general domains. Additionally, it includes 1,200 out-of-scope queries, providing a comprehensive benchmark for intent classification and out-of-scope prediction tasks. |
| MIMIC-III dataset<sup>50</sup> | The MIMIC-III dataset is a large medical database that encompasses a wide range of information. It includes vital signs, medications, laboratory measurements, observations and notes charted by care providers, fluid balance, procedure codes, diagnostic codes, imaging reports, hospital length of stay, survival data, and more. It serves as a valuable resource for various medical research and analysis tasks. |
| WikiSection Dataset<sup>51</sup> | The WikiSection Dataset consists of 38,000 full-text documents from English and German Wikipedia that are comprehensively annotated with sections and topic labels. The dataset focuses on medical topics and covers up to 30 different topics related to diseases (e.g., symptoms, treatments, diagnosis) or cities (e.g., history, politics, economy, climate). It provides a rich resource for studying specific medical domains within Wikipedia articles. |
| PubMedSection dataset<sup>51,52</sup> | The PubMedSection dataset is a topic classification dataset based on medical research articles from PubMed. It comprises 51,500 articles that are annotated section-wise with topic labels. This dataset is particularly useful for training and evaluating models focused on medical text classification tasks. |
| MedQuAD: Medical Question Answering Dataset<sup>53</sup> | The MedQuAD dataset consists of 47,457 question-answer pairs sourced from 12 NIH websites. It covers a wide range of medical entities, including diseases, drugs, and tests, and includes 37 question types (e.g., Treatment, Diagnosis, Side Effects). This dataset serves as a valuable resource for training and evaluating medical question-answering systems. |
| **Unrelated** |  |
| Bloomberg News<sup>54</sup> | The Bloomberg News dataset comprises 400,000 financial articles published by Bloomberg between 2006 and 2013. It provides a valuable resource for training language models focused on the financial domain. |
| TRC2-financial<sup>54</sup> | Thomson Reuters Text Research Collection (TRC2) corpus– The TRC2-financial dataset consists of 1,800,370 news stories published by Reuters, covering the period from 2008 to 2010. It serves as a comprehensive collection of financial news, enabling the development of models tailored to the financial domain. |
| Corporate Reports | The Corporate Reports dataset is sourced from the EDGAR database, where publicly traded companies are required to file annual reports (10-K) and quarterly reports (10-Q) to the Securities Exchange Commission (SEC). It contains 154,354 documents of 10-K reports from 1996 to 2015 and 37,646 quarterly reports (10-Q). This dataset is filtered based on relevant keywords, making it suitable for research and analysis of corporate financial information. |

**Figure S1.**
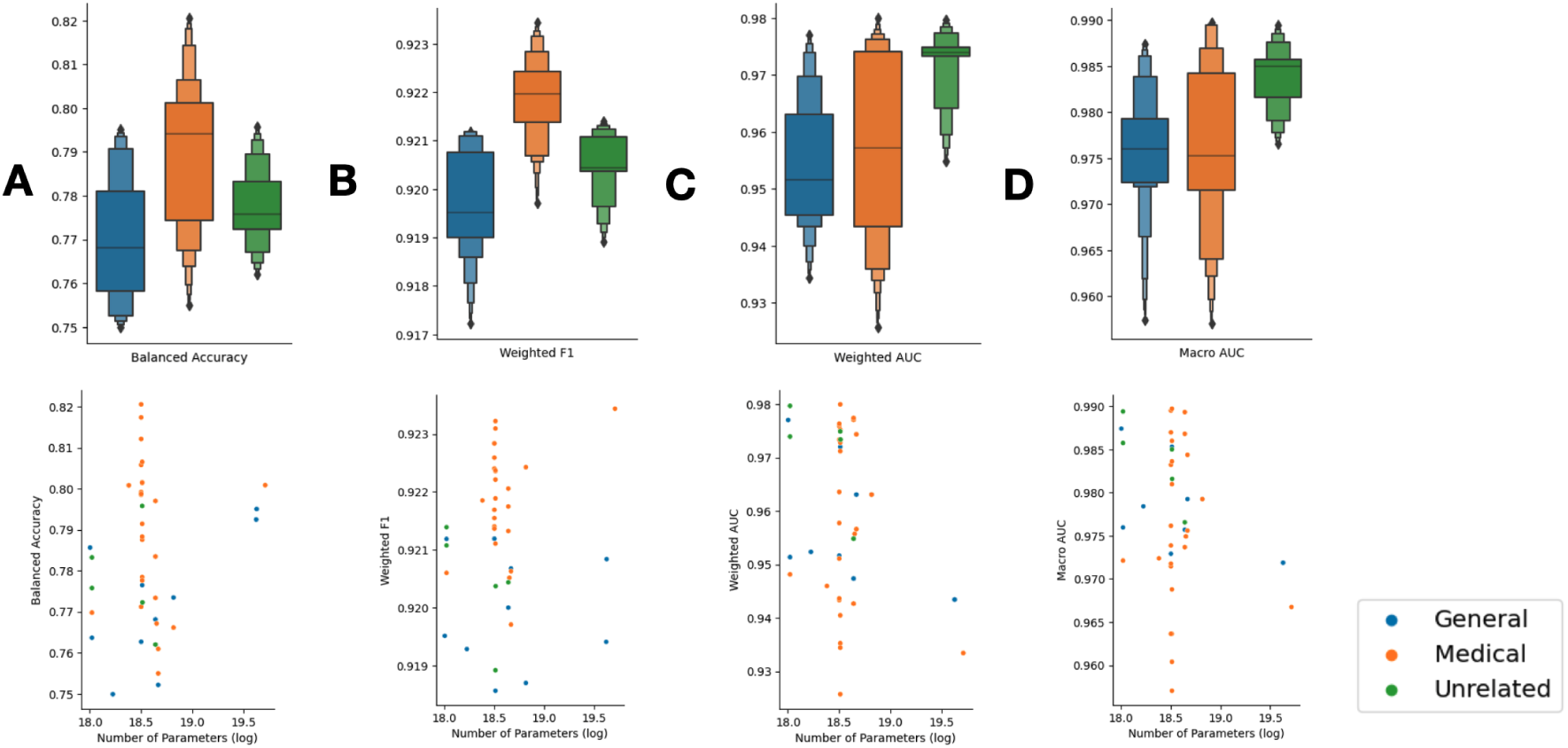
CPT Code Classification: A. Box plot for balanced accuracy score (up), scatter plot for balanced accuracy score and number of model parameters (down), B. Box plot for weighted F1 score (up), scatter plot for weighted F1 score and number of model parameters (down), C. Box plot for Weighted AUC (up), scatter plot for weighted AUC and number of model parameters (down), D. Box plot for Macro AUC (up), scatter plot for weighted AUC and number of model parameters (down) (Without models with number of parameters smaller than 1*e*^17^)

**Figure S2.**
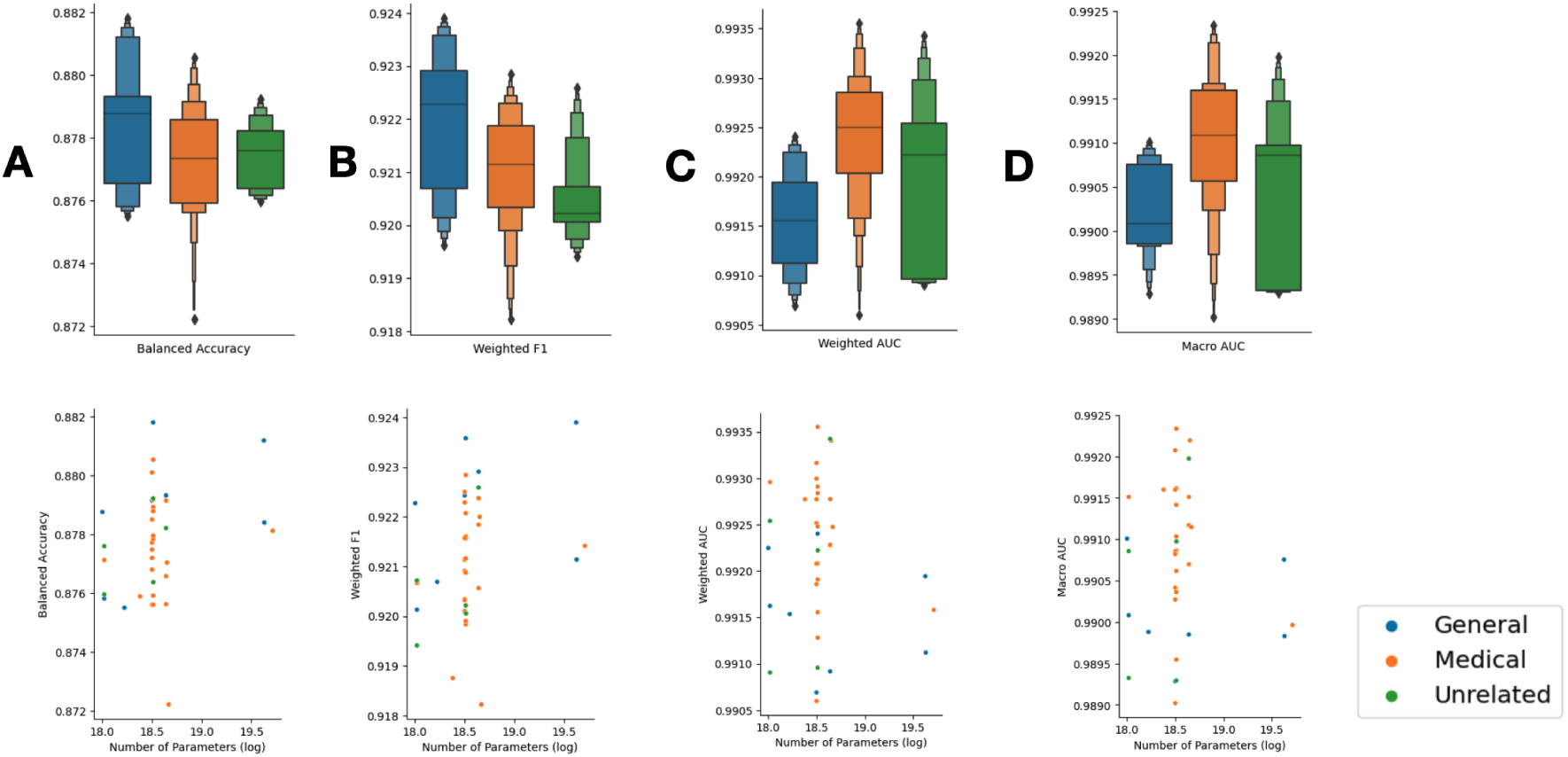
Pathologists Classification: A. Box plot for balanced accuracy score (up), scatter plot for balanced accuracy score and number of model parameters (down), B. Box plot for weighted F1 score (up), scatter plot for weighted F1 score and number of model parameters (down), C. Box plot for Weighted AUC (up), scatter plot for weighted AUC and number of model parameters (down), D. Box plot for Macro AUC (up), scatter plot for weighted AUC and number of model parameters (down) (Without models with number of parameters smaller than 1*e*^17^)

**Figure S3.**
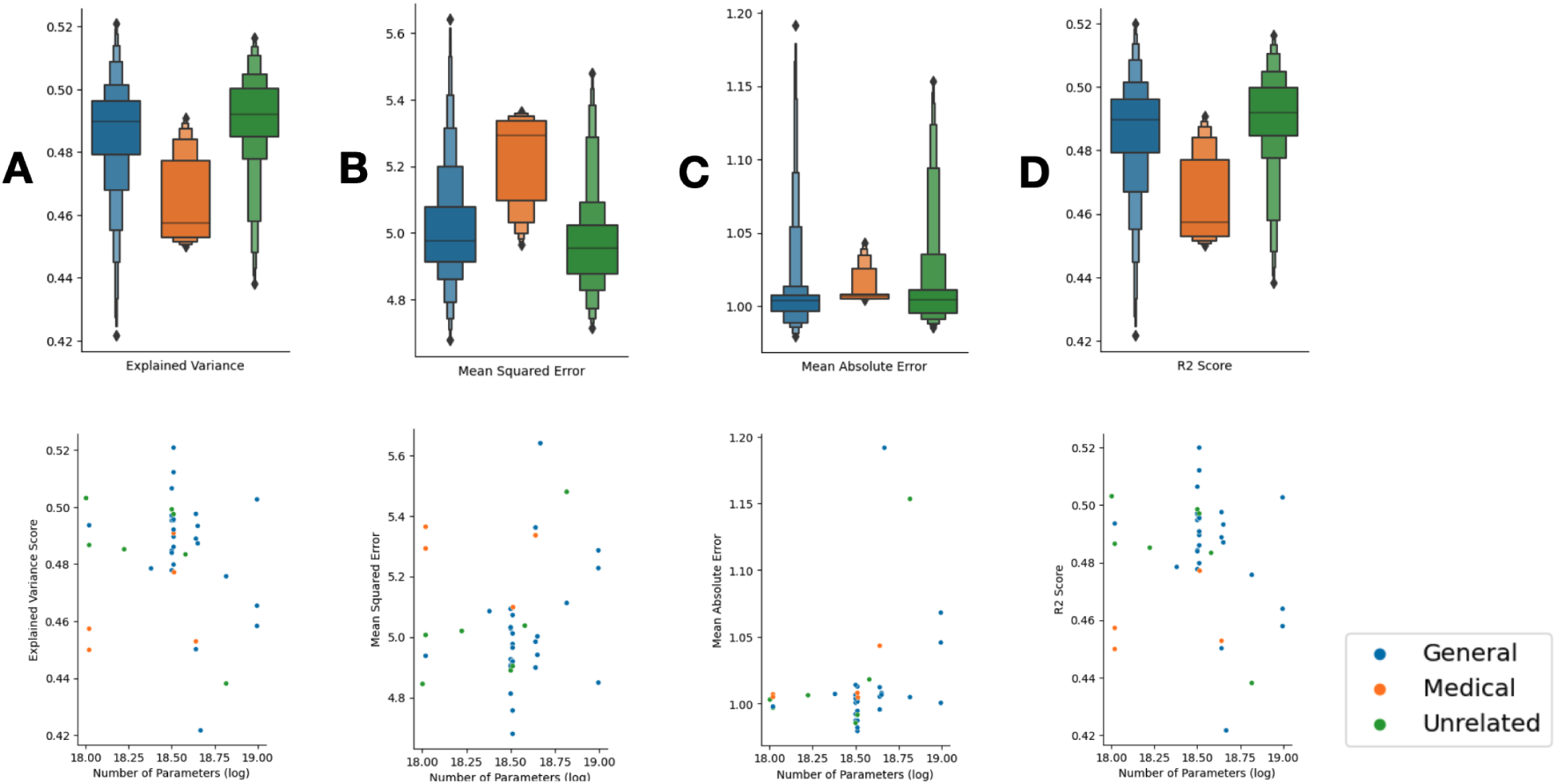
Sign-out Time Regression (Outliers removed): A. Box plot for explained variance score (up), scatter plot for explained variance score and number of model parameters (down), B. Box plot for mean squared error (up), scatter plot for mean squared error and number of model parameters (down), C. Box plot for mean absolute error (up), scatter plot for mean squared error and number of model parameters (down), D. Box plot for R2 score (up), scatter plot for R2 score and number of model parameters (down) (Without models with number of parameters smaller than 1*e*^17^)

**Table S7.** CPT Code Classification Results Across All LLMs, broken down by domain relevance of pretraining corpora.

| Model Name | Loss | Accuracy | F1 | Precision | Recall | Weighted AUC | Macro AUC | Runtime |
| --- | --- | --- | --- | --- | --- | --- | --- | --- |
| **Bert Base Models** |  |  |  |  |  |  |  |  |
| bert-base-cased | 0.257 | 0.773 | 0.921 | 0.924 | 0.922 | 0.952 | 0.973 | 26.1 |
| bert-base-uncased | 0.247 | 0.765 | 0.921 | 0.924 | 0.922 | 0.972 | 0.985 | 24.4 |
| bert-large-cased | 0.264 | 0.784 | 0.922 | 0.926 | 0.923 | 0.943 | 0.972 | 43.2 |
| bert-large-uncased | 0.257 | 0.795 | 0.921 | 0.925 | 0.922 | 0.943 | 0.972 | 44.3 |
| **ALBert Base Model** |  |  |  |  |  |  |  |  |
| albert-base-v2 | 0.271 | 0.754 | 0.917 | 0.921 | 0.919 | 0.934 | 0.957 | 23.6 |
| **DistilBert Base Model** |  |  |  |  |  |  |  |  |
| distilbert-base-uncased | 0.245 | 0.764 | 0.921 | 0.925 | 0.923 | 0.951 | 0.976 | 9.9 |
| distilbert-base-cased | 0.247 | 0.786 | 0.920 | 0.924 | 0.920 | 0.977 | 0.987 | 9.5 |
| **Distil-RoBERTa Base Model** |  |  |  |  |  |  |  |  |
| distilroberta-base | 0.249 | 0.750 | 0.919 | 0.923 | 0.921 | 0.952 | 0.978 | 10.2 |
| **RoBERTa Base Model** |  |  |  |  |  |  |  |  |
| roberta-base | 0.246 | 0.768 | 0.920 | 0.924 | 0.921 | 0.947 | 0.976 | 23.9 |
| **BigBird Base Model** |  |  |  |  |  |  |  |  |
| bigbird-roberta-base | 0.245 | 0.752 | 0.921 | 0.924 | 0.922 | 0.963 | 0.979 | 26.8 |
| **Longformer base Model** |  |  |  |  |  |  |  |  |
| allenai/longformer-base-4096 | 0.248 | 0.773 | 0.919 | 0.919 | 0.920 | 0.963 | 0.979 | 61.8 |
| **Bert Base Models Finetuned on Medical Corpus** |  |  |  |  |  |  |  |  |
| samrawal/bert-base-uncased_clinical-ner | 0.253 | 0.778 | 0.922 | 0.926 | 0.924 | 0.975 | 0.986 | 25.6 |
| transformersbook/bert-base-uncased-finetuned-clinc | 0.247 | 0.791 | 0.922 | 0.926 | 0.923 | 0.935 | 0.969 | 26.0 |
| emilyalsentzer/Bio_ClinicalBERT | 0.247 | 0.799 | 0.921 | 0.925 | 0.922 | 0.943 | 0.971 | 26.2 |
| bvanaken/CORE-clinical-outcome-biobert-v1 | 0.242 | 0.821 | 0.923 | 0.927 | 0.924 | 0.973 | 0.983 | 25.9 |
| JacopoBandoni/BioBertRelationG | 0.251 | 0.799 | 0.922 | 0.925 | 0.923 | 0.958 | 0.974 | 25.9 |
| ChrisUPM/BioBERT_Re_trained | 0.247 | 0.817 | 0.923 | 0.927 | 0.924 | 0.951 | 0.972 | 26.0 |
| blizrys/biobert-v1.1-finetuned-pubmedqa | 0.243 | 0.812 | 0.923 | 0.926 | 0.923 | 0.964 | 0.976 | 24.9 |
| blizrys/biobert-base-cased-v1.1-finetuned-pubmedqa | 0.248 | 0.799 | 0.921 | 0.926 | 0.922 | 0.976 | 0.987 | 24.4 |
| dms-lab/biobert-v1.1 | 0.246 | 0.806 | 0.922 | 0.926 | 0.923 | 0.976 | 0.990 | 25.1 |
| dms-lab/biobert-large-cased-v1.1-mnli | 0.260 | 0.801 | 0.923 | 0.927 | 0.924 | 0.933 | 0.967 | 44.0 |
| bluebert_pubmed_mimic_uncased | 0.244 | 0.778 | 0.922 | 0.926 | 0.924 | 0.973 | 0.984 | 26.1 |
| bluebert_pubmed_uncased | 0.242 | 0.788 | 0.921 | 0.925 | 0.922 | 0.980 | 0.990 | 26.3 |
| microsoft/BiomedNLP-PubMedBERT-base-uncased-abstract-fulltext | 0.238 | 0.807 | 0.923 | 0.927 | 0.924 | 0.934 | 0.960 | 26.1 |
| microsoft/BiomedNLP-PubMedBERT-base-uncased-abstract | 0.247 | 0.802 | 0.922 | 0.926 | 0.923 | 0.926 | 0.957 | 25.9 |
| microsoft/BiomedNLP-KRISSBERT-PubMed-UMLS-EL | 0.240 | 0.788 | 0.922 | 0.926 | 0.923 | 0.971 | 0.981 | 26.2 |
| ml4pubmed/BiomedNLP-PubMedBERT-base-uncased-abstract-fulltext_pub_section | 0.238 | 0.801 | 0.923 | 0.927 | 0.924 | 0.940 | 0.964 | 26.3 |
| bvanaken/clinical-assertion-negation-bert | 0.247 | 0.771 | 0.922 | 0.925 | 0.923 | 0.944 | 0.964 | 25.7 |
| tsantos/PathologyBERT | 0.249 | 0.801 | 0.922 | 0.925 | 0.923 | 0.946 | 0.972 | 26.4 |
| **DistilBert Base Model Finetuned on Medical Corpus** |  |  |  |  |  |  |  |  |
| distilbert-pubmed-MLM | 0.249 | 0.770 | 0.921 | 0.925 | 0.922 | 0.948 | 0.972 | 9.6 |
| **RoBerta Base Model Finetuned on Medical Corpus** |  |  |  |  |  |  |  |  |
| biomed_roberta_base | 0.244 | 0.783 | 0.922 | 0.926 | 0.923 | 0.943 | 0.974 | 23.7 |
| bio_roberta-base_pubmed | 0.245 | 0.773 | 0.921 | 0.925 | 0.922 | 0.977 | 0.987 | 23.1 |
| roberta-pubmed | 0.241 | 0.797 | 0.922 | 0.926 | 0.923 | 0.977 | 0.989 | 23.8 |
| roberta-base-biomedical-clinical | 0.250 | 0.767 | 0.921 | 0.925 | 0.922 | 0.955 | 0.975 | 23.4 |
| **BigBird Base Model Finetuned on Medical Corpus** |  |  |  |  |  |  |  |  |
| bigbird-base-mimic-mortality | 0.248 | 0.755 | 0.920 | 0.924 | 0.921 | 0.957 | 0.976 | 25.6 |
| bigbird-base-health-fact | 0.245 | 0.761 | 0.921 | 0.925 | 0.922 | 0.974 | 0.984 | 27.0 |
| **Longformer base Model Finetuned on Medical Corpus** |  |  |  |  |  |  |  |  |
| yikuan8/Clinical-Longformer | 0.242 | 0.766 | 0.922 | 0.927 | 0.924 | 0.963 | 0.979 | 66.2 |
| **Bert Base Models Finetuned on Non Related Corpus** |  |  |  |  |  |  |  |  |
| FinancialBERT-Sentiment-Analysis | 0.260 | 0.772 | 0.920 | 0.924 | 0.921 | 0.973 | 0.982 | 26.1 |
| MathBERT | 0.267 | 0.796 | 0.919 | 0.922 | 0.920 | 0.975 | 0.985 | 26.4 |
| **DistilBert Base Model Finetuned on Non Related Corpus** |  |  |  |  |  |  |  |  |
| distilbert-base-uncased-finetuned-sst-2 | 0.245 | 0.783 | 0.921 | 0.925 | 0.923 | 0.974 | 0.986 | 9.7 |
| distilbert-base-uncased-emotion | 0.244 | 0.776 | 0.921 | 0.925 | 0.922 | 0.980 | 0.990 | 9.6 |

**Table S8.** Pathologists Classification Results Across All LLMs, broken down by domain relevance of pretraining corpora.

| ModelName | Loss | Accuracy | F1 | Precision | Recall | Weighted AUC | Macro AUC | Runtime |
| --- | --- | --- | --- | --- | --- | --- | --- | --- |
| **Bert Base Models** |  |  |  |  |  |  |  |  |
| bert-base-cased | 0.206 | 0.879 | 0.923 | 0.927 | 0.924 | 0.991 | 0.989 | 60.0 |
| bert-base-uncased | 0.201 | 0.882 | 0.924 | 0.928 | 0.925 | 0.992 | 0.991 | 62.4 |
| bert-large-cased | 0.215 | 0.881 | 0.924 | 0.928 | 0.925 | 0.992 | 0.991 | 115.0 |
| bert-large-uncased | 0.214 | 0.878 | 0.921 | 0.925 | 0.923 | 0.991 | 0.990 | 115.1 |
| **ALBert Base Model** |  |  |  |  |  |  |  |  |
| albert-base-v2 | 0.217 | 0.877 | 0.920 | 0.923 | 0.921 | 0.992 | 0.990 | 64.6 |
| **DistilBert Base Model** |  |  |  |  |  |  |  |  |
| distilbert-base-uncased | 0.207 | 0.876 | 0.920 | 0.924 | 0.922 | 0.992 | 0.990 | 24.9 |
| distilbert-base-cased | 0.210 | 0.879 | 0.922 | 0.926 | 0.924 | 0.992 | 0.991 | 24.6 |
| **Distil-RoBERTa Base Model** |  |  |  |  |  |  |  |  |
| distilroberta-base | 0.210 | 0.876 | 0.921 | 0.925 | 0.922 | 0.992 | 0.990 | 27.0 |
| **RoBERTa Base Model** |  |  |  |  |  |  |  |  |
| roberta-base | 0.207 | 0.879 | 0.923 | 0.928 | 0.924 | 0.991 | 0.990 | 57.0 |
| **Bert Base Models Finetuned on Medical Corpus** |  |  |  |  |  |  |  |  |
| samrawal/bert-base-uncased_clinical-ner | 0.208 | 0.879 | 0.922 | 0.925 | 0.923 | 0.992 | 0.991 | 61.1 |
| transformersbook/bert-base-uncased-finetuned-clinc | 0.209 | 0.879 | 0.922 | 0.927 | 0.923 | 0.993 | 0.991 | 57.8 |
| emilyalsentzer/Bio_ClinicalBERT | 0.210 | 0.878 | 0.920 | 0.924 | 0.922 | 0.993 | 0.991 | 58.1 |
| bvanaken/CORE-clinical-outcome-biobert-v1 | 0.211 | 0.876 | 0.921 | 0.926 | 0.923 | 0.992 | 0.990 | 57.9 |
| JacopoBandoni/BioBertRelationG | 0.206 | 0.876 | 0.920 | 0.925 | 0.922 | 0.993 | 0.992 | 58.0 |
| ChrisUPM/BioBERT_Re_trained | 0.207 | 0.877 | 0.921 | 0.926 | 0.922 | 0.993 | 0.991 | 58.8 |
| blizrys/biobert-v1.1-finetuned-pubmedqa | 0.207 | 0.877 | 0.922 | 0.926 | 0.923 | 0.991 | 0.989 | 58.8 |
| blizrys/biobert-base-cased-v1.1-finetuned-pubmedqa | 0.215 | 0.880 | 0.922 | 0.927 | 0.924 | 0.993 | 0.992 | 78.4 |
| dms-lab/biobert-large-cased-v1.1-mnli | 0.222 | 0.878 | 0.921 | 0.926 | 0.923 | 0.992 | 0.990 | 117.2 |
| dms-lab/biobert-v1.1 | 0.203 | 0.879 | 0.923 | 0.927 | 0.924 | 0.992 | 0.990 | 57.6 |
| bluebert_pubmed_mimic_uncased | 0.207 | 0.878 | 0.921 | 0.925 | 0.923 | 0.992 | 0.990 | 60.3 |
| bluebert_pubmed_uncased | 0.208 | 0.876 | 0.920 | 0.924 | 0.921 | 0.994 | 0.992 | 59.8 |
| microsoft/BiomedNLP-PubMedBERT-base-uncased-abstract-fulltext | 0.207 | 0.879 | 0.923 | 0.927 | 0.924 | 0.992 | 0.991 | 58.8 |
| microsoft/BiomedNLP-PubMedBERT-base-uncased-abstract | 0.201 | 0.881 | 0.922 | 0.925 | 0.923 | 0.993 | 0.992 | 59.7 |
| microsoft/BiomedNLP-KRISSBERT-PubMed-UMLS-EL | 0.210 | 0.876 | 0.920 | 0.925 | 0.921 | 0.991 | 0.990 | 58.6 |
| ml4pubmed/BiomedNLP-PubMedBERT-base-uncased-abstract-fulltext_pub_section | 0.208 | 0.878 | 0.921 | 0.925 | 0.922 | 0.992 | 0.991 | 58.2 |
| bvanaken/clinical-assertion-negation-bert | 0.209 | 0.877 | 0.920 | 0.924 | 0.921 | 0.993 | 0.992 | 58.4 |
| tsantos/PathologyBERT | 0.216 | 0.876 | 0.919 | 0.923 | 0.920 | 0.993 | 0.992 | 58.1 |
| **DistilBert Base Model Finetuned on Medical Corpus** |  |  |  |  |  |  |  |  |
| Gaborandi/distilbert-pubmed-MLM | 0.206 | 0.877 | 0.921 | 0.925 | 0.922 | 0.993 | 0.992 | 25.9 |
| **RoBERTa Base Model Finetuned on Medical Corpus** |  |  |  |  |  |  |  |  |
| allenai/biomed_roberta_base | 0.205 | 0.877 | 0.922 | 0.928 | 0.924 | 0.993 | 0.992 | 57.8 |
| minhpqn/bio_roberta-base_pubmed | 0.209 | 0.876 | 0.921 | 0.926 | 0.922 | 0.992 | 0.991 | 56.7 |
| raynardj/roberta-pubmed | 0.211 | 0.879 | 0.922 | 0.926 | 0.923 | 0.992 | 0.991 | 57.9 |
| **BigBird Base Model Finetuned on Medical Corpus** |  |  |  |  |  |  |  |  |
| nbroad/bigbird-base-health-fact | 0.221 | 0.872 | 0.918 | 0.924 | 0.920 | 0.992 | 0.991 | 199.9 |
| **Bert Base Models Finetuned on Non Related Corpus** |  |  |  |  |  |  |  |  |
| ahmedrachid/FinancialBERT-Sentiment-Analysis | 0.214 | 0.876 | 0.920 | 0.924 | 0.922 | 0.992 | 0.991 | 58.0 |
| MathBERT | 0.220 | 0.879 | 0.920 | 0.924 | 0.922 | 0.991 | 0.989 | 58.5 |
| **DistilBert Base Model Finetuned on Non Related Corpus** |  |  |  |  |  |  |  |  |
| distilbert-base-uncased-finetuned-sst-2-english | 0.210 | 0.876 | 0.919 | 0.924 | 0.921 | 0.991 | 0.989 | 24.4 |
| bhadresh-savani/distilbert-base-uncased-emotion | 0.207 | 0.878 | 0.921 | 0.925 | 0.922 | 0.993 | 0.991 | 24.4 |
| **RoBERTa Base Model Finetuned on Non Related Corpus** |  |  |  |  |  |  |  |  |
| cardiffnlp/twitter-roberta-base | 0.210 | 0.878 | 0.923 | 0.927 | 0.924 | 0.993 | 0.992 | 57.5 |

**Table S9.**
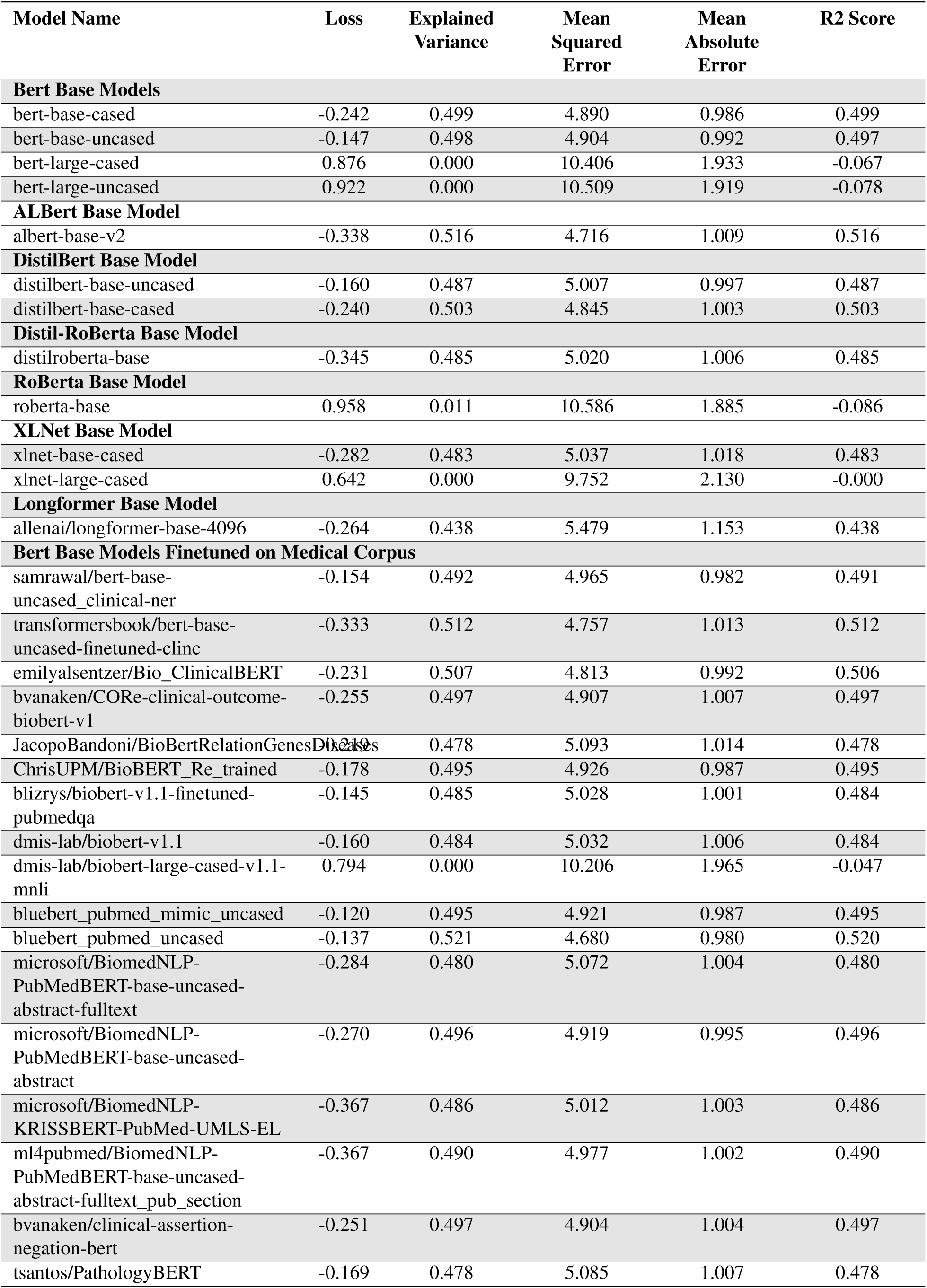
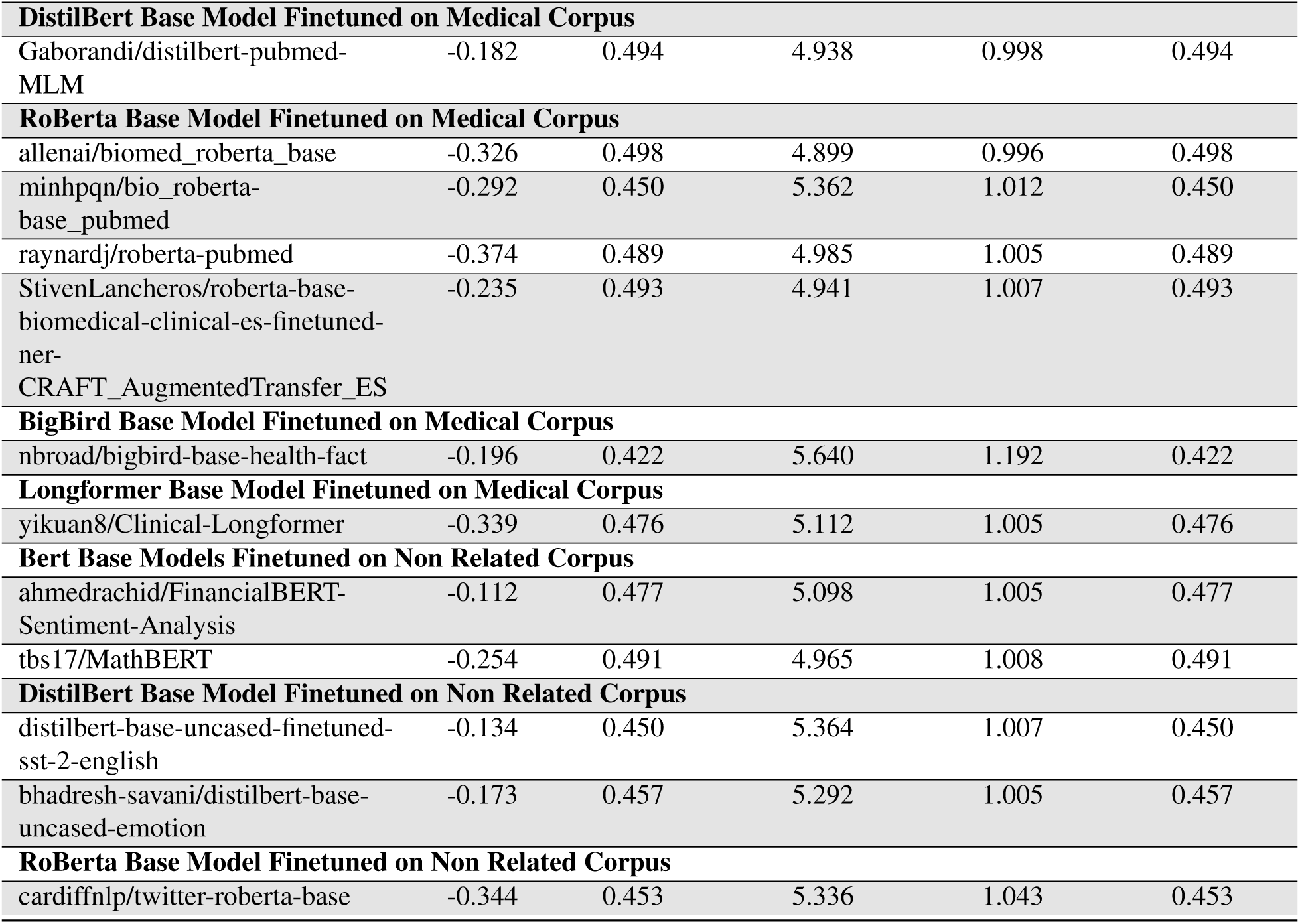
Sign-out Time Regression Results Across All LLMs, broken down by domain relevance of pretraining corpora.

| Model Name | Loss | Explained Variance | Mean Squared Error | Mean Absolute Error | R2 Score |
| --- | --- | --- | --- | --- | --- |
| **Bert Base Models** |  |  |  |  |  |
| bert-base-cased | -0.242 | 0.499 | 4.890 | 0.986 | 0.499 |
| bert-base-uncased | -0.147 | 0.498 | 4.904 | 0.992 | 0.497 |
| bert-large-cased | 0.876 | 0.000 | 10.406 | 1.933 | -0.067 |
| bert-large-uncased | 0.922 | 0.000 | 10.509 | 1.919 | -0.078 |
| **ALBert Base Model** |  |  |  |  |  |
| albert-base-v2 | -0.338 | 0.516 | 4.716 | 1.009 | 0.516 |
| **DistilBert Base Model** |  |  |  |  |  |
| distilbert-base-uncased | -0.160 | 0.487 | 5.007 | 0.997 | 0.487 |
| distilbert-base-cased | -0.240 | 0.503 | 4.845 | 1.003 | 0.503 |
| **Distil-RoBerta Base Model** |  |  |  |  |  |
| distilroberta-base | -0.345 | 0.485 | 5.020 | 1.006 | 0.485 |
| **RoBerta Base Model** |  |  |  |  |  |
| roberta-base | 0.958 | 0.011 | 10.586 | 1.885 | -0.086 |
| **XLNet Base Model** |  |  |  |  |  |
| xlnet-base-cased | -0.282 | 0.483 | 5.037 | 1.018 | 0.483 |
| xlnet-large-cased | 0.642 | 0.000 | 9.752 | 2.130 | -0.000 |
| **Longformer Base Model** |  |  |  |  |  |
| allenai/longformer-base-4096 | -0.264 | 0.438 | 5.479 | 1.153 | 0.438 |
| **Bert Base Models Finetuned on Medical Corpus** |  |  |  |  |  |
| samrawal/bert-base-uncased-clinical-ner | -0.154 | 0.492 | 4.965 | 0.982 | 0.491 |
| transformersbook/bert-base-uncased-finetuned-clinc | -0.333 | 0.512 | 4.757 | 1.013 | 0.512 |
| emilyalsentzer/Bio_ClinicalBERT | -0.231 | 0.507 | 4.813 | 0.992 | 0.506 |
| bvanaken/CORE-clinical-outcome-biobert-v1 | -0.255 | 0.497 | 4.907 | 1.007 | 0.497 |
| JacopoBandoni/BioBERTRelationGenesDiseases | -0.219 | 0.478 | 5.093 | 1.014 | 0.478 |
| ChrisUPM/BioBERT_Re_trained | -0.178 | 0.495 | 4.926 | 0.987 | 0.495 |
| blizrys/biobert-v1.1-finetuned-pubmedqa | -0.145 | 0.485 | 5.028 | 1.001 | 0.484 |
| dmis-lab/biobert-v1.1 | -0.160 | 0.484 | 5.032 | 1.006 | 0.484 |
| dmis-lab/biobert-large-cased-v1.1-mnli | 0.794 | 0.000 | 10.206 | 1.965 | -0.047 |
| bluebert_pubmed_mimic_uncased | -0.120 | 0.495 | 4.921 | 0.987 | 0.495 |
| bluebert_pubmed_uncased | -0.137 | 0.521 | 4.680 | 0.980 | 0.520 |
| microsoft/BiomedNLP-PubMedBERT-base-uncased-abstract-fulltext | -0.284 | 0.480 | 5.072 | 1.004 | 0.480 |
| microsoft/BiomedNLP-PubMedBERT-base-uncased-abstract | -0.270 | 0.496 | 4.919 | 0.995 | 0.496 |
| microsoft/BiomedNLP-KRISSBERT-PubMed-UMLS-EL | -0.367 | 0.486 | 5.012 | 1.003 | 0.486 |
| ml4pubmed/BiomedNLP-PubMedBERT-base-uncased-abstract-fulltext_pub_section | -0.367 | 0.490 | 4.977 | 1.002 | 0.490 |
| bvanaken/clinical-assertion-negation-bert | -0.251 | 0.497 | 4.904 | 1.004 | 0.497 |
| tsantos/PathologyBERT | -0.169 | 0.478 | 5.085 | 1.007 | 0.478 |
| **DistilBert Base Model Finetuned on Medical Corpus** |  |  |  |  |  |
| Gaborandi/distilbert-pubmed-MLM | -0.182 | 0.494 | 4.938 | 0.998 | 0.494 |
| **RoBerta Base Model Finetuned on Medical Corpus** |  |  |  |  |  |
| allenai/biomed_roberta_base | -0.326 | 0.498 | 4.899 | 0.996 | 0.498 |
| minhpqn/bio_roberta-base_pubmed | -0.292 | 0.450 | 5.362 | 1.012 | 0.450 |
| raynardj/roberta-pubmed | -0.374 | 0.489 | 4.985 | 1.005 | 0.489 |
| StivenLancheros/roberta-base-biomedical-clinical-es-finetuned-ner-CRAFT_AugmentedTransfer_ES | -0.235 | 0.493 | 4.941 | 1.007 | 0.493 |
| **BigBird Base Model Finetuned on Medical Corpus** |  |  |  |  |  |
| nbroad/bigbird-base-health-fact | -0.196 | 0.422 | 5.640 | 1.192 | 0.422 |
| **Longformer Base Model Finetuned on Medical Corpus** |  |  |  |  |  |
| yikuan8/Clinical-Longformer | -0.339 | 0.476 | 5.112 | 1.005 | 0.476 |
| **Bert Base Models Finetuned on Non Related Corpus** |  |  |  |  |  |
| ahmedrachid/FinancialBERT-Sentiment-Analysis | -0.112 | 0.477 | 5.098 | 1.005 | 0.477 |
| tbs17/MathBERT | -0.254 | 0.491 | 4.965 | 1.008 | 0.491 |
| **DistilBert Base Model Finetuned on Non Related Corpus** |  |  |  |  |  |
| distilbert-base-uncased-finetuned-sst-2-english | -0.134 | 0.450 | 5.364 | 1.007 | 0.450 |
| bhadresh-savani/distilbert-base-uncased-emotion | -0.173 | 0.457 | 5.292 | 1.005 | 0.457 |
| **RoBerta Base Model Finetuned on Non Related Corpus** |  |  |  |  |  |
| cardiffnlp/twitter-roberta-base | -0.344 | 0.453 | 5.336 | 1.043 | 0.453 |

**Table S10.** Text Generation (Next 5 Tokens) Results: Accuracies for correct assignment of five subsequent words reported at varying percentiles of seed text.

| Model Name | 0 % | 20 % | 40 % | 60 % | 80 % |
| --- | --- | --- | --- | --- | --- |
| **GPT2 Base Models** |  |  |  |  |  |
| sshleifer_tiny-gpt2 | 0.004 | 0.003 | 0.003 | 0.003 | 0.002 |
| gpt2 | 0.582 | 0.567 | 0.583 | 0.610 | 0.653 |
| gpt2-medium | 0.603 | 0.604 | 0.617 | 0.643 | 0.682 |
| gpt2-large | 0.618 | 0.626 | 0.639 | 0.667 | 0.700 |
| **GPT2 Base Models Finetuned on Medical Corpus** |  |  |  |  |  |
| stanford-crfm_pubmed_gpt | 0.519 | 0.488 | 0.492 | 0.517 | 0.572 |
| **EleutherAI's replication of the GPT-3 architecture Base Models** |  |  |  |  |  |
| EleutherAI_gpt-neo-125M | 0.396 | 0.414 | 0.419 | 0.441 | 0.396 |
| **OPT Base Models** |  |  |  |  |  |
| facebook_opt-125m | 0.025 | 0.026 | 0.033 | 0.034 | 0.075 |
| **DistilGPT2 Base Models** |  |  |  |  |  |
| distilgpt2 | 0.561 | 0.546 | 0.553 | 0.580 | 0.624 |

**Table S11.** Text Generation (Next 3 Tokens) Results: Accuracies for correct assignment of three subsequent words reported at varying percentiles of seed text.

| Model Name | 0 % | 20 % | 40 % | 60 % | 80 % |
| --- | --- | --- | --- | --- | --- |
| **GPT2 Base Models** |  |  |  |  |  |
| sshleifer_tiny-gpt2 | 0.004 | 0.003 | 0.003 | 0.003 | 0.002 |
| gpt2 | 0.576 | 0.654 | 0.667 | 0.675 | 0.712 |
| gpt2-medium | 0.603 | 0.687 | 0.696 | 0.705 | 0.737 |
| gpt2-large | 0.617 | 0.705 | 0.715 | 0.724 | 0.755 |
| **GPT2 Base Models Finetuned on Medical Corpus** |  |  |  |  |  |
| stanford-crfm_pubmed_gpt | 0.429 | 0.580 | 0.580 | 0.589 | 0.636 |
| **EleutherAI's replication of the GPT-3 architecture Base Models** |  |  |  |  |  |
| EleutherAI_gpt-neo-125M | 0.309 | 0.378 | 0.396 | 0.415 | 0.430 |
| **OPT Base Models** |  |  |  |  |  |
| facebook_opt-125m | 0.016 | 0.020 | 0.025 | 0.035 | 0.033 |
| **DistilGPT2 Base Models** |  |  |  |  |  |
| distilgpt2 | 0.558 | 0.634 | 0.640 | 0.648 | 0.686 |

**Figure S4.**
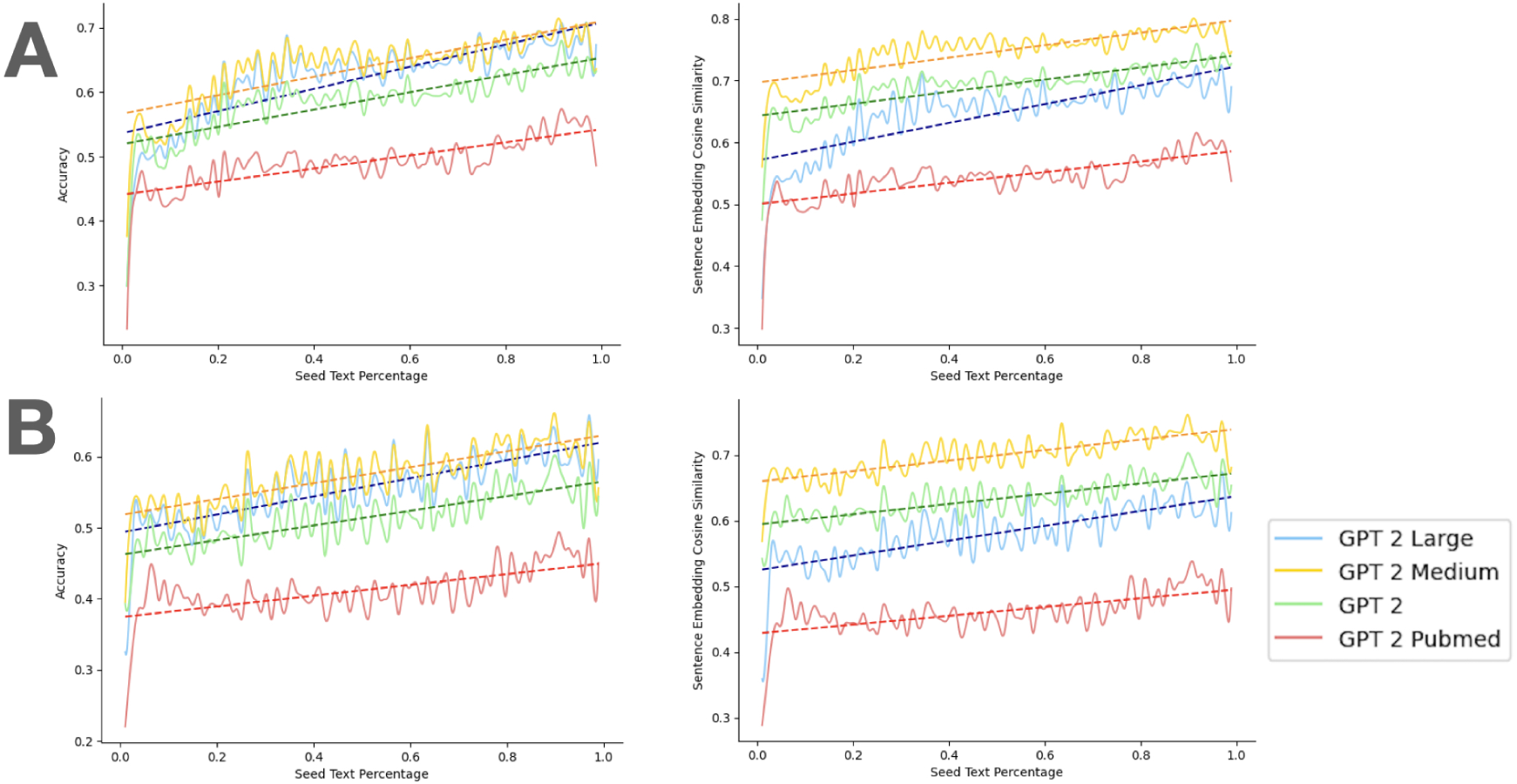
Text generation results with different training iterations: A. Predict next 3 tokens. Accuracy score (left), word embedding cosine similarity (right). B. Predict next 5 tokens. Accuracy score (left), word embedding cosine similarity (right)

**Table S12.**
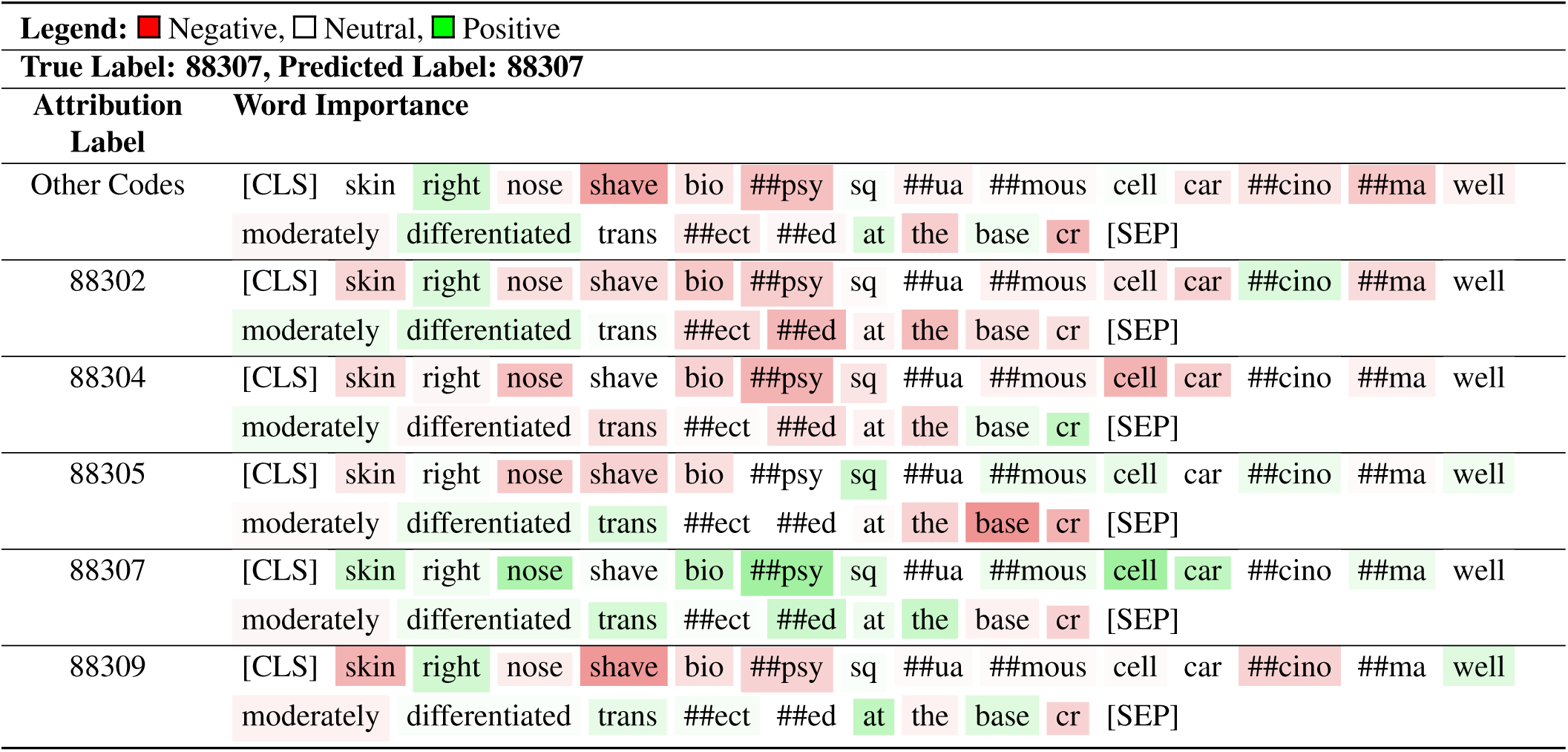
Bert base uncased model attribution interpretation: CPT Code Classification Example 1; ## indicates presence of subtokens.

**Table S13.**
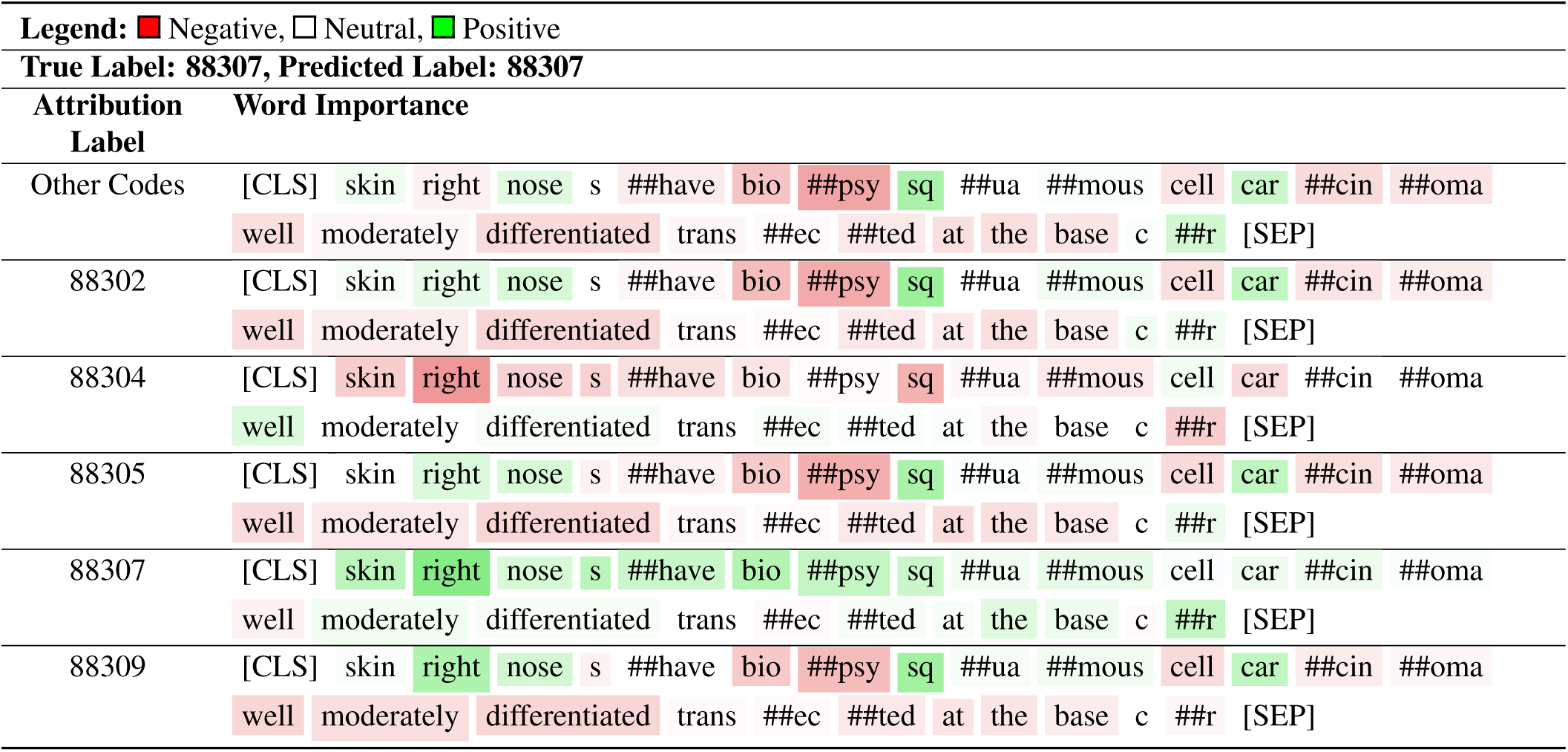
Clinical Bert model attribution interpretation: CPT Code Classification Example 1.

**Table S14.**
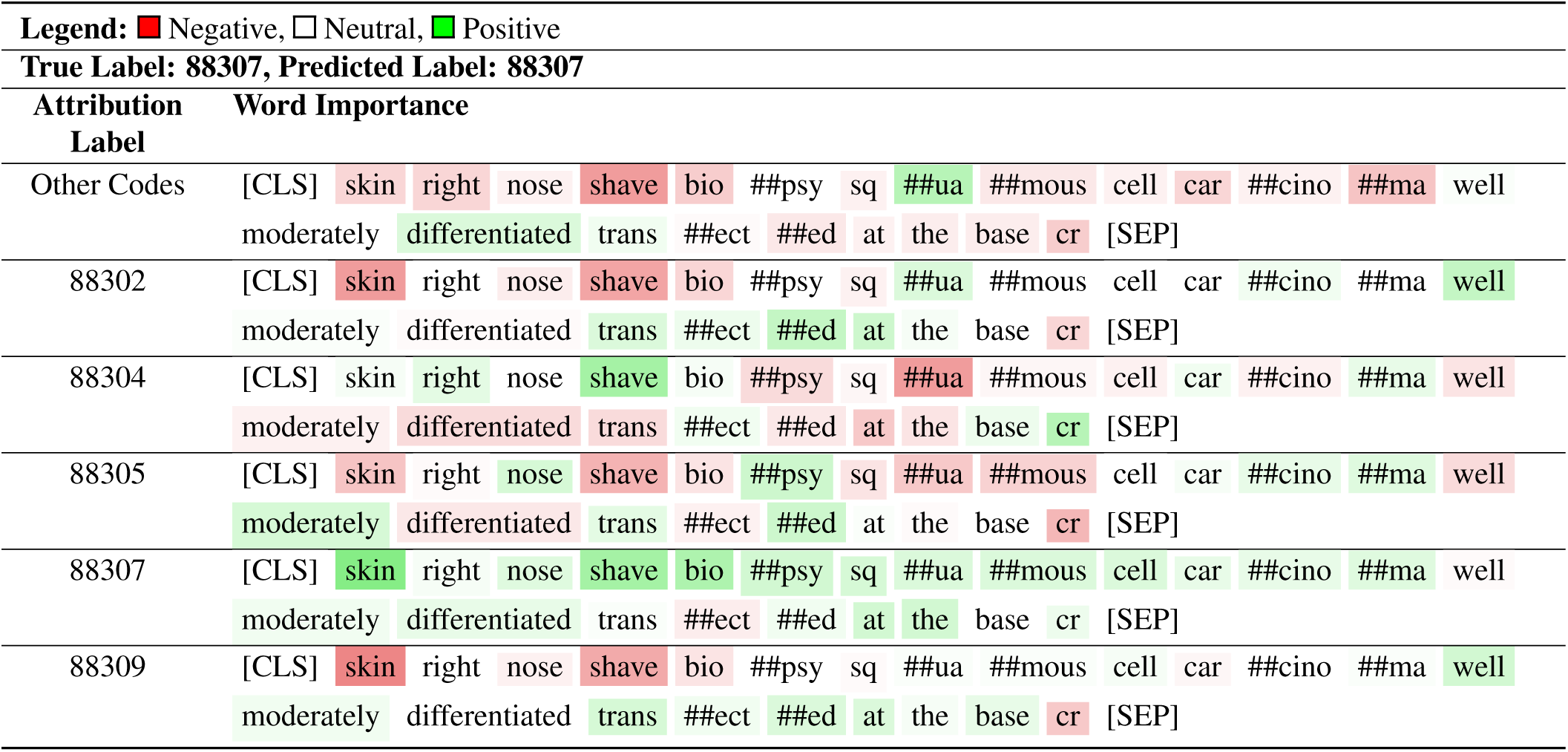
Math Bert model attribution interpretation: CPT Code Classification Example 1.

**Table S15.**
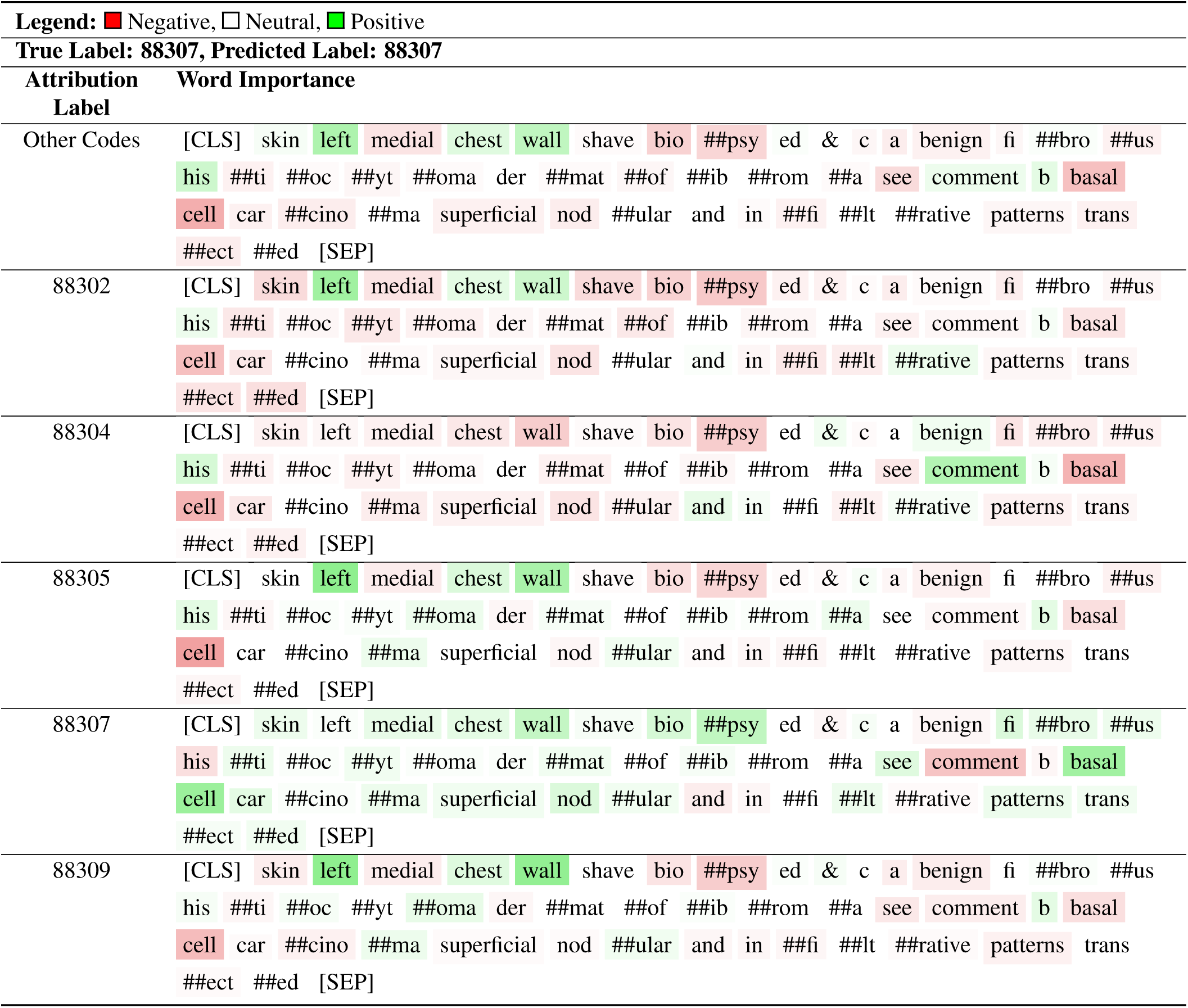
Bert base uncased model attribution interpretation: CPT Code Classification Example 2.

**Table S16.**
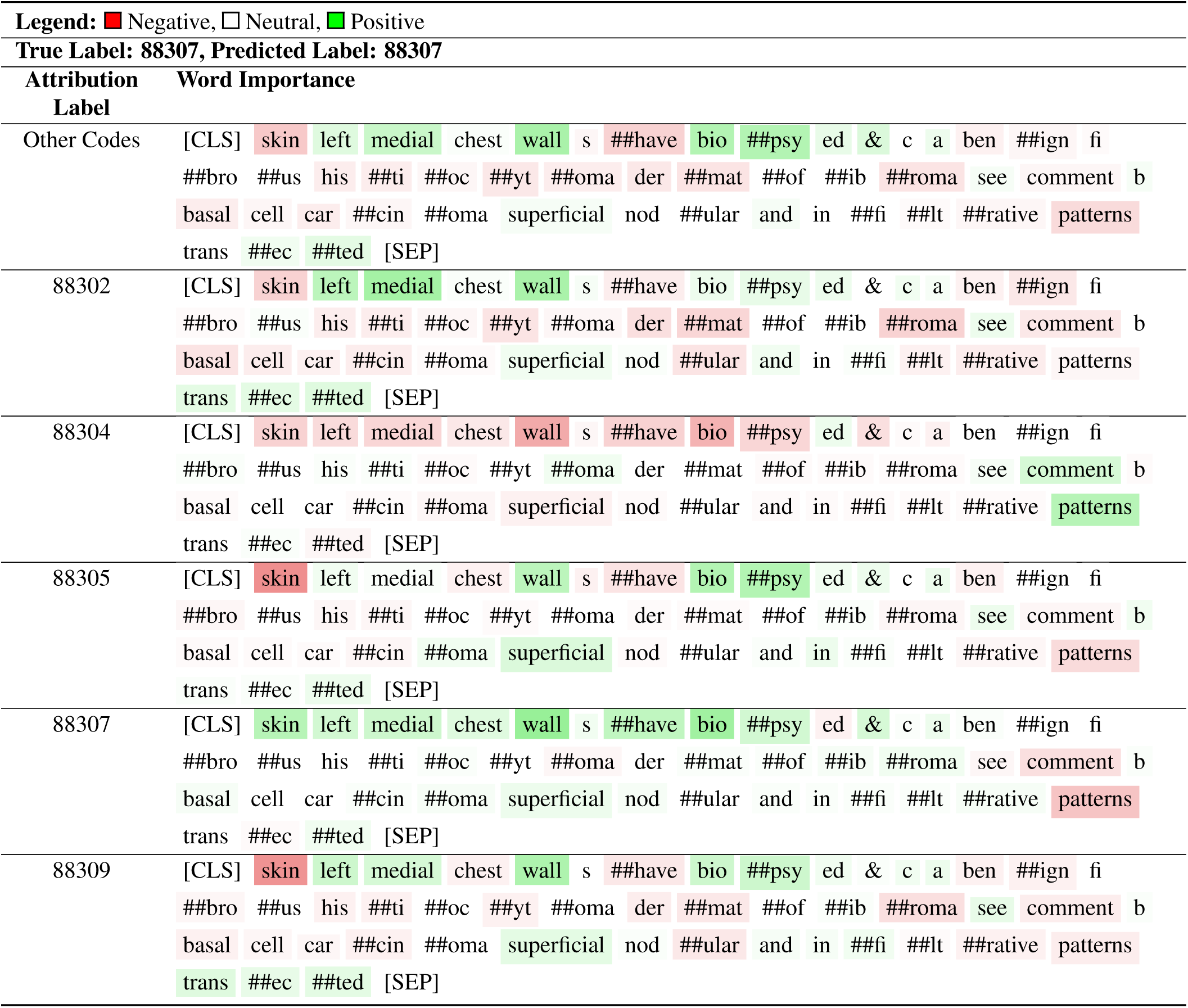
Clinical Bert model attribution interpretation: CPT Code Classification Example 2.

**Table S17.**
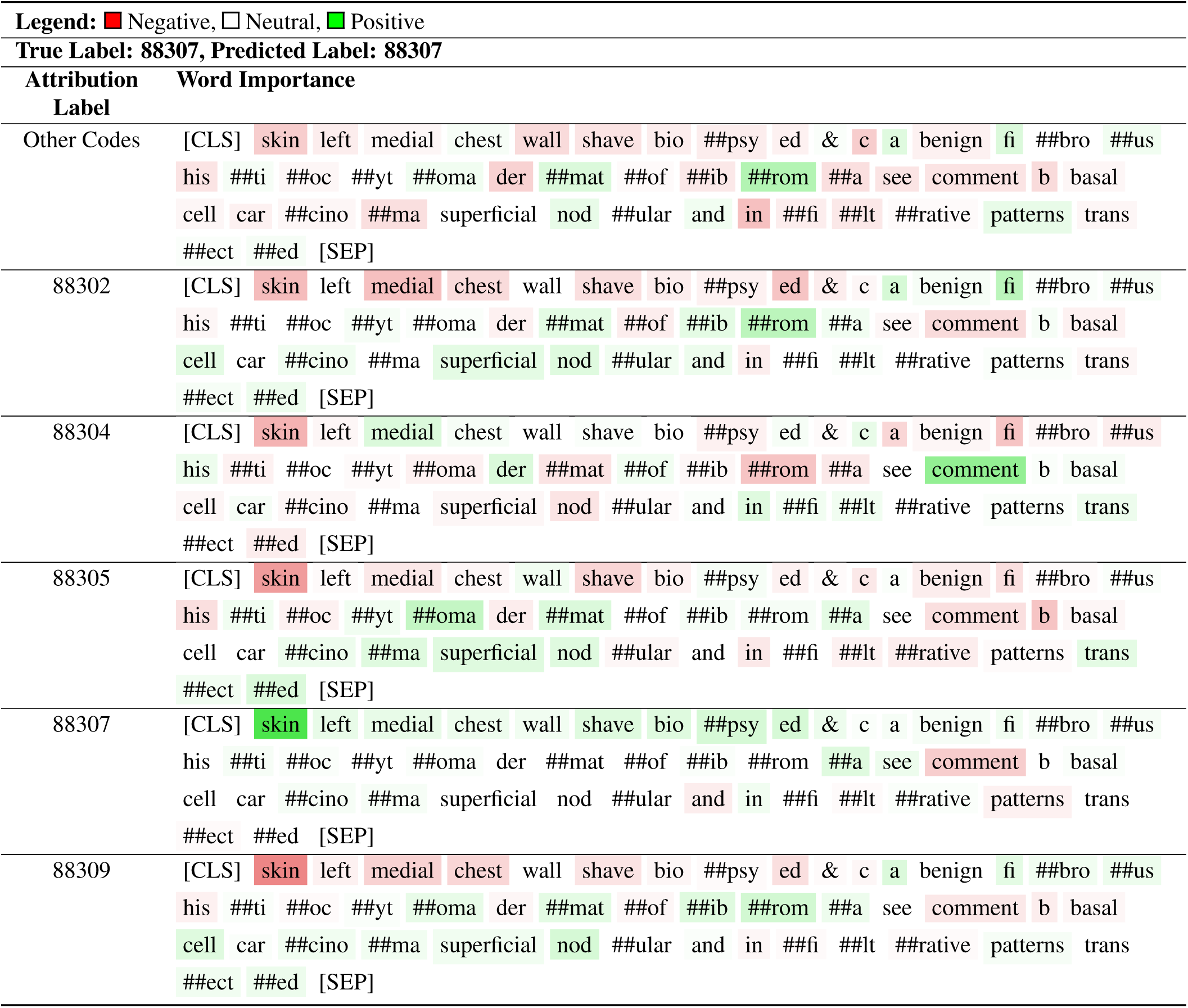
Math Bert model attribution interpretation: CPT Code Classification Example 2.

**Table S18.**
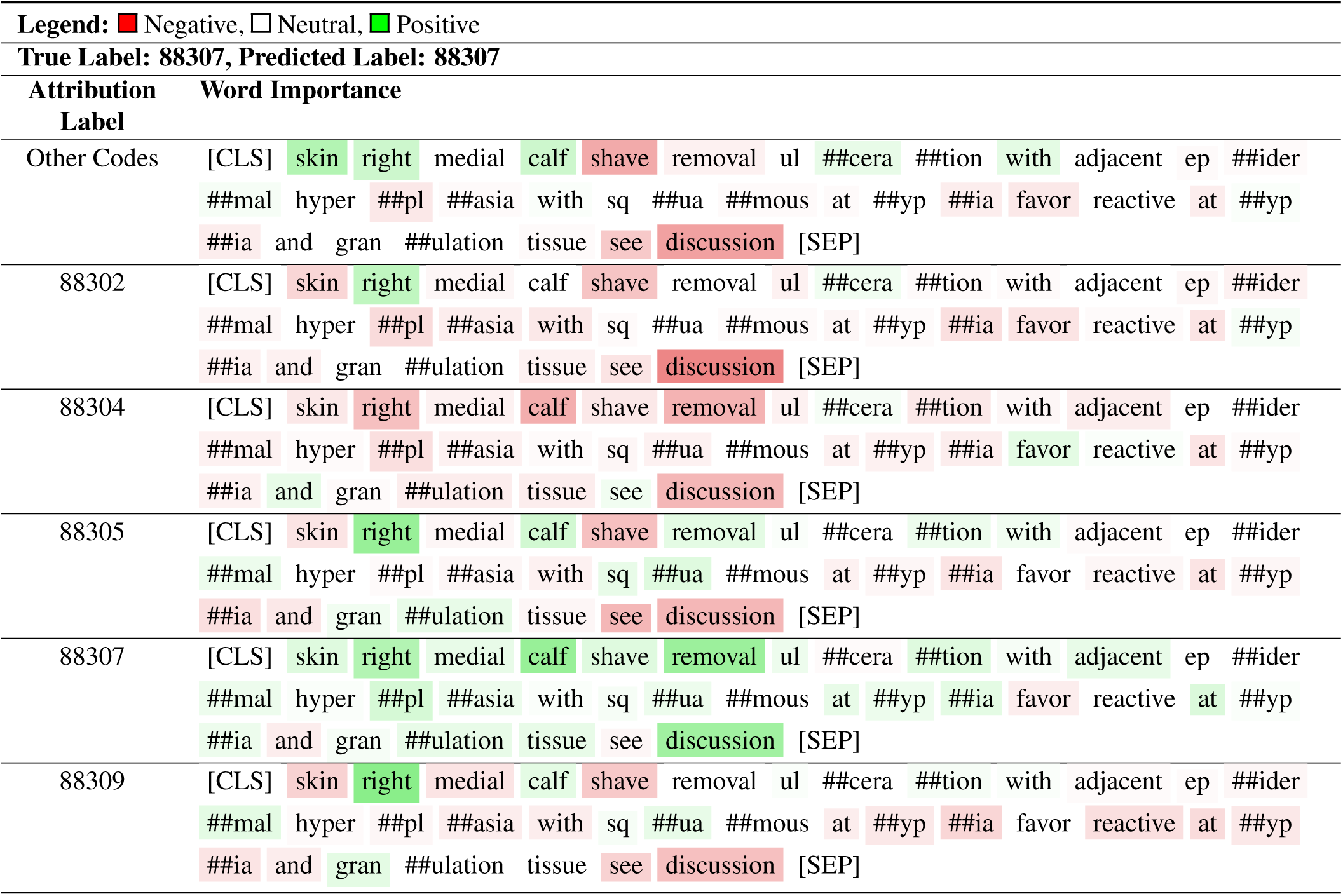
Bert base uncased model attribution interpretation: CPT Code Classification Example 3.

**Table S19.** Clinical Bert model attribution interpretation: CPT Code Classification Example 3.

| **Legend:** ■ Negative, □ Neutral, ■ Positive |  |
| --- | --- |
| **True Label: 88307, Predicted Label: 88307** |  |
| Attribution Label | Word Importance |
| Other Codes | [CLS] skin right medial calf s ##have removal ul ##cer ##ation with adjacent e<br>##pid ##er ##mal h ##yper ##p ##lasia with sq ##ua ##mous at ##y ##pia favor<br>reactive at ##y ##pia and g ##ran ##ulation tissue see discussion [SEP] |
| 88302 | [CLS] skin right medial calf s ##have removal ul ##cer ##ation with adjacent e<br>##pid ##er ##mal h ##yper ##p ##lasia with sq ##ua ##mous at ##y ##pia favor<br>reactive at ##y ##pia and g ##ran ##ulation tissue see discussion [SEP] |
| 88304 | [CLS] skin right medial calf s ##have removal ul ##cer ##ation with adjacent e<br>##pid ##er ##mal h ##yper ##p ##lasia with sq ##ua ##mous at ##y ##pia favor<br>reactive at ##y ##pia and g ##ran ##ulation tissue see discussion [SEP] |
| 88305 | [CLS] skin right medial calf s ##have removal ul ##cer ##ation with adjacent e<br>##pid ##er ##mal h ##yper ##p ##lasia with sq ##ua ##mous at ##y ##pia favor<br>reactive at ##y ##pia and g ##ran ##ulation tissue see discussion [SEP] |
| 88307 | [CLS] skin right medial calf s ##have removal ul ##cer ##ation with adjacent e<br>##pid ##er ##mal h ##yper ##p ##lasia with sq ##ua ##mous at ##y ##pia favor<br>reactive at ##y ##pia and g ##ran ##ulation tissue see discussion [SEP] |
| 88309 | [CLS] skin right medial calf s ##have removal ul ##cer ##ation with adjacent e<br>##pid ##er ##mal h ##yper ##p ##lasia with sq ##ua ##mous at ##y ##pia favor<br>reactive at ##y ##pia and g ##ran ##ulation tissue see discussion [SEP] |

**Table S20.**
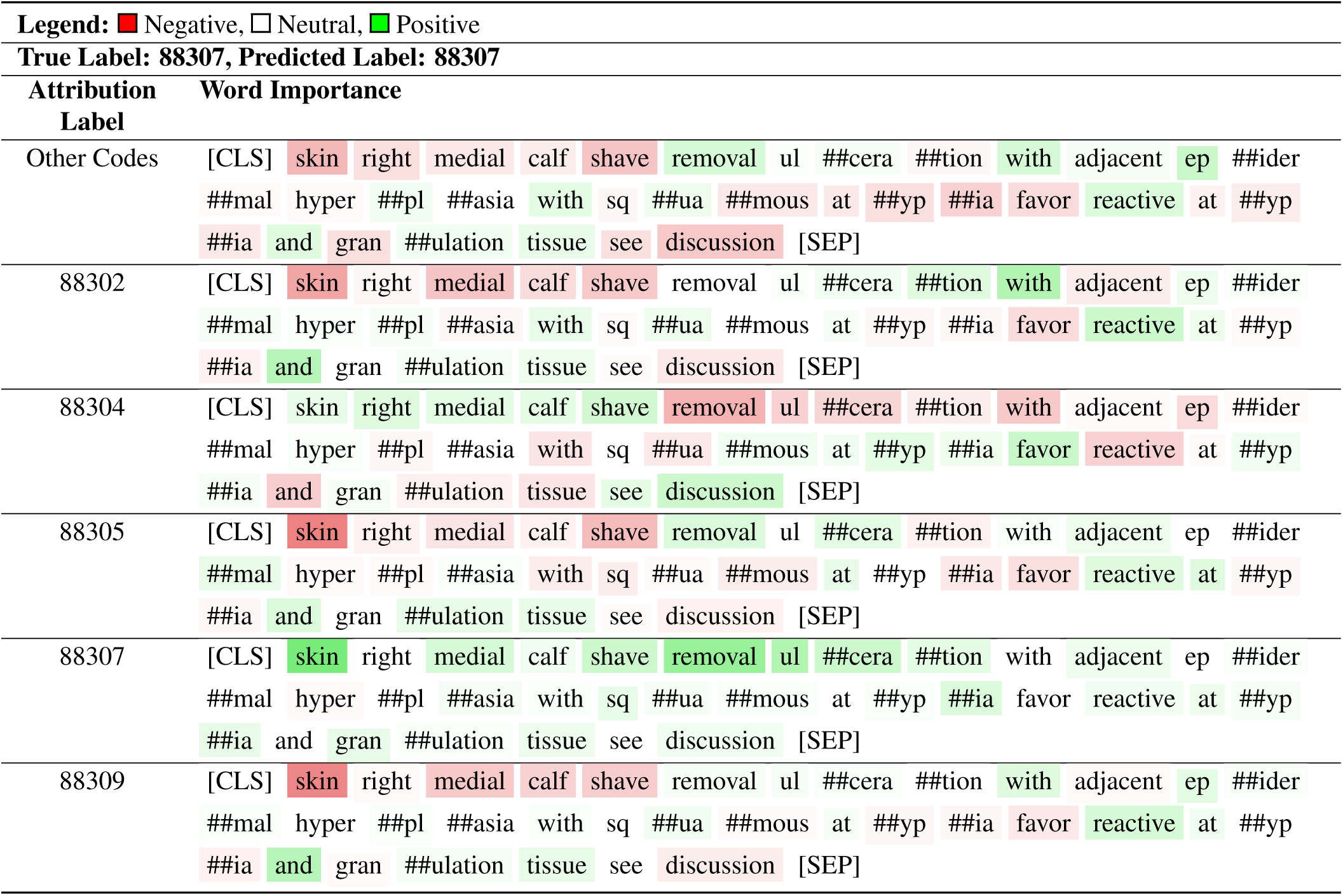
Math Bert model attribution interpretation: CPT Code Classification Example 3.

**Table S21.**
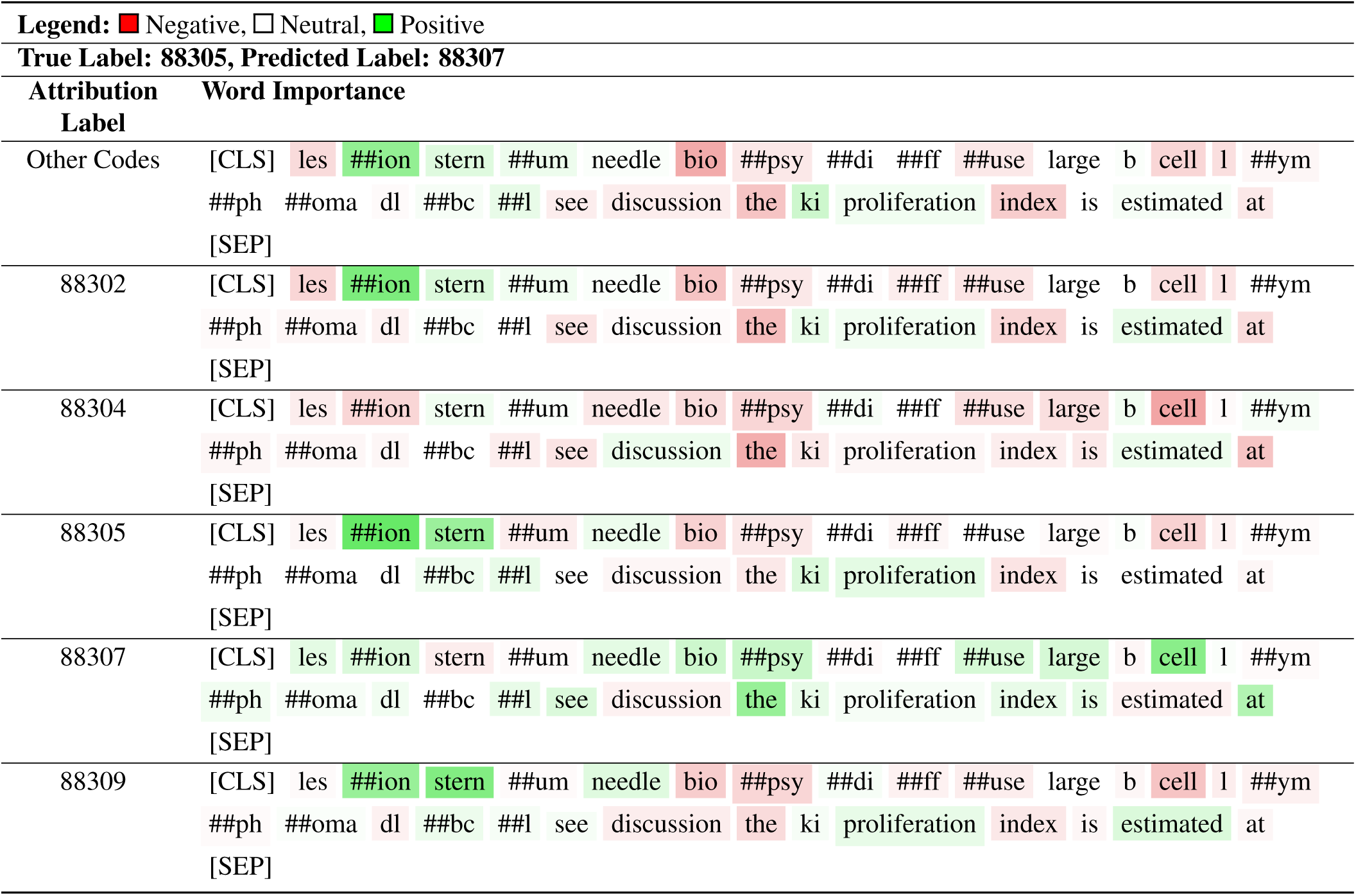
Bert base uncased model attribution interpretation: CPT Code Classification Example 4.

**Table S22.** Clinical Bert model attribution interpretation: CPT Code Classification Example 4.

| Legend: ■ Negative, □ Neutral, ■ Positive |  |
| --- | --- |
| True Label: 88305, Predicted Label: 88307 |  |
| Attribution Label | Word Importance |
| Other Codes | [CLS] les ##ion stern ##um needle bio ##psy ##di ##ff ##use large b cell l<br>##ymph ##oma d ##l ##b ##c ##l see discussion the k ##i proliferation index is<br>estimated at [SEP] |
| 88302 | [CLS] les ##ion stern ##um needle bio ##psy ##di ##ff ##use large b cell l<br>##ymph ##oma d ##l ##b ##c ##l see discussion the k ##i proliferation index is<br>estimated at [SEP] |
| 88304 | [CLS] les ##ion stern ##um needle bio ##psy ##di ##ff ##use large b cell l<br>##ymph ##oma d ##l ##b ##c ##l see discussion the k ##i proliferation index is<br>estimated at [SEP] |
| 88305 | [CLS] les ##ion stern ##um needle bio ##psy ##di ##ff ##use large b cell l<br>##ymph ##oma d ##l ##b ##c ##l see discussion the k ##i proliferation index is<br>estimated at [SEP] |
| 88307 | [CLS] les ##ion stern ##um needle bio ##psy ##di ##ff ##use large b cell l<br>##ymph ##oma d ##l ##b ##c ##l see discussion the k ##i proliferation index is<br>estimated at [SEP] |
| 88309 | [CLS] les ##ion stern ##um needle bio ##psy ##di ##ff ##use large b cell l<br>##ymph ##oma d ##l ##b ##c ##l see discussion the k ##i proliferation index is<br>estimated at [SEP] |

**Table S23.**
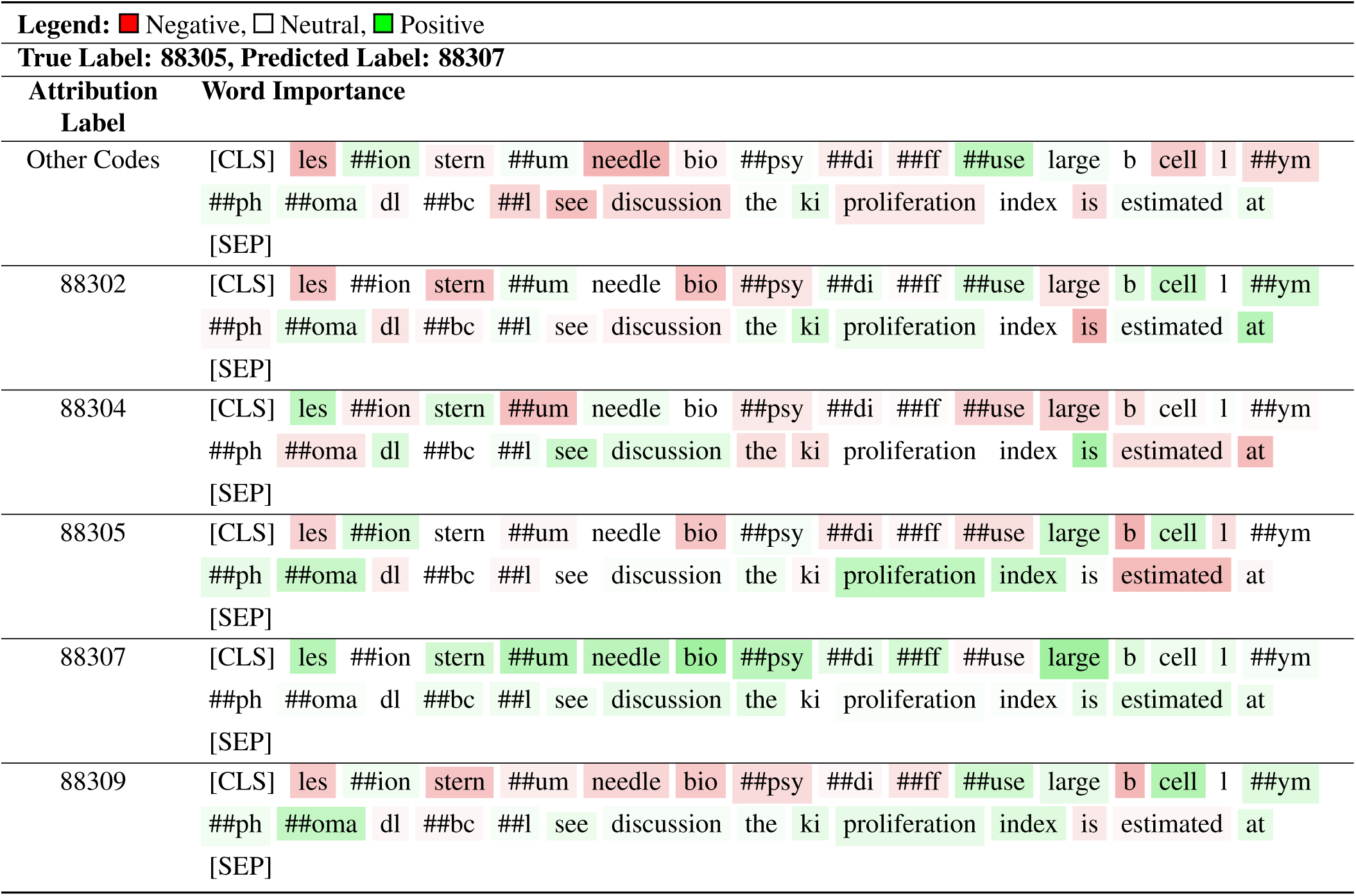
Math Bert model attribution interpretation: CPT Code Classification Example 4.

**Table S24.**
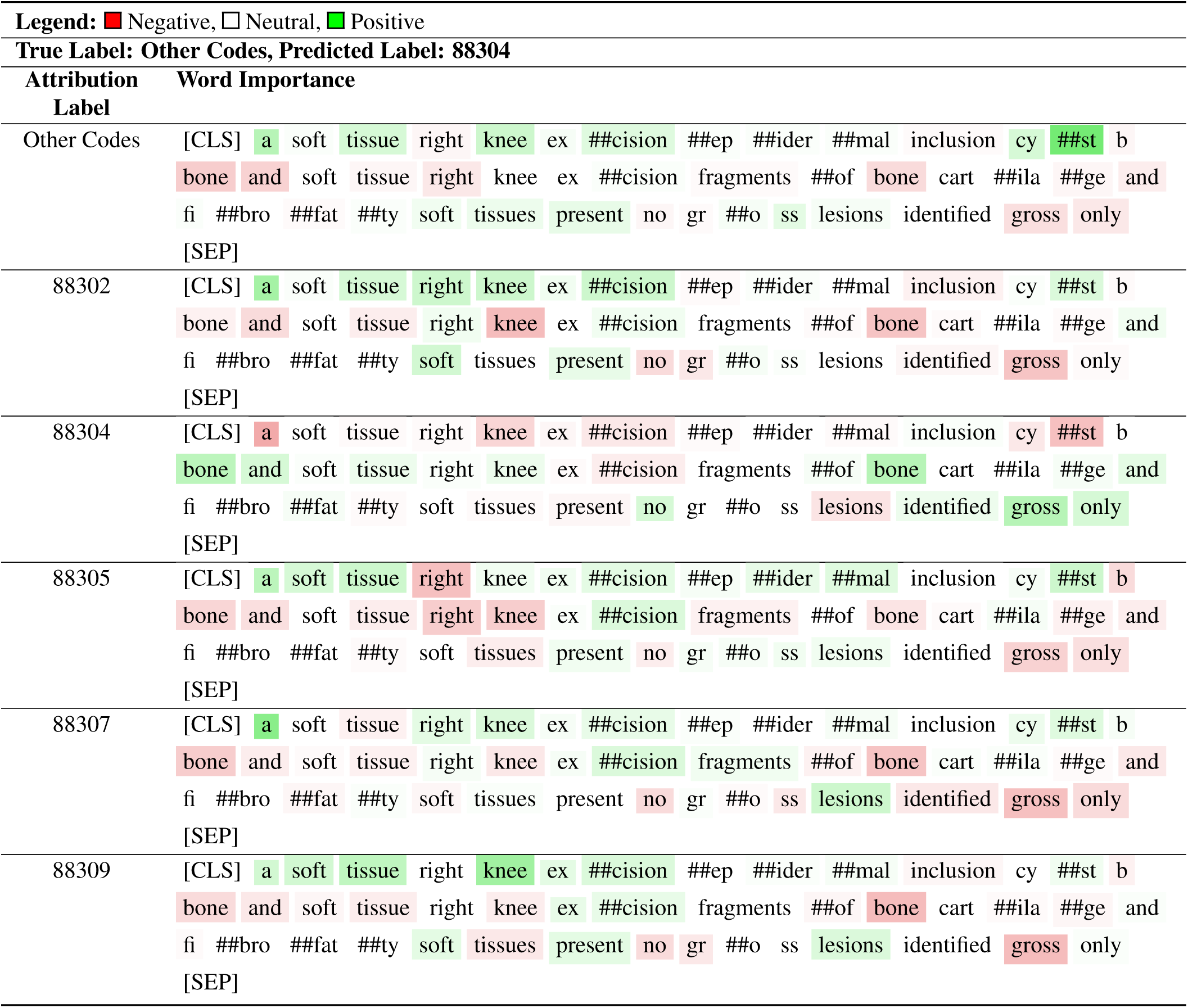
Bert base uncased model attribution interpretation: CPT Code Classification Example 5.

**Table S25.**
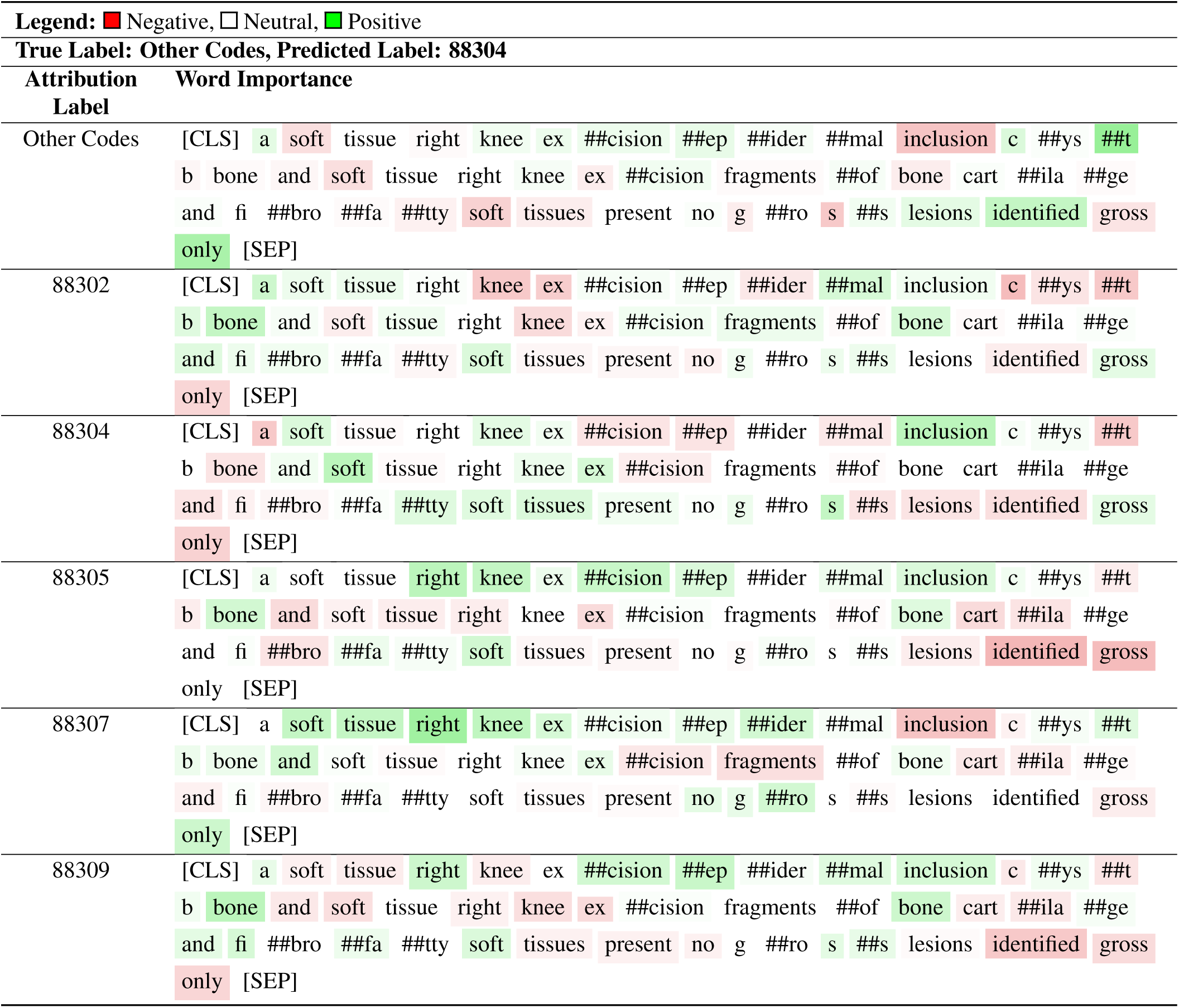
CLinical Bert model attribution interpretation: CPT Code Classification Example 5.

| Legend: ■ Negative, □ Neutral, ■ Positive |  |
| --- | --- |
| True Label: Other Codes, Predicted Label: 88304 |  |
| Attribution Label | Word Importance |
| Other Codes | [CLS] a soft tissue right knee ex ##cision ##ep ##ider ##mal inclusion c ##ys ##t<br>b bone and soft tissue right knee ex ##cision fragments ##of bone cart ##ila ##ge<br>and fi ##bro ##fa ##tty soft tissues present no g ##ro s ##s lesions identified gross<br>only [SEP] |
| 88302 | [CLS] a soft tissue right knee ex ##cision ##ep ##ider ##mal inclusion c ##ys ##t<br>b bone and soft tissue right knee ex ##cision fragments ##of bone cart ##ila ##ge<br>and fi ##bro ##fa ##tty soft tissues present no g ##ro s ##s lesions identified gross<br>only [SEP] |
| 88304 | [CLS] a soft tissue right knee ex ##cision ##ep ##ider ##mal inclusion c ##ys ##t<br>b bone and soft tissue right knee ex ##cision fragments ##of bone cart ##ila ##ge<br>and fi ##bro ##fa ##tty soft tissues present no g ##ro s ##s lesions identified gross<br>only [SEP] |
| 88305 | [CLS] a soft tissue right knee ex ##cision ##ep ##ider ##mal inclusion c ##ys ##t<br>b bone and soft tissue right knee ex ##cision fragments ##of bone cart ##ila ##ge<br>and fi ##bro ##fa ##tty soft tissues present no g ##ro s ##s lesions identified gross<br>only [SEP] |
| 88307 | [CLS] a soft tissue right knee ex ##cision ##ep ##ider ##mal inclusion c ##ys ##t<br>b bone and soft tissue right knee ex ##cision fragments ##of bone cart ##ila ##ge<br>and fi ##bro ##fa ##tty soft tissues present no g ##ro s ##s lesions identified gross<br>only [SEP] |
| 88309 | [CLS] a soft tissue right knee ex ##cision ##ep ##ider ##mal inclusion c ##ys ##t<br>b bone and soft tissue right knee ex ##cision fragments ##of bone cart ##ila ##ge<br>and fi ##bro ##fa ##tty soft tissues present no g ##ro s ##s lesions identified gross<br>only [SEP] |

**Table S26.**
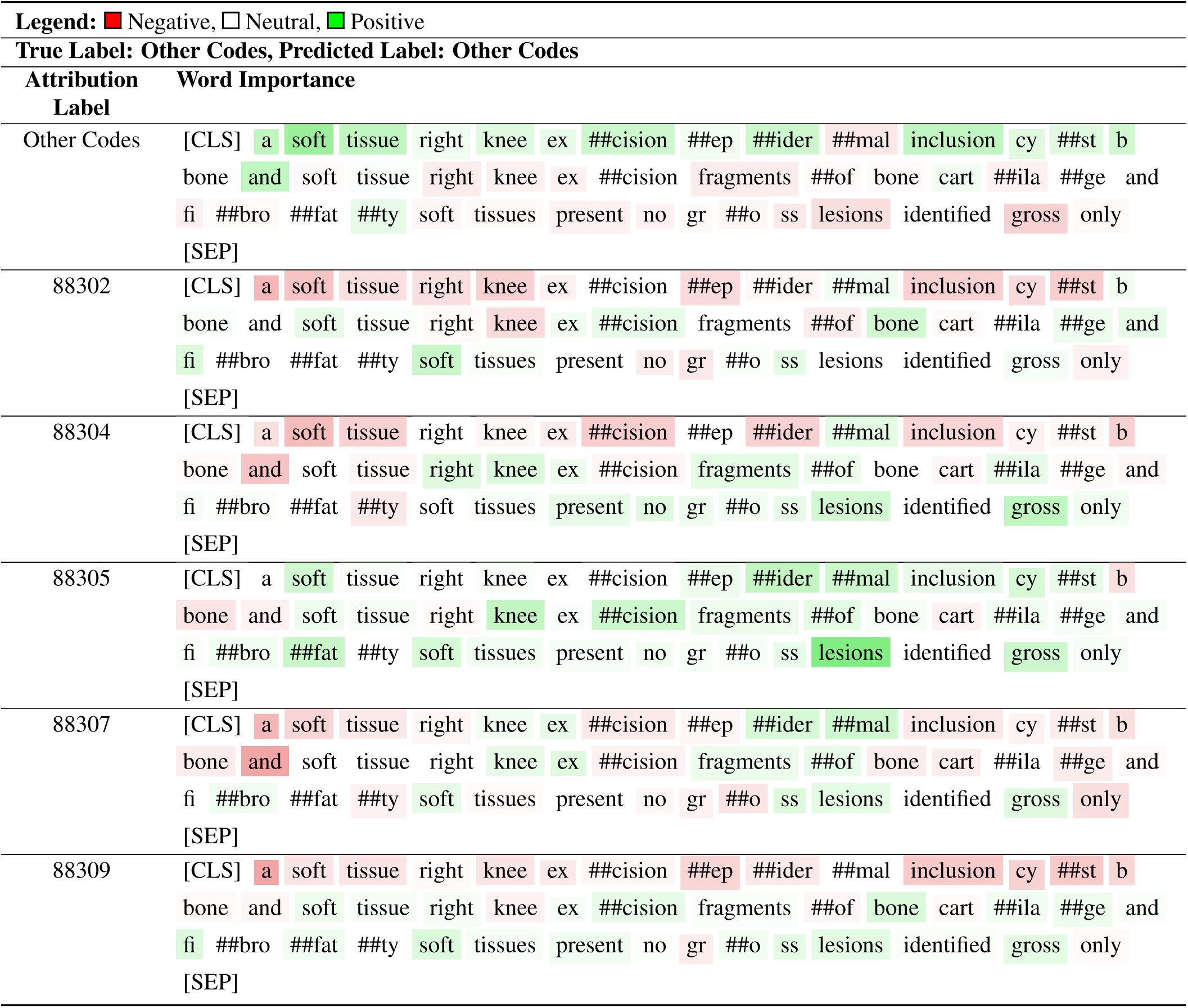
Math Bert model attribution interpretation: CPT Code Classification Example 5.

**Table S27.**
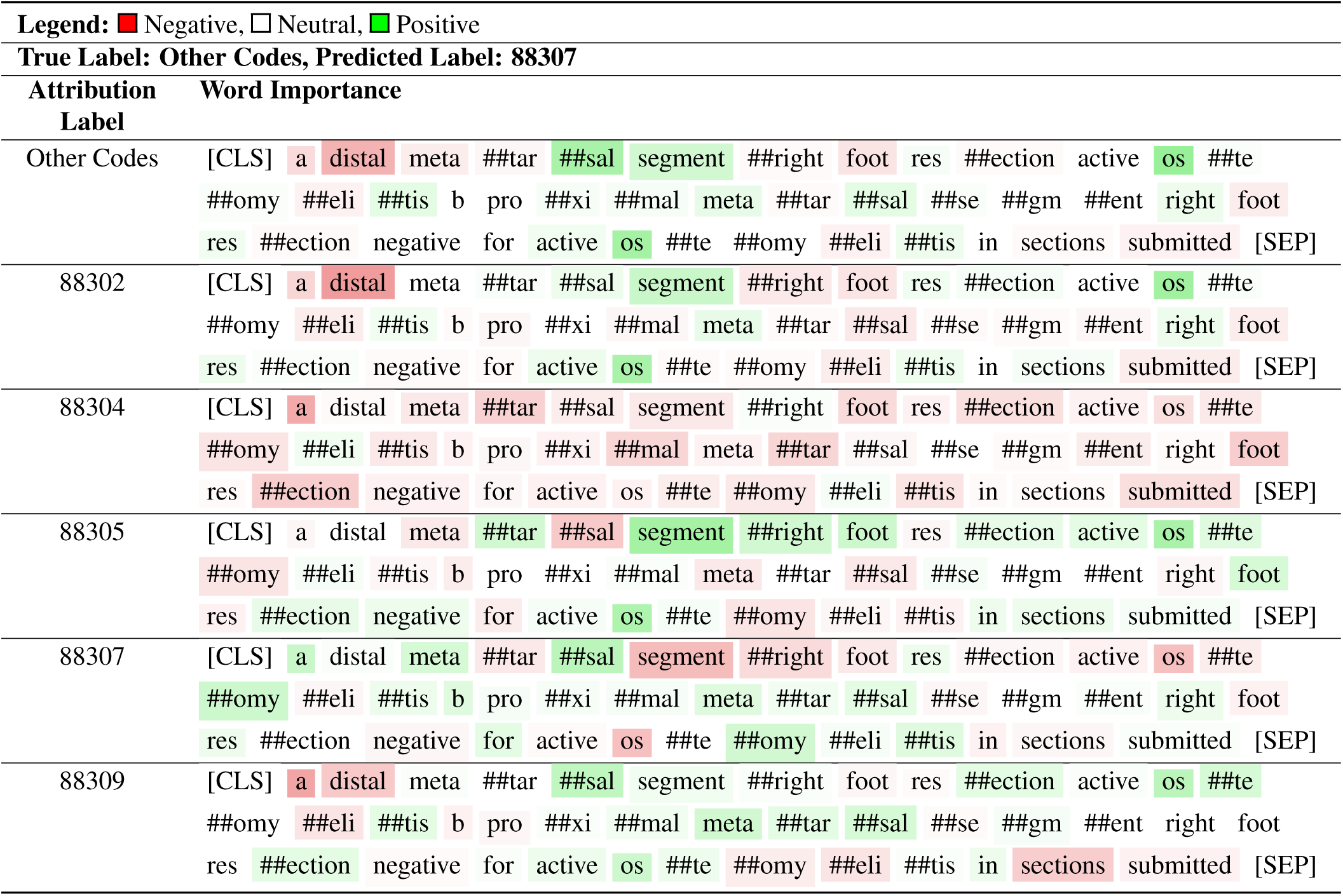
Bert base uncased model attribution interpretation: CPT Code Classification Example 6.

**Table S28.** Clinical Bert model attribution interpretation: CPT Code Classification Example 6.

| Legend: ■ Negative, □ Neutral, ■ Positive |  |
| --- | --- |
| True Label: Other Codes, Predicted Label: 88307 |  |
| Attribution Label | Word Importance |
| Other Codes | [CLS] a di ##stal meta ##tars ##al segment ##right foot re ##section active o ##ste ##omy ##eli ##tis b pro ##ximal meta ##tars ##als ##eg ##ment right foot re ##section negative for active o ##ste ##omy ##eli ##tis in sections submitted [SEP] |
| 88302 | [CLS] a di ##stal meta ##tars ##al segment ##right foot re ##section active o ##ste ##omy ##eli ##tis b pro ##ximal meta ##tars ##als ##eg ##ment right foot re ##section negative for active o ##ste ##omy ##eli ##tis in sections submitted [SEP] |
| 88304 | [CLS] a di ##stal meta ##tars ##al segment ##right foot re ##section active o ##ste ##omy ##eli ##tis b pro ##ximal meta ##tars ##als ##eg ##ment right foot re ##section negative for active o ##ste ##omy ##eli ##tis in sections submitted [SEP] |
| 88305 | [CLS] a di ##stal meta ##tars ##al segment ##right foot re ##section active o ##ste ##omy ##eli ##tis b pro ##ximal meta ##tars ##als ##eg ##ment right foot re ##section negative for active o ##ste ##omy ##eli ##tis in sections submitted [SEP] |
| 88307 | [CLS] a di ##stal meta ##tars ##al segment ##right foot re ##section active o ##ste ##omy ##eli ##tis b pro ##ximal meta ##tars ##als ##eg ##ment right foot re ##section negative for active o ##ste ##omy ##eli ##tis in sections submitted [SEP] |
| 88309 | [CLS] a di ##stal meta ##tars ##al segment ##right foot re ##section active o ##ste ##omy ##eli ##tis b pro ##ximal meta ##tars ##als ##eg ##ment right foot re ##section negative for active o ##ste ##omy ##eli ##tis in sections submitted [SEP] |

**Table S29.**
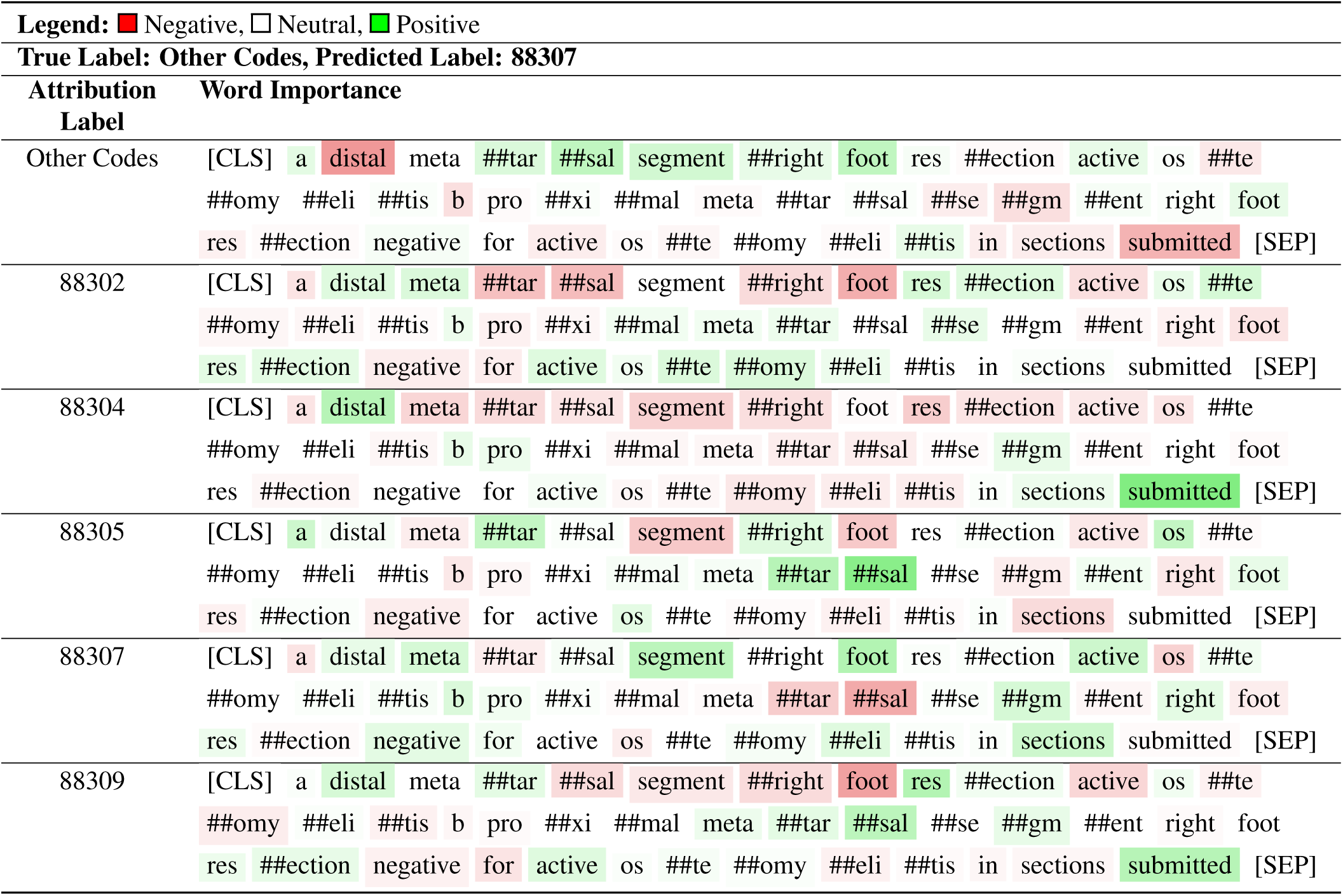
Math Bert model attribution interpretation: CPT Code Classification Example 6.

**Table S30.**
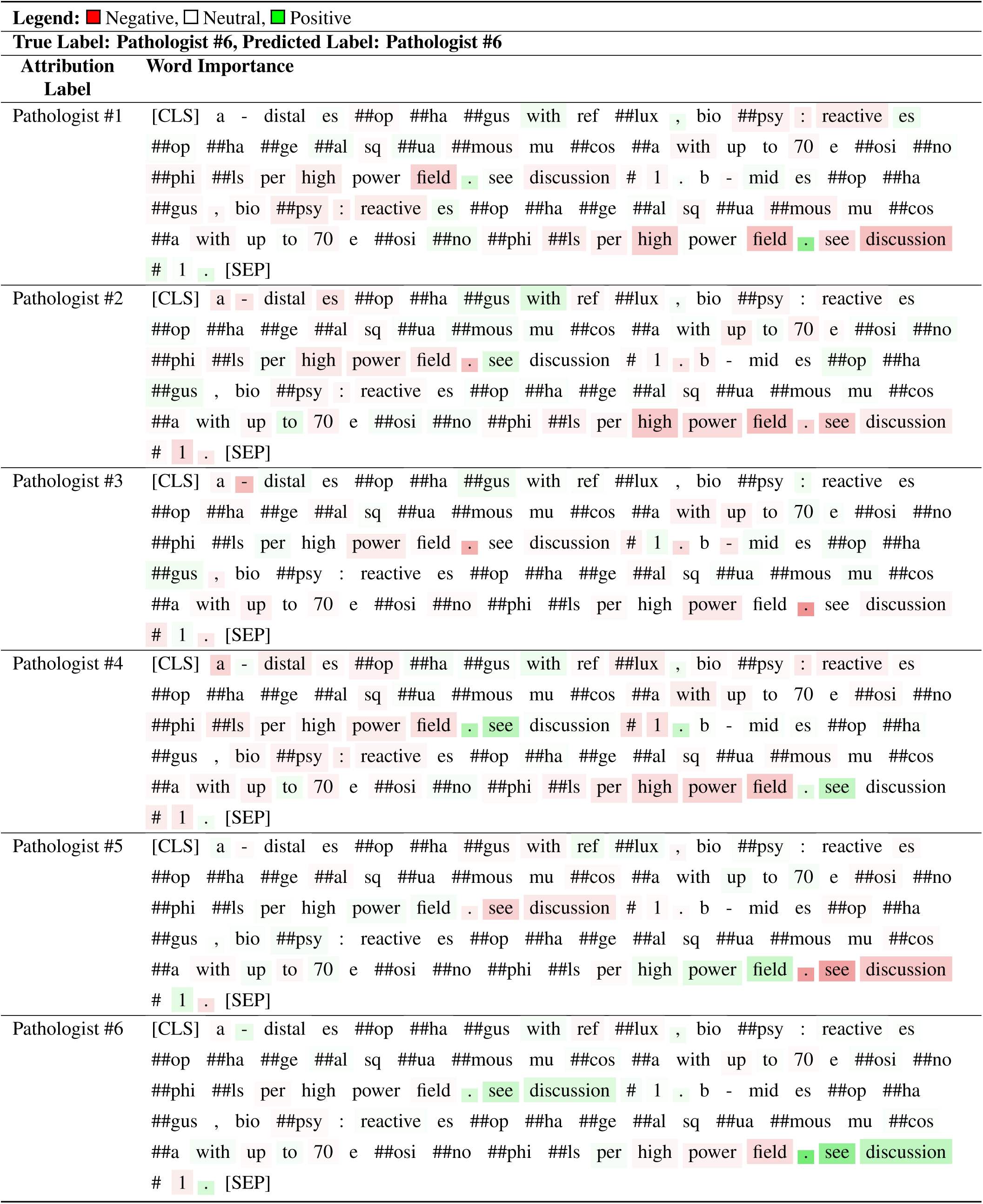
Bert base uncased model attribution interpretation: Pathologist Classification Example 1.

**Table S31.**
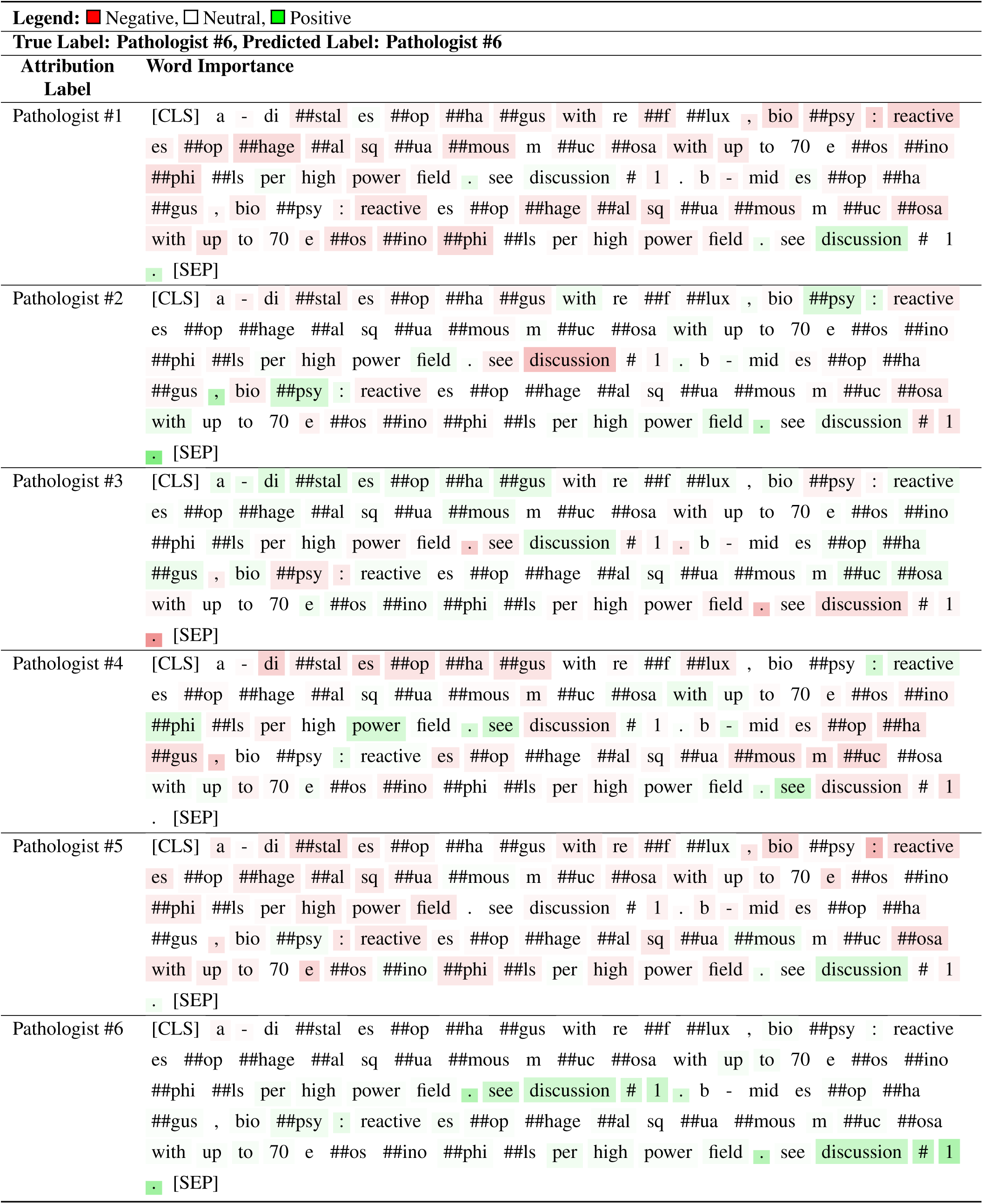
Clinical Bert model attribution interpretation: Pathologist Classification Example 1.

**Table S32.**
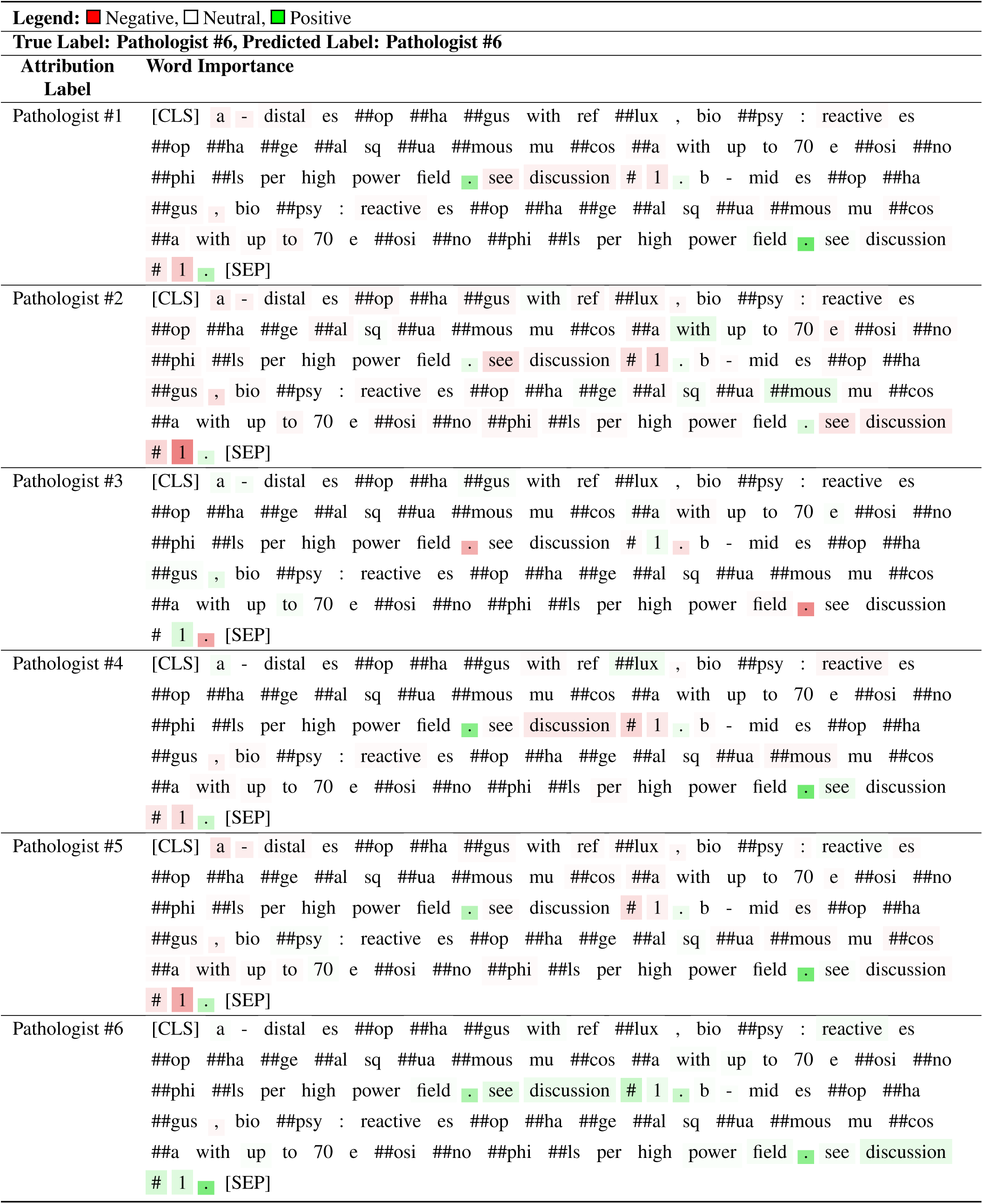
Bert base uncased model attribution interpretation: Pathologist Classification Example 1.

**Table S33.**
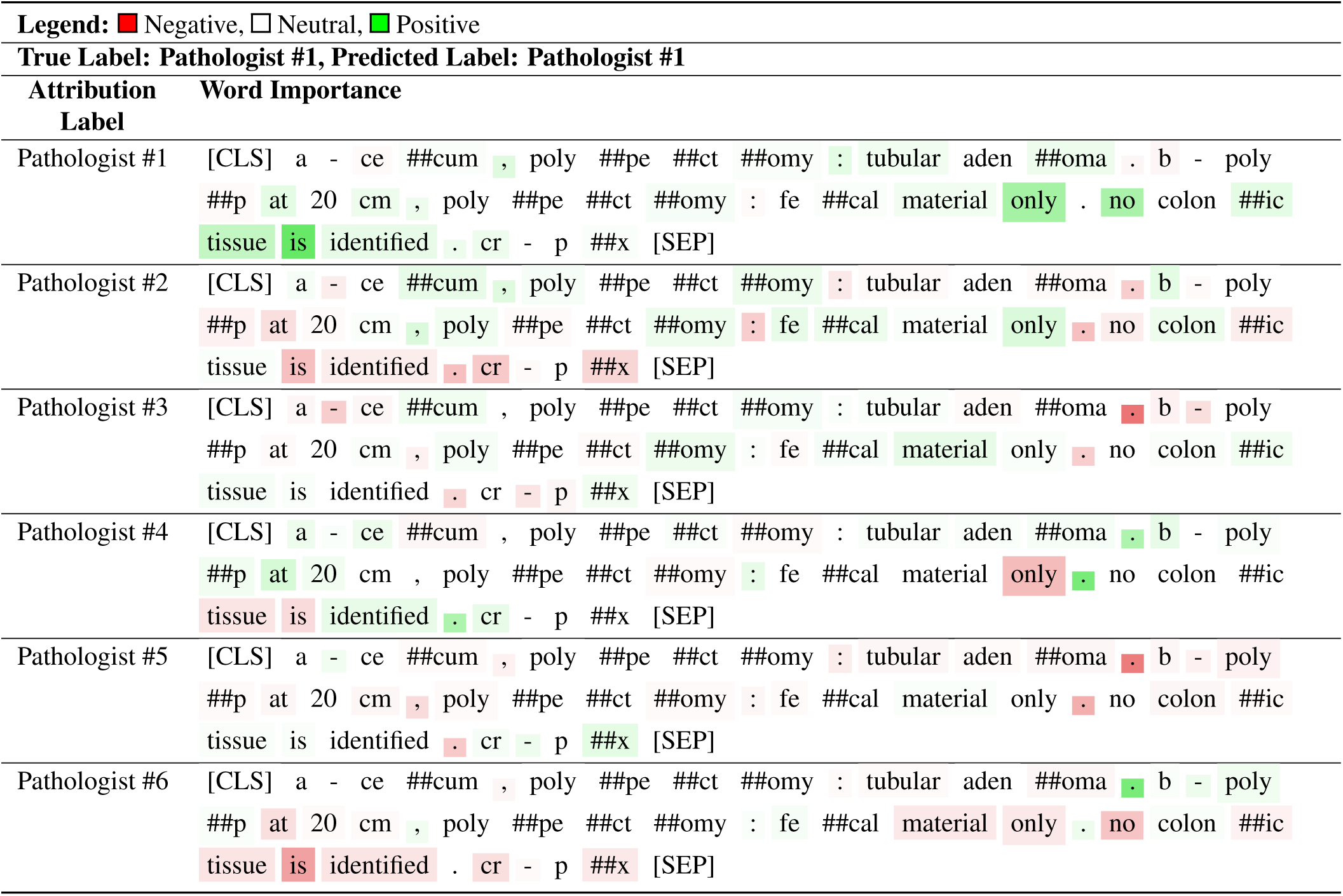
Bert base uncased model attribution interpretation: Pathologist Classification Example 2.

**Table S34.** Clinical Bert model attribution interpretation: Pathologist Classification Example 2.

| Legend: ■ Negative, □ Neutral, ■ Positive |  |
| --- | --- |
| True Label: Pathologist #1, Predicted Label: Pathologist #1 |  |
| Attribution Label | Word Importance |
| Pathologist #1 | [CLS] a - c ##ec ##um , p ##oly ##pect ##omy : tub ##ular ad ##eno ##ma . b<br>- p ##oly ##p at 20 cm , p ##oly ##pect ##omy : f ##eca ##l material only .<br>no co ##lon ##ic tissue is identified . c ##r - p ##x [SEP] |
| Pathologist #2 | [CLS] a - c ##ec ##um , p ##oly ##pect ##omy : tub ##ular ad ##eno ##ma . b<br>- p ##oly ##p at 20 cm , p ##oly ##pect ##omy : f ##eca ##l material only .<br>no co ##lon ##ic tissue is identified . c ##r - p ##x [SEP] |
| Pathologist #3 | [CLS] a - c ##ec ##um , p ##oly ##pect ##omy : tub ##ular ad ##eno ##ma . b<br>- p ##oly ##p at 20 cm , p ##oly ##pect ##omy : f ##eca ##l material only .<br>no co ##lon ##ic tissue is identified . c ##r - p ##x [SEP] |
| Pathologist #4 | [CLS] a - c ##ec ##um , p ##oly ##pect ##omy : tub ##ular ad ##eno ##ma . b<br>- p ##oly ##p at 20 cm , p ##oly ##pect ##omy : f ##eca ##l material only .<br>no co ##lon ##ic tissue is identified . c ##r - p ##x [SEP] |
| Pathologist #5 | [CLS] a - c ##ec ##um , p ##oly ##pect ##omy : tub ##ular ad ##eno ##ma . b<br>- p ##oly ##p at 20 cm , p ##oly ##pect ##omy : f ##eca ##l material only .<br>no co ##lon ##ic tissue is identified . c ##r - p ##x [SEP] |
| Pathologist #6 | [CLS] a - c ##ec ##um , p ##oly ##pect ##omy : tub ##ular ad ##eno ##ma . b<br>- p ##oly ##p at 20 cm , p ##oly ##pect ##omy : f ##eca ##l material only .<br>no co ##lon ##ic tissue is identified . c ##r - p ##x [SEP] |

**Table S35.**
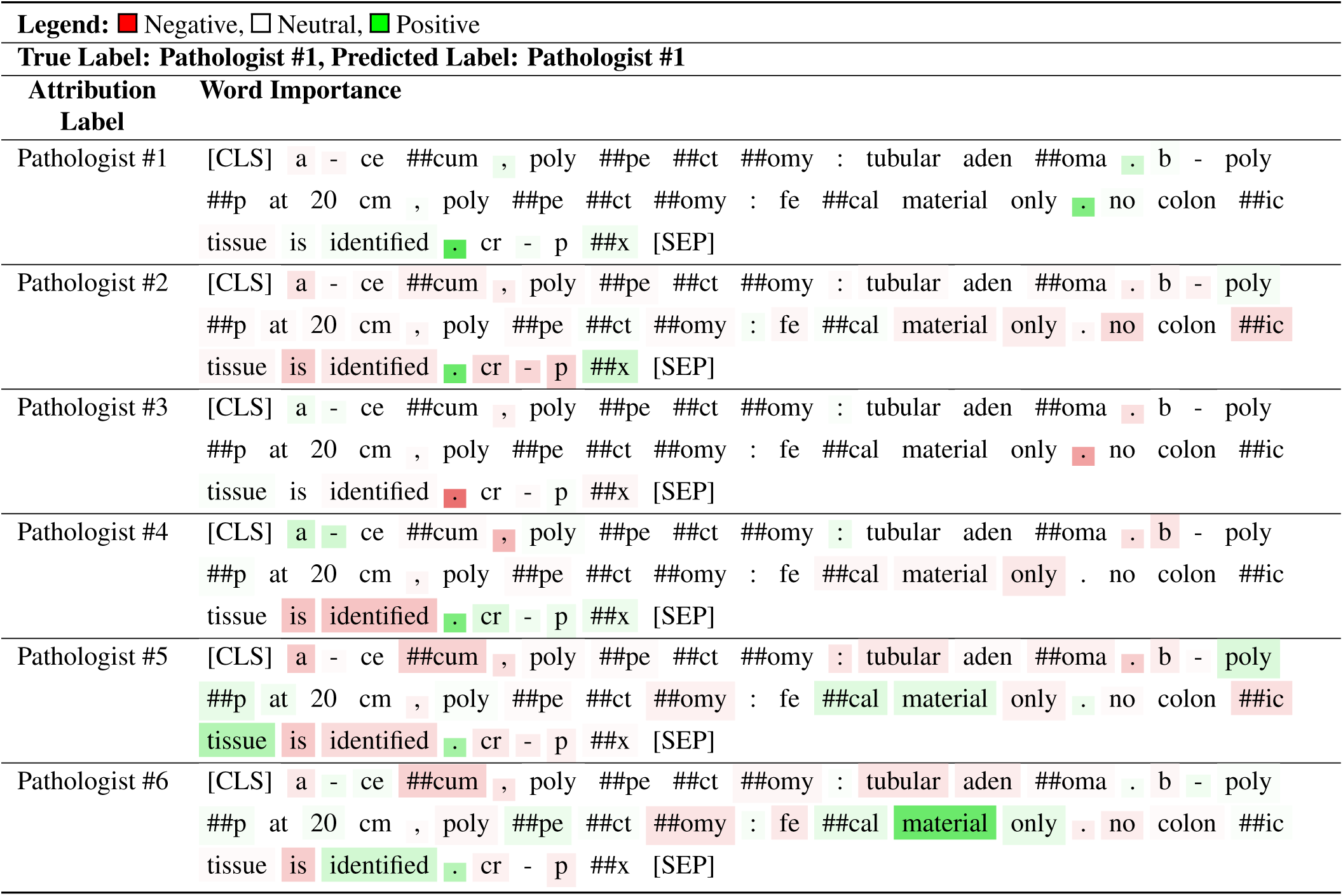
Math Bert model attribution interpretation: Pathologist Classification Example 2.

**Table S36.**
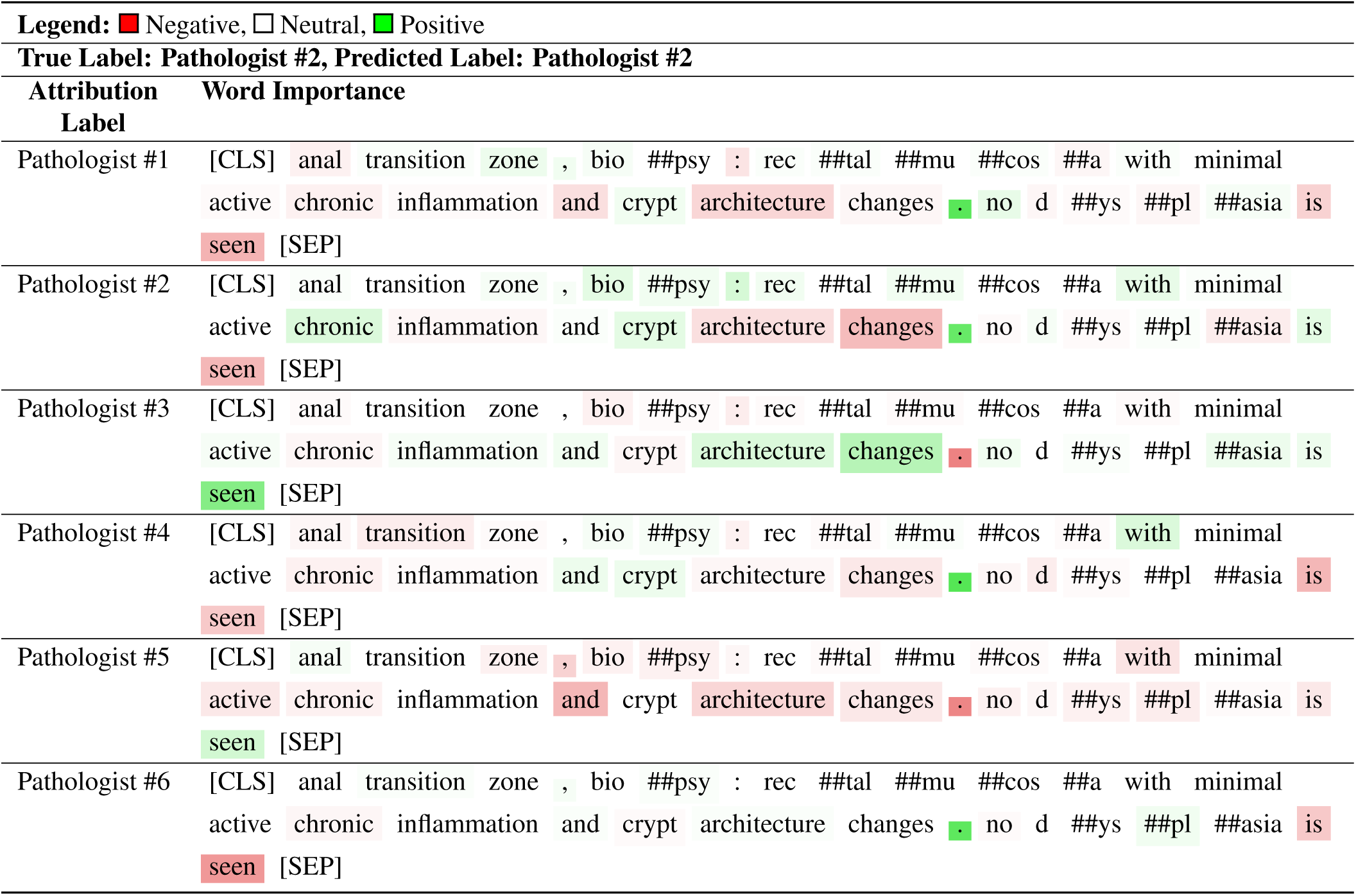
Bert base uncased model attribution interpretation: Pathologist Classification Example 3.

**Table S37.** Clinical Bert model attribution interpretation: Pathologist Classification Example 3.

| Legend: ■ Negative, □ Neutral, ■ Positive |  |
| --- | --- |
| True Label: Pathologist #2, Predicted Label: Pathologist #2 |  |
| Attribution Label | Word Importance |
| Pathologist #1 | [CLS] anal transition zone , bio ##psy : re ##ct ##al ##mu ##cos ##a with minimal active chronic inflammation and cry ##pt architecture changes . no d ##ys ##p ##lasia is seen [SEP] |
| Pathologist #2 | [CLS] anal transition zone , bio ##psy : re ##ct ##al ##mu ##cos ##a with minimal active chronic inflammation and cry ##pt architecture changes . no d ##ys ##p ##lasia is seen [SEP] |
| Pathologist #3 | [CLS] anal transition zone , bio ##psy : re ##ct ##al ##mu ##cos ##a with minimal active chronic inflammation and cry ##pt architecture changes . no d ##ys ##p ##lasia is seen [SEP] |
| Pathologist #4 | [CLS] anal transition zone , bio ##psy : re ##ct ##al ##mu ##cos ##a with minimal active chronic inflammation and cry ##pt architecture changes . no d ##ys ##p ##lasia is seen [SEP] |
| Pathologist #5 | [CLS] anal transition zone , bio ##psy : re ##ct ##al ##mu ##cos ##a with minimal active chronic inflammation and cry ##pt architecture changes . no d ##ys ##p ##lasia is seen [SEP] |
| Pathologist #6 | [CLS] anal transition zone , bio ##psy : re ##ct ##al ##mu ##cos ##a with minimal active chronic inflammation and cry ##pt architecture changes . no d ##ys ##p ##lasia is seen [SEP] |

**Table S38.**
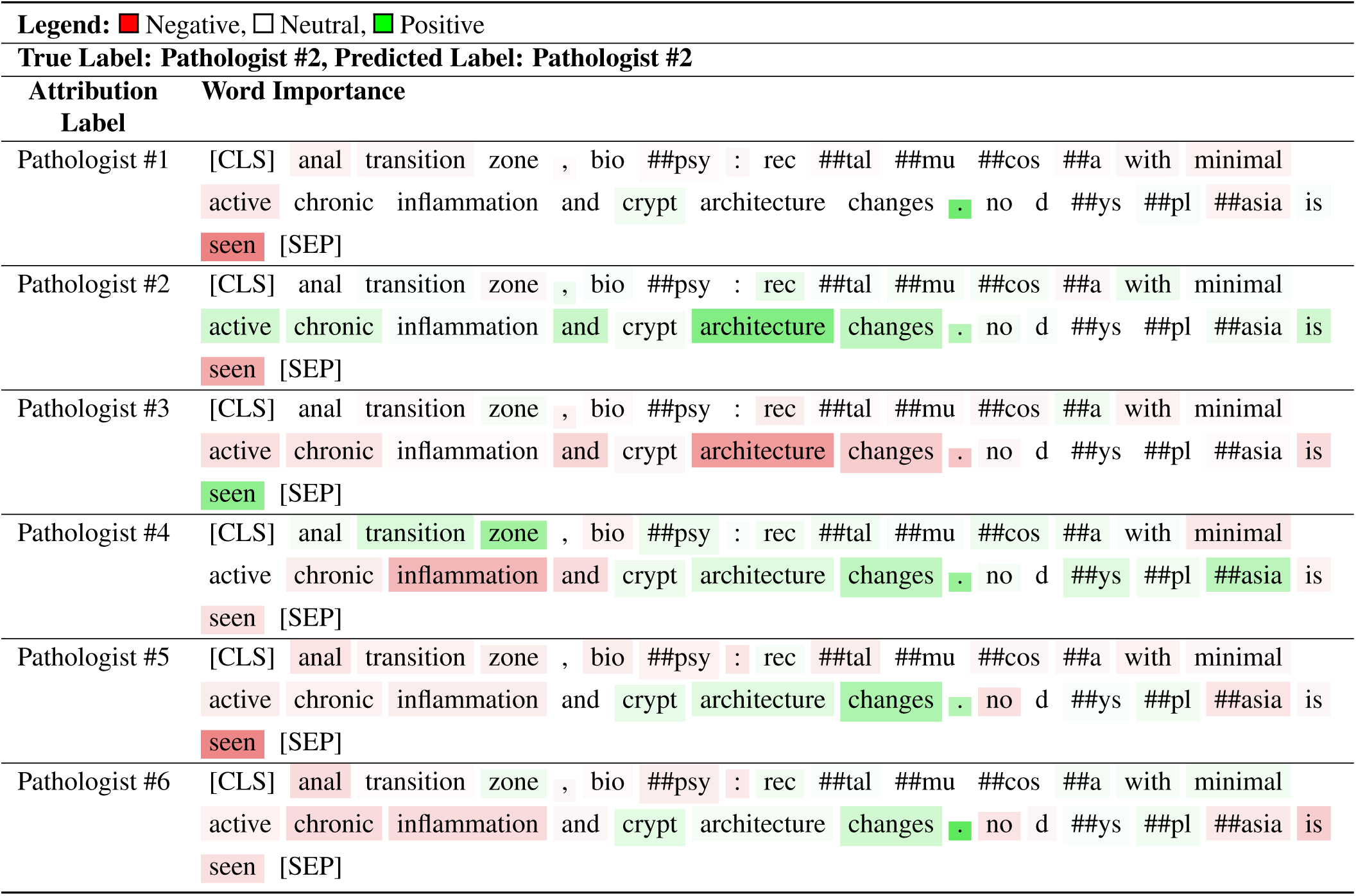
Math Bert model attribution interpretation: Pathologist Classification Example 3.

**Table S39.** Bert base uncased model attribution interpretation: Pathologist Classification Example 4.

| **Legend:** ■ Negative, □ Neutral, ■ Positive |  |  |  |  |  |  |  |  |  |  |  |  |  |  |  |  |  |
| --- | --- | --- | --- | --- | --- | --- | --- | --- | --- | --- | --- | --- | --- | --- | --- | --- | --- |
| **True Label: Pathologist #2, Predicted Label: Pathologist #3** |  |  |  |  |  |  |  |  |  |  |  |  |  |  |  |  |  |
| **Attribution** | **Word Importance** |  |  |  |  |  |  |  |  |  |  |  |  |  |  |  |  |
| **Label** |  |  |  |  |  |  |  |  |  |  |  |  |  |  |  |  |  |
| Pathologist #1 | [CLS] | si | ##gm | ##oid | colon | , | poly | ##pe | ##ct | ##omy | : | tubular | aden | ##oma | cr | - | p |
|  | ##x | [SEP] |  |  |  |  |  |  |  |  |  |  |  |  |  |  |  |
| Pathologist #2 | [CLS] | si | ##gm | ##oid | colon | , | poly | ##pe | ##ct | ##omy | : | tubular | aden | ##oma | cr | - | p |
|  | ##x | [SEP] |  |  |  |  |  |  |  |  |  |  |  |  |  |  |  |
| Pathologist #3 | [CLS] | si | ##gm | ##oid | colon | , | poly | ##pe | ##ct | ##omy | : | tubular | aden | ##oma | cr | - | p |
|  | ##x | [SEP] |  |  |  |  |  |  |  |  |  |  |  |  |  |  |  |
| Pathologist #4 | [CLS] | si | ##gm | ##oid | colon | , | poly | ##pe | ##ct | ##omy | : | tubular | aden | ##oma | cr | - | p |
|  | ##x | [SEP] |  |  |  |  |  |  |  |  |  |  |  |  |  |  |  |
| Pathologist #5 | [CLS] | si | ##gm | ##oid | colon | , | poly | ##pe | ##ct | ##omy | : | tubular | aden | ##oma | cr | - | p |
|  | ##x | [SEP] |  |  |  |  |  |  |  |  |  |  |  |  |  |  |  |
| Pathologist #6 | [CLS] | si | ##gm | ##oid | colon | , | poly | ##pe | ##ct | ##omy | : | tubular | aden | ##oma | cr | - | p |
|  | ##x | [SEP] |  |  |  |  |  |  |  |  |  |  |  |  |  |  |  |

**Table S40.**
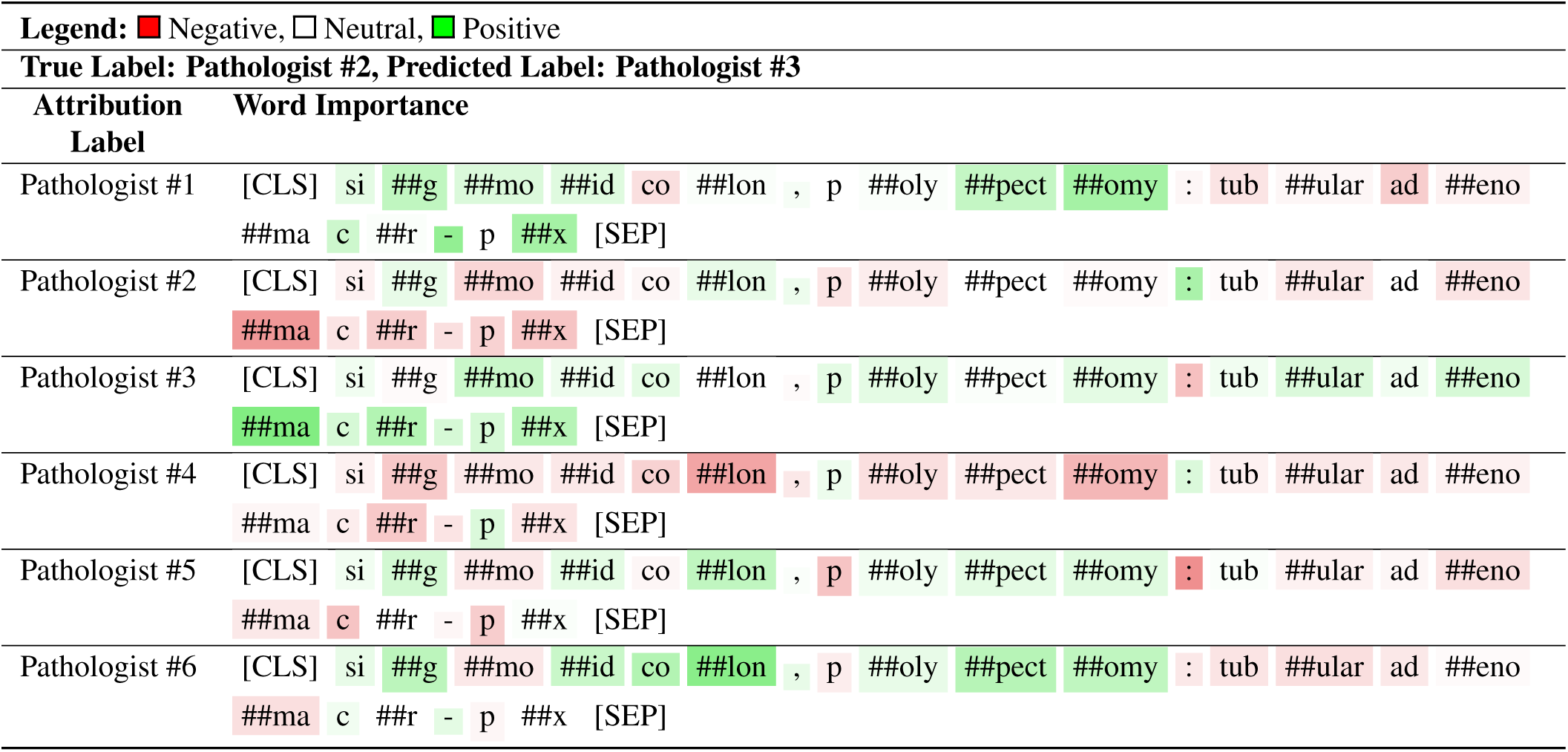
Clinical Bert model attribution interpretation: Pathologist Classification Example 4.

| Legend: ■ Negative, □ Neutral, ■ Positive |  |
| --- | --- |
| True Label: Pathologist #2, Predicted Label: Pathologist #3 |  |
| Attribution Label | Word Importance |
| Pathologist #1 | [CLS] si ##g ##mo ##id co ##lon , p ##oly ##pect ##omy : tub ##ular ad ##eno<br>##ma c ##r - p ##x [SEP] |
| Pathologist #2 | [CLS] si ##g ##mo ##id co ##lon , p ##oly ##pect ##omy : tub ##ular ad ##eno<br>##ma c ##r - p ##x [SEP] |
| Pathologist #3 | [CLS] si ##g ##mo ##id co ##lon , p ##oly ##pect ##omy : tub ##ular ad ##eno<br>##ma c ##r - p ##x [SEP] |
| Pathologist #4 | [CLS] si ##g ##mo ##id co ##lon , p ##oly ##pect ##omy : tub ##ular ad ##eno<br>##ma c ##r - p ##x [SEP] |
| Pathologist #5 | [CLS] si ##g ##mo ##id co ##lon , p ##oly ##pect ##omy : tub ##ular ad ##eno<br>##ma c ##r - p ##x [SEP] |
| Pathologist #6 | [CLS] si ##g ##mo ##id co ##lon , p ##oly ##pect ##omy : tub ##ular ad ##eno<br>##ma c ##r - p ##x [SEP] |

**Table S41.**
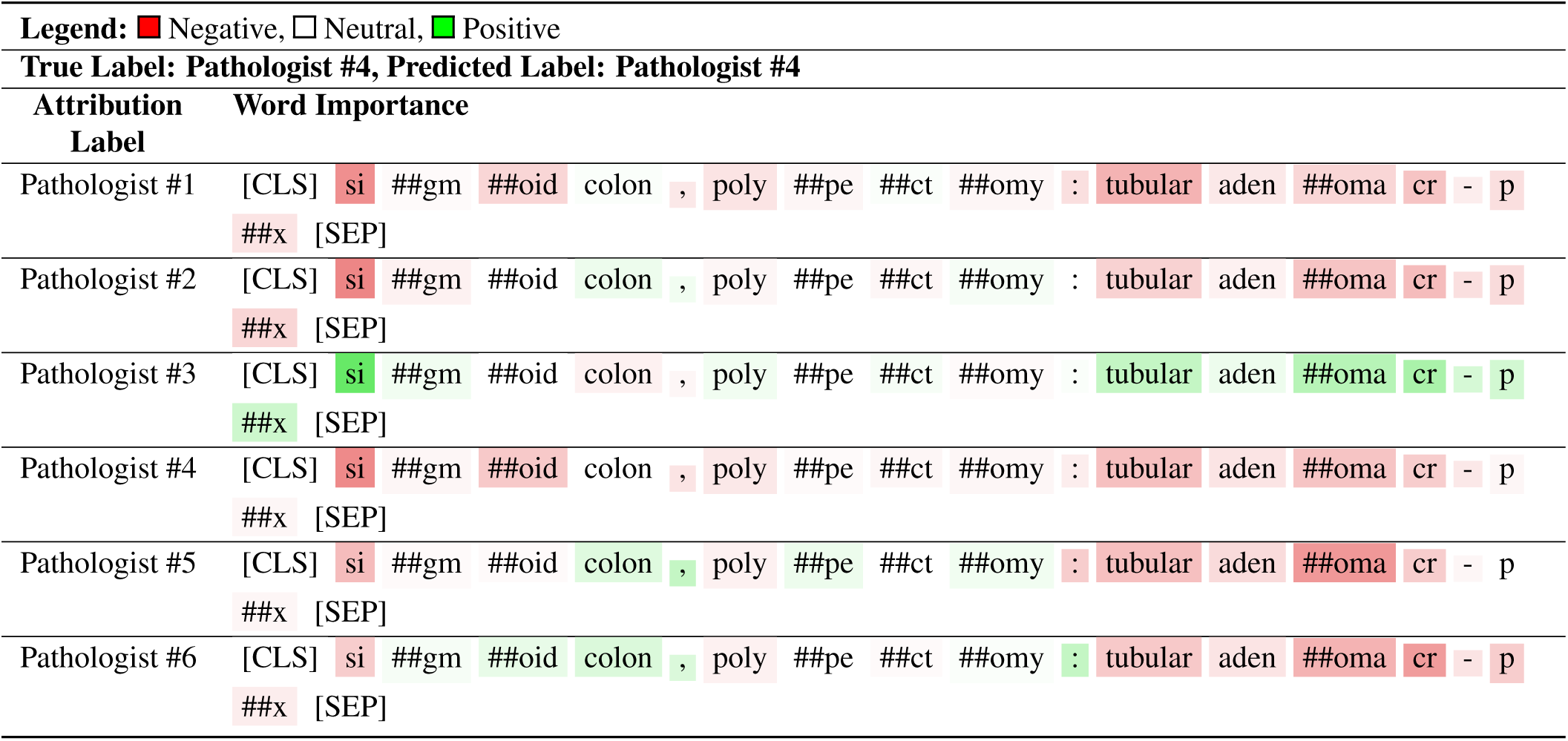
Math Bert model attribution interpretation: Pathologist Classification Example 4.

**Table S42.**
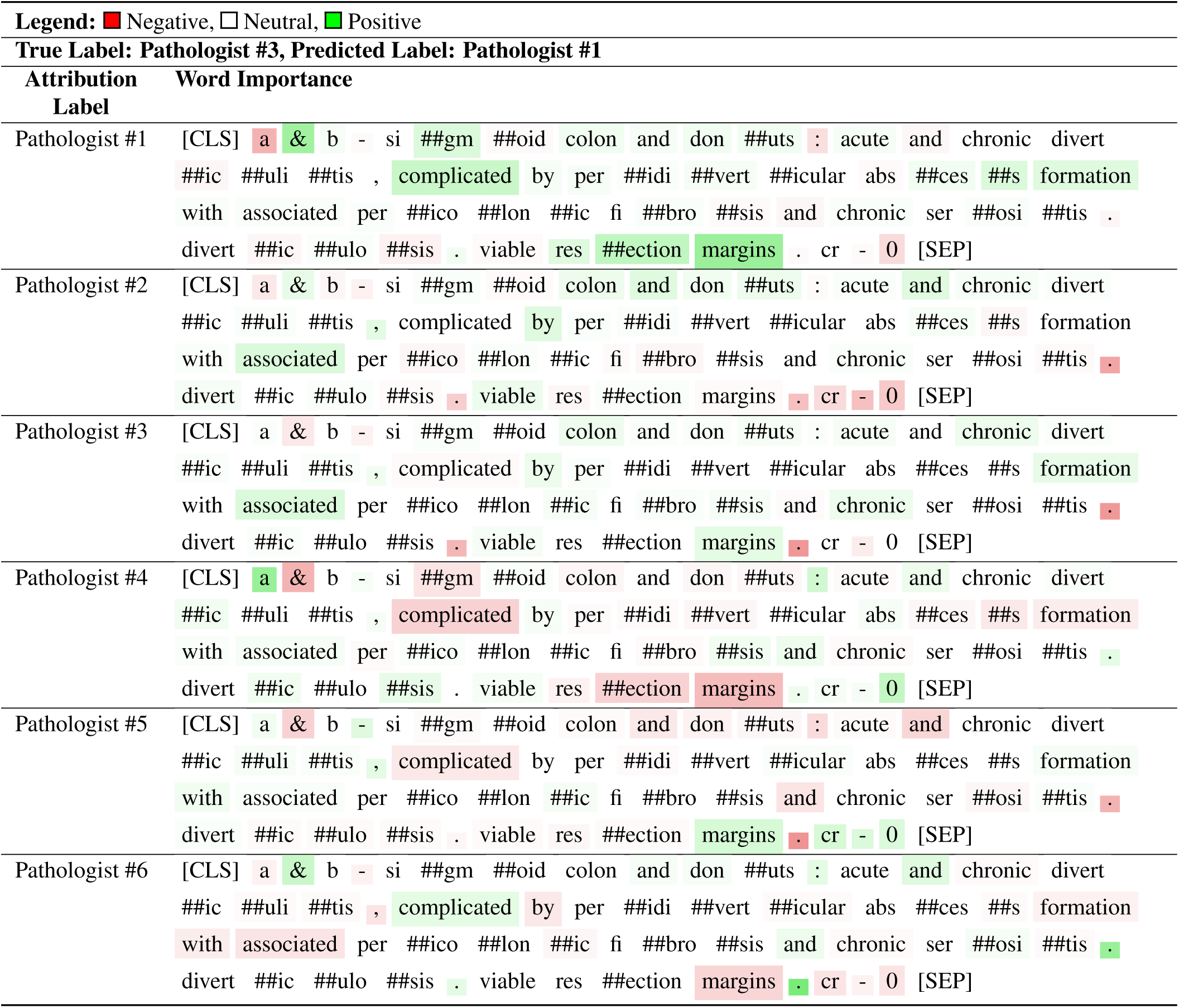
Bert base uncased model attribution interpretation: Pathologist Classification Example 5.

**Table S43.**
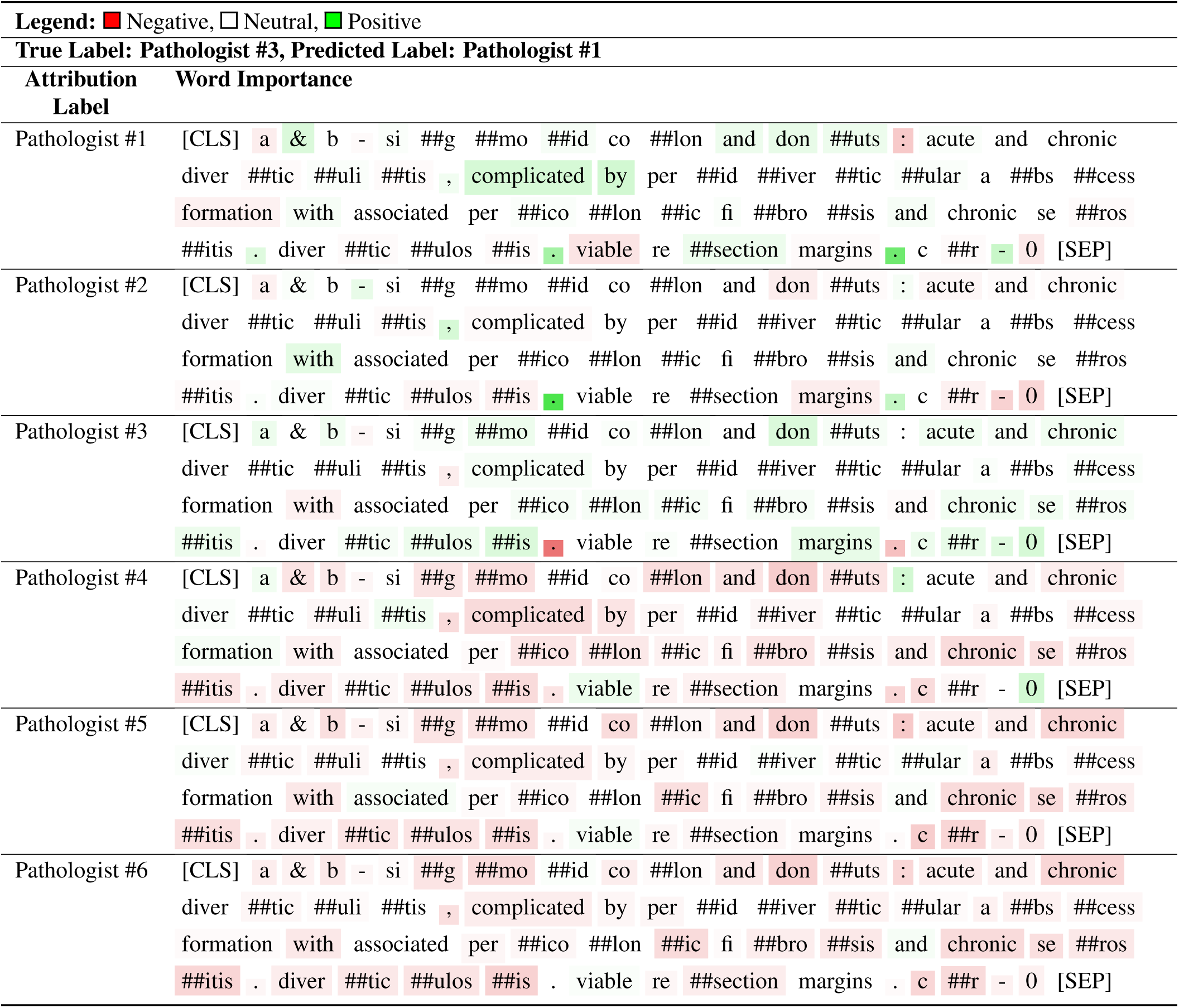
Clinical Bert model attribution interpretation: Pathologist Classification Example 5.

**Table S44.**
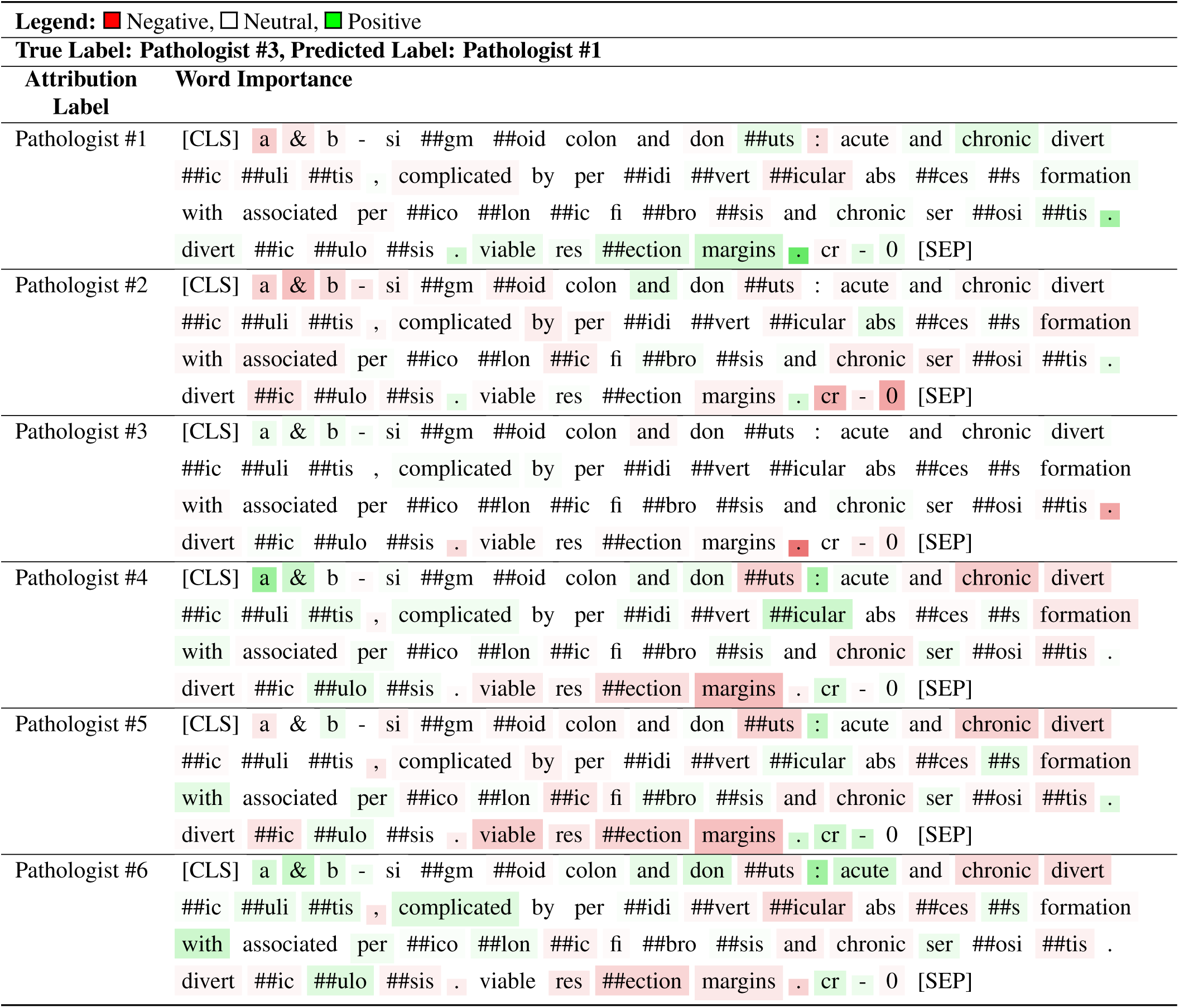
Math Bert model attribution interpretation: Pathologist Classification Example 5.

**Table S45.**
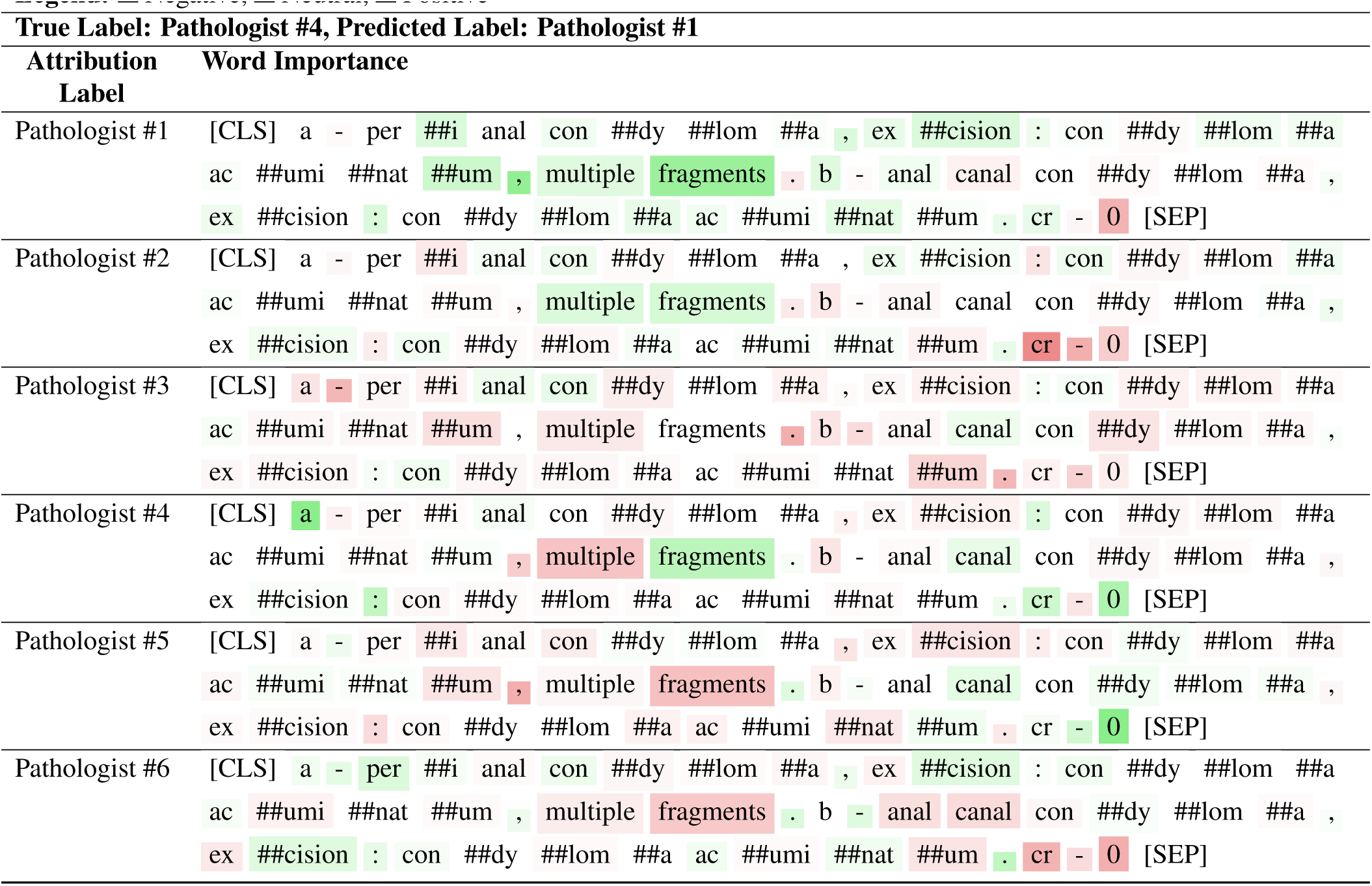
Bert base uncased model attribution interpretation: Pathologist Classification Example 6.

**Table S46.**
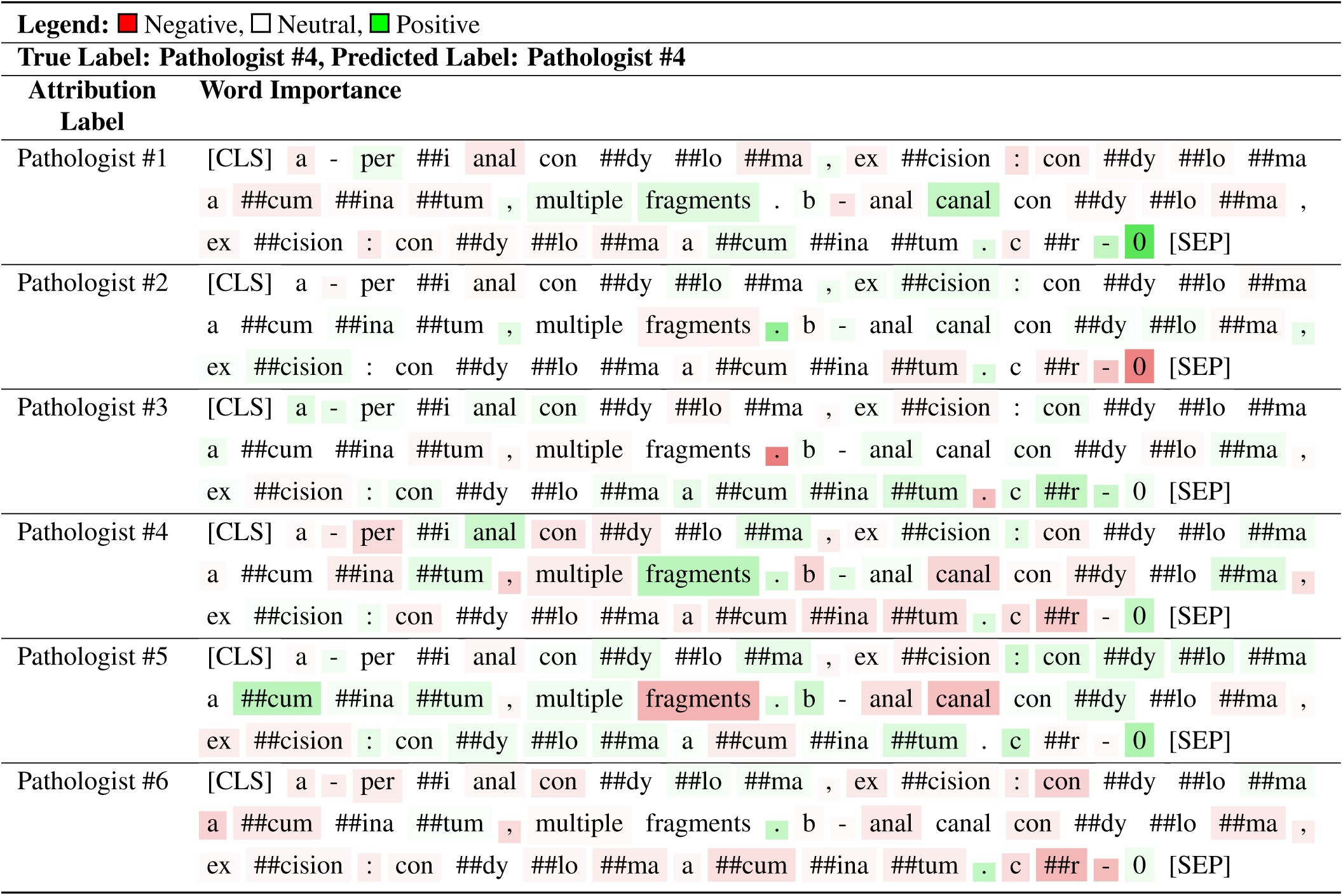
Clinical Bert model attribution interpretation: Pathologist Classification Example 6.

**Table S47.**
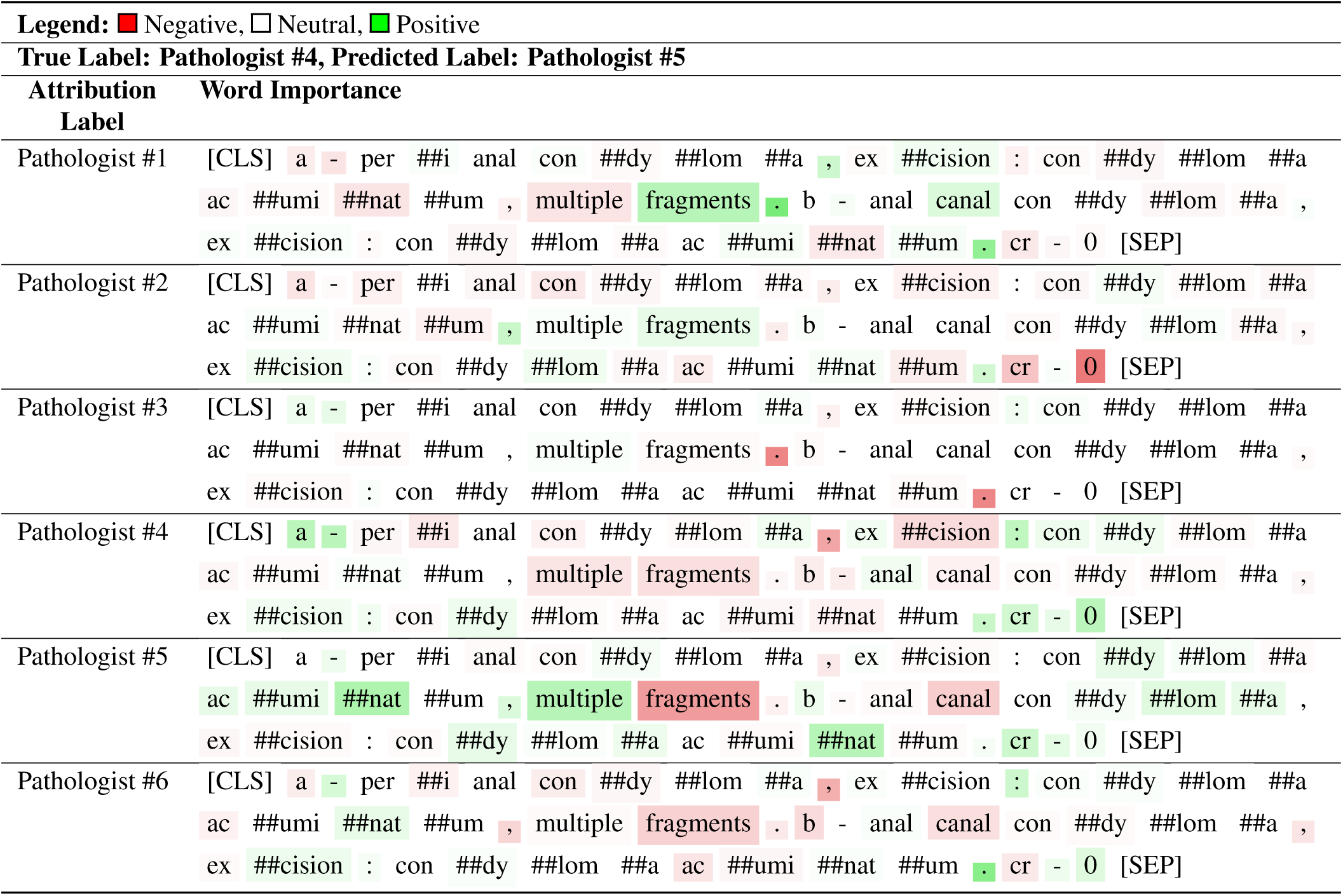
Math Bert model attribution interpretation: Pathologist Classification Example 6.

